# Quantifying associations of genotype, proteinuria and eGFR with long-term kidney outcomes in Alport Syndrome using data from the UK National Registry of Rare Kidney Diseases (RaDaR)

**DOI:** 10.64898/2026.06.08.26355110

**Authors:** Katie Wong, David Pitcher, Sherry Masoud, Konstantinos Tzoumkas, Angela Branson, Thomas Oates, Susie Gear, Hannah Russell, RaDaR Consortium, Klaus Francke, Elif Inan-Eroglu, Khaled Abdelgawwad, Shiguang Liu, Pronabesh Dasmahaptra, Julie Lin, Alex Mercer, Bruce Hendry, Rachel Lennon, A. Neil Turner, Daniel P. Gale

**Affiliations:** Centre for Kidney and Bladder Health, University College London, UK and National Registry of Rare Kidney Diseases, Bristol, UK; Centre for Kidney and Bladder Health, University College London, UK; Manchester University NHS Foundation Trust; Royal London Hospital, UK; Alport UK; Multiple Affiliations; Bayer Pharmaceuticals, Berlin, Gemany; Rare Disease and Rare Blood Disorders Development, Sanofi, Cambridge, MA, USA; Travere Therapeutics, Inc., San Diego, California; JAMCO Pharma Consulting, Stockholm, Sweden; Manchester Cell-Matrix Centre, Faculty of Biology, Medicine and Health, The University of Manchester, UK; University of Edinburgh, Edinburgh, UK

**Author notes:** Corresponding author: Professor Daniel P Gale, Department of Renal Medicine, Royal Free Hospital, Rowland Hill Street, London, NW3 2PF.

## Abstract

**Background:** Alport Syndrome (AS), caused by pathogenic variants in type IV collagen genes *COL4A3/4/5*, is a leading monogenic cause of Kidney Failure (KF). Clinical course varies widely, and disease-specific predictors of progression relevant to clinical care and trial design remain incompletely defined.

**Methods:** In this retrospective cohort study of individuals with AS in the UK National Registry of Rare Kidney Diseases, patients were classified as having AS or heterozygous genotypes and followed to assess proteinuria progression, eGFR slope and kidney survival. Proteinuria and eGFR trajectories were analysed using mixed-effects regression models; kidney survival using Kaplan–Meier analysis.

**Results:** Among 1032 participants (median follow-up 11.6 years; 47% female), 475 (46%) had AS genotypes (Male XLAS or autosomal recessive AS). eGFR decline accelerated with advancing CKD stage across all genotypes (p<0.001). Proteinuria increased as eGFR declined and occurred earlier in AS genotypes. After reaching proteinuria thresholds of ≥1.0 and ≥3.0g/g, kidney survival over the subsequent 5-years did not differ significantly between genotypes (log-rank p=0.14, p=0.17, respectively), although modest differences emerged over longer follow-up. Across eGFR thresholds (90, 60, and 45mL/min/1.73m²), higher proteinuria was associated with shorter time to KF; for example, at eGFR 45mL/min/1.73m², median time to KF was 3.0 years (IQR, 1.6–5.4) for above-median vs 6.5 years (5.1–not estimable) for below-median proteinuria (p<0.0001). Almost all patients who reached KF had developed proteinuria ≥0.3g/g.

**Conclusion:** In this national cohort, eGFR decline accelerated with CKD stage and proteinuria was strongly associated with progression to KF across genotypes. The non-linearity of eGFR decline may inform its interpretation in clinical practice and use as a trial endpoint. Once comparable proteinuria levels were reached, differences in outcomes by genotype were attenuated, supporting proteinuria as a key prognostic marker and strengthening rationale for its use as a surrogate endpoint in AS clinical trials.

**Key points:**

- In this cohort study of 1032 individuals with Alport syndrome, eGFR decline accelerated with advancing CKD stage across all genotypes.
- Proteinuria increased as kidney function declined and was strongly associated with time to kidney failure across a range of eGFR thresholds.
- Once comparable levels of proteinuria were reached, short-term kidney outcomes were similar across genotypes, which may inform interpretation in clinical practice and use as a surrogate endpoint in AS clinical trials.

## Introduction

Alport Syndrome (AS) is the second most common monogenic cause of kidney failure (KF), and is caused by variants in the *COL4A3/COL4A4*/*COL4A5* genes, which encode the collagen (IV) *α*3/*α*4/*α*5 chains that trimerise to form an essential type IV collagen network in the glomerular basement membrane. Defects in this network are strongly associated with haematuria and many patients develop progressive proteinuria, kidney function decline and KF.

Clinical outcomes vary widely, even within families, and limited long-term prognostic data can leave patients and clinicians uncertain and anxious about expected disease course^1^. Genotype-phenotype correlations partly explain this variability. *COL4A5* variants cause X-linked AS (XLAS), whereas *COL4A3/4* variants cause autosomal AS. Previous genotype-phenotype studies have shown differences in kidney outcomes for different AS genotypes; Males with XLAS and individuals with biallelic *COL4A3* or *COL4A4* pathogenic variants (autosomal recessive AS, ARAS) generally experience earlier and more frequent KF than those with single heterozygous variants. In male XLAS, truncating variants have been associated with earlier age at KF compared to missense variants^2–5^. Evidence in other Alport genotypes is more variable^6–8^, with some studies suggesting that monoallelic (single heterozygous) non-truncating variants may confer worse outcomes than monoallelic truncating variants. Nearly half of variants causing AS result in glycine substitutions, with certain molecular characteristics (e.g. proximity to non-collagenous interruptions) linked with milder phenotypes^9^.

Data on rate of kidney function decline prior to KF in AS, and whether this differs by genotype or across disease stage, are limited. Previous studies have reported a faster eGFR decline for male XLAS versus individuals with heterozygous *COL4A3*/4 variants, but were limited by small sample size, incomplete genotype/phenotype data, or <3-year follow-up^10–12^.

Proteinuria is associated with adverse kidney outcomes^13^, but whilst its prognostic value in mostly non-monogenic glomerular disorders such as IgA nephropathy (IgAN)^14,15^ idiopathic nephrotic syndrome (INS)^16^, and immune-complex MPGN/C3 Glomerulopathy^17,18^ has been well delineated, there are minimal studies reporting longitudinal trajectories of proteinuria^19^ in AS. Previous studies have utilised “baseline” proteinuria values at arbitrary time-points (e.g. “first clinic visit”)^10,13,20^; these values may represent patients at different points in their disease course and with differing levels of kidney function, making these results difficult to interpret.

This question has become more important with recent therapeutic developments in AS include a clinical trial of lademirsen^21^, and studies evaluating broader proteinuria-lowering therapies such as SGLT2-inhibitors^22^ and endothelin-targeted therapies^22–24^. Meta-analyses in other kidney diseases support short-term changes in eGFR slope and proteinuria as surrogate end-points for long-term KF ^15,25–29^, but whether, and how, these findings can be extrapolated to AS remains uncertain.

We therefore examined associations between genotype, proteinuria, eGFR and kidney outcomes in 1032 individuals with AS recruited to the UK National Registry of Rare Kidney Diseases (RaDaR).

## Methods

### Data source and study population

RaDaR recruits patients with rare kidney disease from 108 UK National Health Service sites with informed consent. AS recruitment began in 2013. Data were extracted on August 12, 2025. Study design, eligibility criteria, data linkage, and variable definitions have been previously reported^30–32^, with details in Supplementary Methods. All patients were recruited with a clinical diagnosis from their treating clinician consistent with one of the defined eligibility criteria.

### Genetically confirmed diagnoses

Patients were included in the ‘Genetically Confirmed’ group if testing in the patient or a parent identified a variant classified as pathogenic (P) or likely pathogenic (LP) by American College of Medical Genetics criteria^33^ (Supplementary Figure 1). The ‘Clinical Diagnosis’ group comprised patients where diagnosis was made based on family history, histopathology and clinical features, with genetic testing either not performed or where no P/LP variant was identified.

Patients in both groups were then categorised as having either an ‘Alport Syndrome’ (male XLAS or homozygous or biallelic *COL4A3*/*COL4A4* variants, ARAS) or ‘Heterozygous’ genotypes, (heterozygous *COL4A3*/*COL4A4* variants, TBMN and female XLAS), with the definition of Alport Syndrome genotypes consistent with current naming guidance^34^.

### Genotyped cohort

Where clinical genetic reports were available to review, variants were classified into Protein length-altering or Non-protein length-altering. Where >1 variants were present, the variant predicted to be most damaging was analysed, consistent with inclusion criteria for clinical trials. Glycine-substituting missense variants were further classified by position and predicted structural effect. Variant details were cross-checked against contemporary databases (details in Supplementary Methods).

### eGFR, proteinuria and KF

KF was defined as initiation of chronic kidney replacement therapy (KRT), eGFR ≤15mL/min/1.73m^2^ maintained for ≥4 weeks^35^ or death. Definitions of diagnosis date, follow-up and time-averaged proteinuria (TAP) are detailed in Supplementary Methods. eGFR was calculated using the European Kidney Function Consortium equation^36^. Proteinuria threshold attainment was defined as the first value above threshold. Sensitivity analyses using alternative eGFR equations and proteinuria definitions are described in supplementary materials.

### Statistical analyses

Kaplan-Meier analysis and log-rank testing were used to compare age at KF and time from diagnosis to KF by genotype and variant category. Time to KF from sustained eGFR thresholds below 90, 60 and 45ml/min/1.73m^2^ were estimated, stratified by median proteinuria in preceding year.

The proportion of patients within each CKD stage, and with proteinuria above a certain level, by age are displayed in stacked area plots. eGFR slope and proteinuria trajectories were modelled using mixed-effects regression (details in Supplementary Methods).

Available data are presented in Supplementary Table 1; analyses used complete cases.

This study is reported according to Strengthening the Reporting of Observational Studies in Epidemiology guidelines.

## Results

### Demographics

1032 patients with AS were included, aged 3 to 92 years, n=481 (47%) female. Median follow-up was 11.6 years (IQR 7.6-19.1).

Baseline characteristics are presented in Table 1 and Supplementary Table 2 for patients in the genetically confirmed (n=603, 58%) and clinically diagnosed (n=429, 42%) cohorts. Patients with clinical diagnoses were older, had been diagnosed earlier and had experienced more KF events than those with genetically confirmed diagnoses (Supplementary Figure 2, Supplementary Table 3) Within the clinically diagnosed cohort, 238/429 (55%) were diagnosed with Alport Syndrome genotypes (Male XLAS or ARAS), and 191/429 (45%) heterozygous genotypes (Female XLAS, *COL4A3/4* heterozygotes and TBMN). In the genetically confirmed cohort, 237/603 (39%) had Alport Syndrome genotypes and 366/603 (61%) heterozygous genotypes.

**Table 1:** Clinical demographics.

|  | Genetically confirmed |  |  |  | Clinical Diagnosis |  |  |  |
| --- | --- | --- | --- | --- | --- | --- | --- | --- |
|  | Female XLAS and heterozygotes |  | Alport Syndrome |  | Female XLAS and heterozygotes |  | Alport Syndrome |  |
|  | Female XLAS | COL4A3/4 heterozygous | Male XLAS | ARAS | Female XLAS | TBMN | Male XLAS | ARAS |
|  | n=172 | n=194 | n=199 | n=38 | n=85 | n=106 | n=218 | n=20 |
| Median follow-up time (Years, IQR) | 9.8 (6.9-13.5) | 8.2 (4.4-10.9) | 10.5 (6.7-13.0) | 11.1 (6.4-13.8) | 22.7 (16.1-31.4) | 12.6 (8.4-21.3) | 23.8 (12.3-33.5) | 16.8 (12.1-31.9) |
| Median current age (Years, IQR) | 34.6 (19.3-48.8) | 41.6 (21.8-58.4) | 26.6 (17.1-41.9) | 25.8 (17.3-36.6) | 57.7 (41.1-67.1) | 53.7 (39.8-64.9) | 52.5 (38.9-61.8) | 40.1 (29.9-48.0) |
| Female (%) | 172(100.0) | 121(62.4) | 0 (0) | 20(52.6) | 85(100.0) | 73(68.9) | 0 (0) | 10(50.0) |
| **Ethnicity (n, %)** |  |  |  |  |  |  |  |  |
| White | 115 (67) | 117 (60) | 133 (67) | 21 (55) | 62 (73) | 71 (67) | 177 (81) | 16 (80) |
| Not reported | 42 (24) | 43 (22) | 49 (25) | 10 (26) | 13 (15) | 23 (22) | 22 (10) | 3 (15) |
| **Patient status at diagnosis** |  |  |  |  |  |  |  |  |
| Age at diagnosis (Years, IQR) | 23.7 (7.2-34.6) | 32.3 (13.4-49.7) | 15.3 (7.5-30.4) | 15.2 (7.0-29.4) | 30.2 (18.0-43.9) | 36.8 (25.1-48.5) | 22.4 (10.0-36.7) | 18.2 (5.5-28.0) |
| eGFR at diagnosis (ml/min/1.73m<sup>2</sup>, 95% CI) | 102.0 (65.1-111.7) | 88.7 (52.0-108.3) | 79.5 (37.0-111.0) | 74.6 (40.7-111.9) | 60.4 (24.1-101.7) | 80.0 (47.8-109.2) | 55.3 (19.6-98.2) | 65.4 (25.3-111.1) |

Alport Syndrome=Male XLAS or Autosomal Recessive Alport Syndrome (ARAS)-2 likely pathogenic or pathogenic variants in either COL4A3 or COL4A4, confirmed *in trans* in Genetically confirmed group, phase not determined in Clinical diagnosis group.

### Genotype

Within the genetically confirmed cohort, 478/603 (79%) patients had detailed genotyping data available to review (‘Genotyped Cohort’); n=194 had Alport Syndrome (n=30 biallelic *COL4A3/4* variants or ARAS, n=164 Male XLAS), and n=284 heterozygous genotypes (n=130 Female XLAS, n=154 *COL4A3/4* heterozygotes), Supplementary Figure 1. Patients with two affected Alport genes or *MYH9* variants were excluded.

Overall, 214/478 (45%) patients had pathogenic variants predicted to alter protein length (‘protein length-altering variants’), and 264/478 had missense variants or small insertions/deletions (‘non-protein length-altering variants’). 225/264 (85%) patients had missense variants resulting in glycine substitutions (molecular characteristics in Supplementary Table 4).

### Diagnosis

Individuals with Alport syndrome genotypes were younger at diagnosis and had lower eGFR than patients with heterozygous genotypes in both the genetically confirmed and clinically diagnosed cohorts (Table 1).

### Kidney Failure and death

411/1032 (40%) patients reached KF during follow-up (Table 2), n=149 in the genetically confirmed cohort. Across all patients, most started KRT on peritoneal dialysis (n=149, 36%, Table 2). Initial KRT modality did not differ by genotype (p=0.81).

**Table 2:** Kidney outcomes.

|  | Genetically confirmed |  |  |  | Clinical Diagnosis |  |  |  |
| --- | --- | --- | --- | --- | --- | --- | --- | --- |
|  | Female XLAS and heterozygotes |  | Alport Syndrome |  | Female XLAS and heterozygotes |  | Alport Syndrome |  |
|  | Female XLAS | COL4A3/4 heterozygous | Male XLAS | ARAS | Female XLAS | TBMN | Male XLAS | ARAS |
|  | n=172 | n=194 | n=199 | n=38 | n=85 | n=106 | n=218 | n=20 |
| KF events | 17(9.9) | 34(17.5) | 82(41.2) | 16(42.1) | 44(51.8) | 25(23.6) | 179(82.1) | 14(70.0) |
| Haemodialysis | 6(35.3) | 11(32.4) | 26(31.7) | 4(25.0) | 14(31.8) | 9(36.0) | 69(38.5) | 2(14.3) |
| Peritoneal Dialysis | 3(17.6) | 12(35.3) | 30(36.6) | 8(50.0) | 21(47.7) | 7(28.0) | 63(35.2) | 5(35.7) |
| Transplant | 8(47.1) | 11(32.4) | 26(31.7) | 4(25.0) | 9(20.5) | 9(36.0) | 47(26.3) | 7(50.0) |
| 25<sup>th</sup> centile age at KF (years, IQR) | 57.2 (44.2-NE) | 54.7 (46.9-64.9) | 22.7 (19.7-25.1) | 17.7 (14.6-23.4) | 38.8 (29.5-41.0) | 59.1 (51.6-65.2) | 20.7 (20.0-21.8) | 20.5 (13.8-24.3) |
| 25<sup>th</sup> percentile time from diagnosis to KF (years, IQR) | 37.7 (NE) | 14.2 (11.1- NE) | 8.4 (5.9-10.2) | 7.3 (2.9- 16.4) | 7.5 (3.2- 14.8) | 18.2 (12.4-25.9) | 2.8 (1.2- 5.1) | 11.6 (0.8-21.7) |
| 10-year kidney survival from diagnosis (years, IQR) | 0.90 (0.82-0.95) | 0.86 (0.79-0.91) | 0.69 (0.58-0.77) | 0.62 (0.39-0.79) | 0.72 (0.60-0.81) | 0.89 (0.80-0.94) | 0.60 (0.52-0.67) | 0.81 (0.51-0.93) |
| 25<sup>th</sup> percentile time from eGFR 90 to 30 ml/min/1.73m<sup>2</sup> (years, IQR) | 14.2 (6.2-.) | NE | 4.6 (2.1-5.6) | 4.8 (0.3-6.7) | 8.0 (1.94-NE) | 16.5 (5.95-NE) | 2.8 (1.5-NE) | NE |
| 10-year survival from reaching eGFR 30 from 90ml/min/1.73m<sup>2</sup> | 0.91 (0.51-0.99) | 0.78 (0.32-0.95) | 0.34 (0.12-0.57) | 0.40 (0.10-0.69) | NE | 0.86 (0.33-0.98) | 0.52 (0.19-0.77) | NE |

Alport Syndrome=Male XLAS or Autosomal Recessive Alport Syndrome (ARAS)-2 likely pathogenic or pathogenic variants in either COL4A3 or COL4A4- confirmed *in trans* in Genetically confirmed group- phase not determined in Clinical diagnosis group. NE= Not estimable

Time from diagnosis to KF varied. Across both genetically confirmed and clinically diagnosed cohorts, males with XLAS and individuals with ARAS or heterozygous *COL4A3/4* variants reached KF within an estimated 3–18 years (25th centile estimates). In contrast, females with genetically confirmed XLAS had a significantly longer time to KF (38 years; p<0.0001, Figure 1b), likely reflecting earlier diagnosis through familial screening.

**Figure 1:**
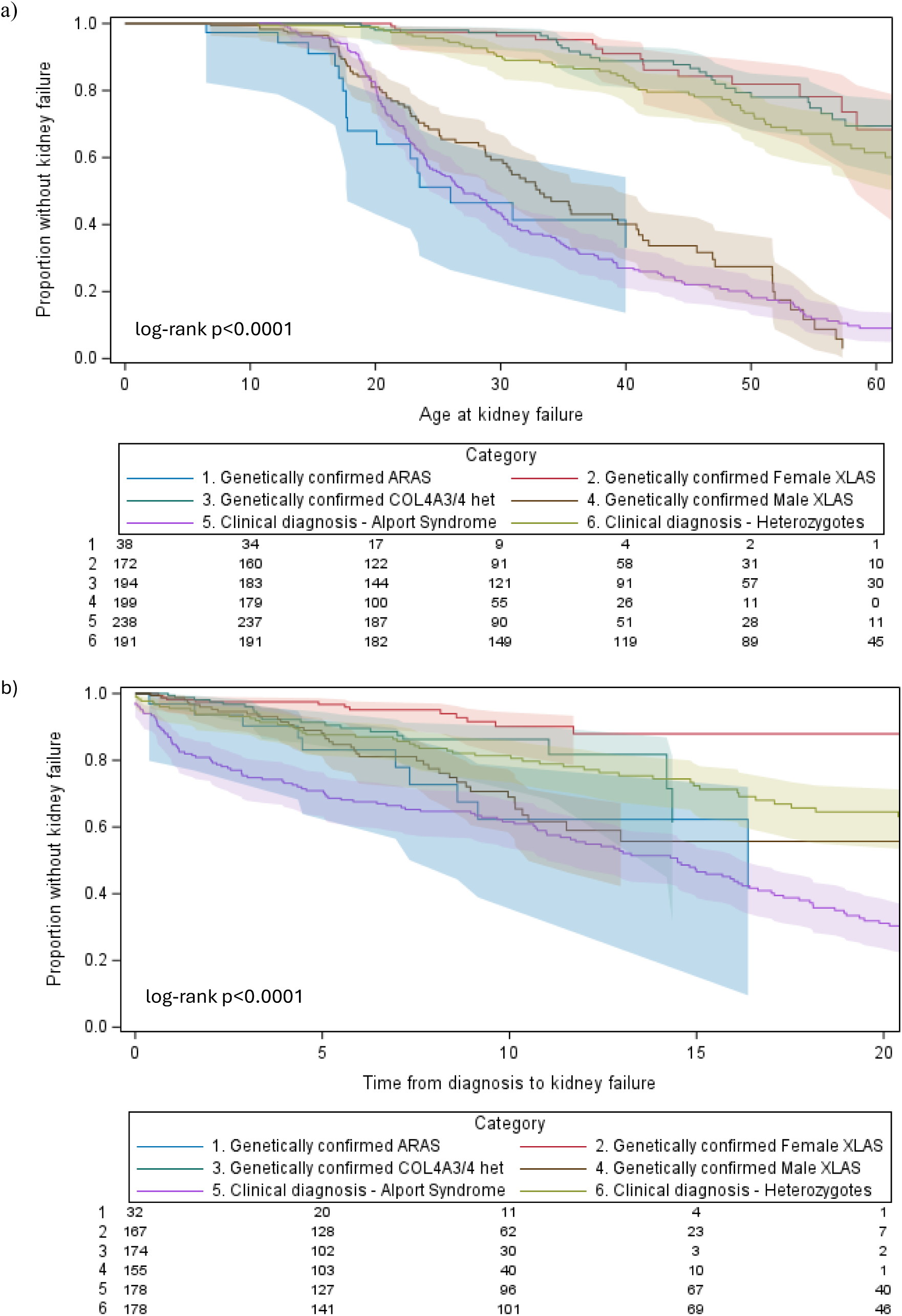
Age at kidney failure (a) and time in years from diagnosis to kidney failure (b) by genetically confirmed and clinically diagnosed subgroupings

Age at KF differed by genotype (p<0.0001, Figure 1a, Supplementary Figure 3). Within the genetically confirmed group, those with ARAS and Male XLAS had the youngest ages at KF (25^th^ centile estimates of 18 and 23 years, respectively), with similar findings in those clinically diagnosed (Table 2). n=58 patients across all groups died during follow-up (6%), and of these only n=3 (0.3%) prior to KF.

### Effect of pathogenic variant type

Within the genotyped cohort, having an Alport Syndrome genotype and protein length-altering variant was associated with a younger median age at KF compared to those with non-protein length-altering variants: 30.2 (23.4-35.6) vs 41.8 (30.5-51.7), p=0.0098 (Figure 2a), mainly driven by differences in males with XLAS (Supplementary Figure 4). For those with Alport syndrome genotypes, stop-gain variants were associated with earliest median age at KF: 23 years (95% CI 17.1–NE), compared with 31 years (23.5–39.4) for other protein length-altering and 42 years (30.5–51.7) for non-protein length-altering variants (p=0.034, Supplementary Figure 5).

**Figure 2:**
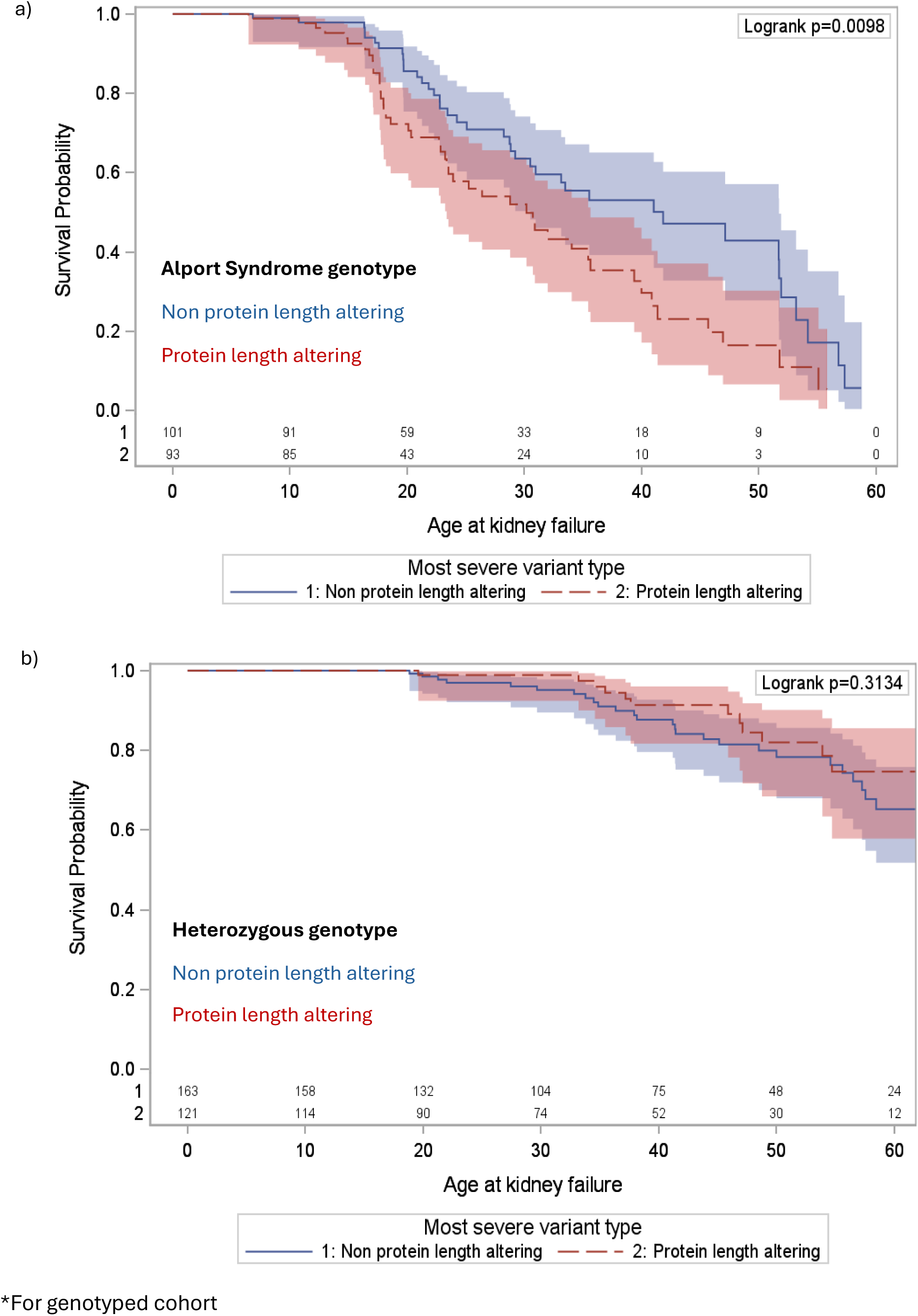
Age at kidney failure- stratified by variant type for a) Alport Syndrome and b) Heterozygous genotypes*

No significant differences were observed when stratifying by variant type within heterozygous genotypes (Figure 2b). Comparing individuals with biallelic *COL4A3/4* protein length-altering variants and male XLAS with protein length-altering variants, no difference in kidney survival was observed (p=0.78), suggesting that the younger age at KF observed with ARAS versus male XLAS may be explained by differing proportions of protein length-altering variants in each group (Supplementary Figure 6). Amongst *COL4A3/4* heterozygotes, missense variants were significantly over-represented relative to protein length-altering variants when compared with the variant distribution observed in ARAS (Supplementary Table 5, p=0.001).

No statistically significant differences were observed when stratifying by molecular characteristics (Supplementary Figure 7).

### Distribution of patients across CKD stages and proteinuria levels

Supplementary figures 8 and 9 include both genetically and clinically diagnosed patients. For patients with Alport Syndrome genotypes there were a limited number of patients with moderate kidney impairment able to contribute data: most patients <20 years old were in CKD stage 1, however by 24 years, the greatest proportion of patients had reached KF, with <28% of patients in CKD stages 2-5 at any age. Progression to a UPCR of ≥1.0g/g was similarly rapid.

### eGFR slope

Within the genetically confirmed cohort, eGFR slope varied by genotype (Figure 3), but accelerated between CKD stages 1 to 4 across all genotypes (p-value <0.0001 for difference between CKD stages for all groups). For instance, in male XLAS, eGFR slope was -1.4 ml/min/1.73m^2^/year (95% CI -2.0 to -0.8) in CKD stage 1 and -6.4 (-7.0 to -5.8) in stage 4, making the average time to progress from eGFR 30ml/min/1.73m^2^ to KF only 2.4 years (Supplementary Table 6). Results for clinically diagnosed patients showed similar results (p-value <0.0001, Supplementary Figure 10), as did sensitivity analyses using eGFR calculated using the CKD-EPI and bedside Schwartz equations (Supplementary Figure 11).

**Figure 3:**
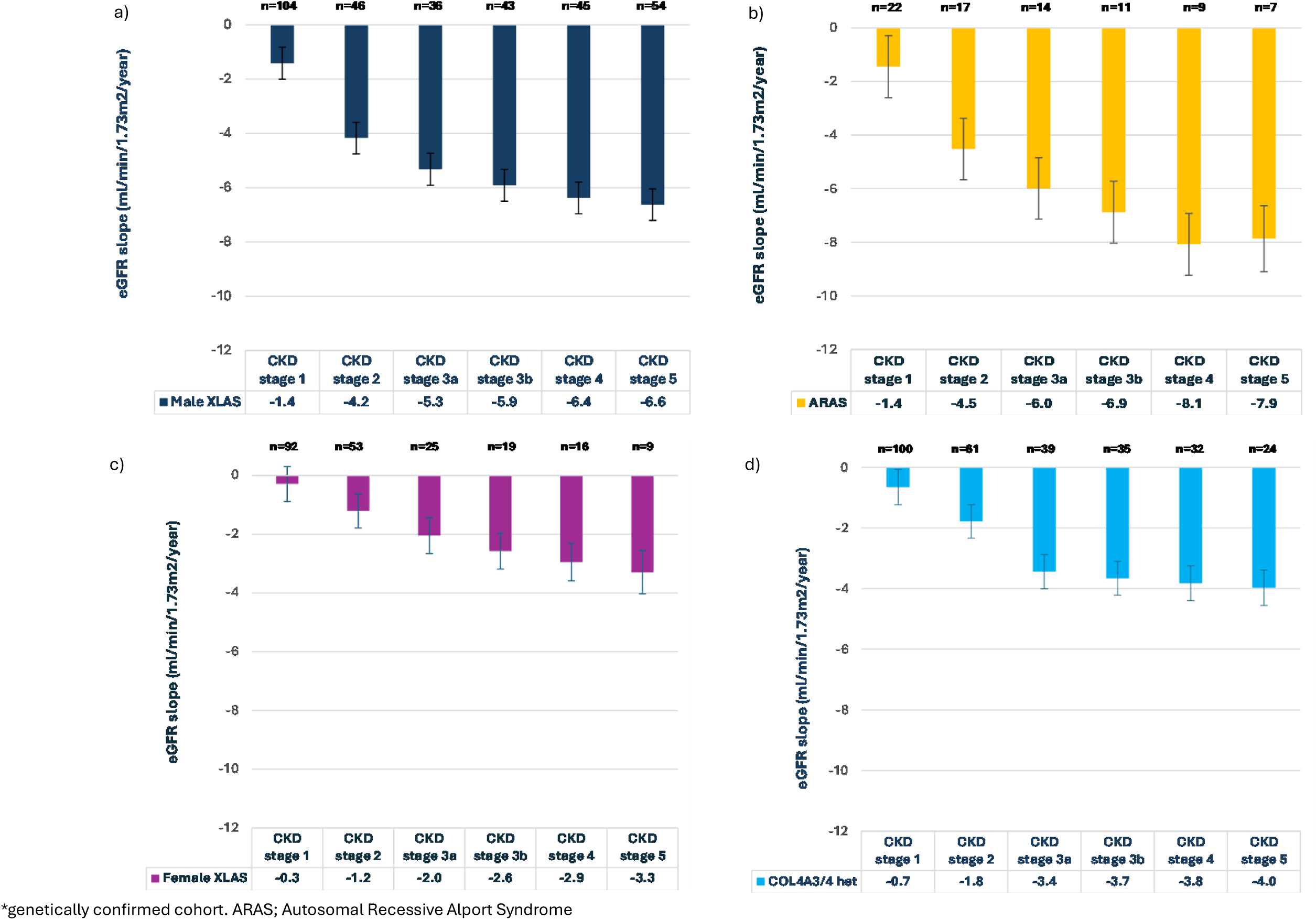
Annualised eGFR slope by CKD stage, stratified by genotype*

### Proteinuria progression and kidney outcomes

Median TAP prior to KRT initiation increased with CKD stage and was highest in those with Alport Syndrome genotypes (Table 3).

**Table 3:**
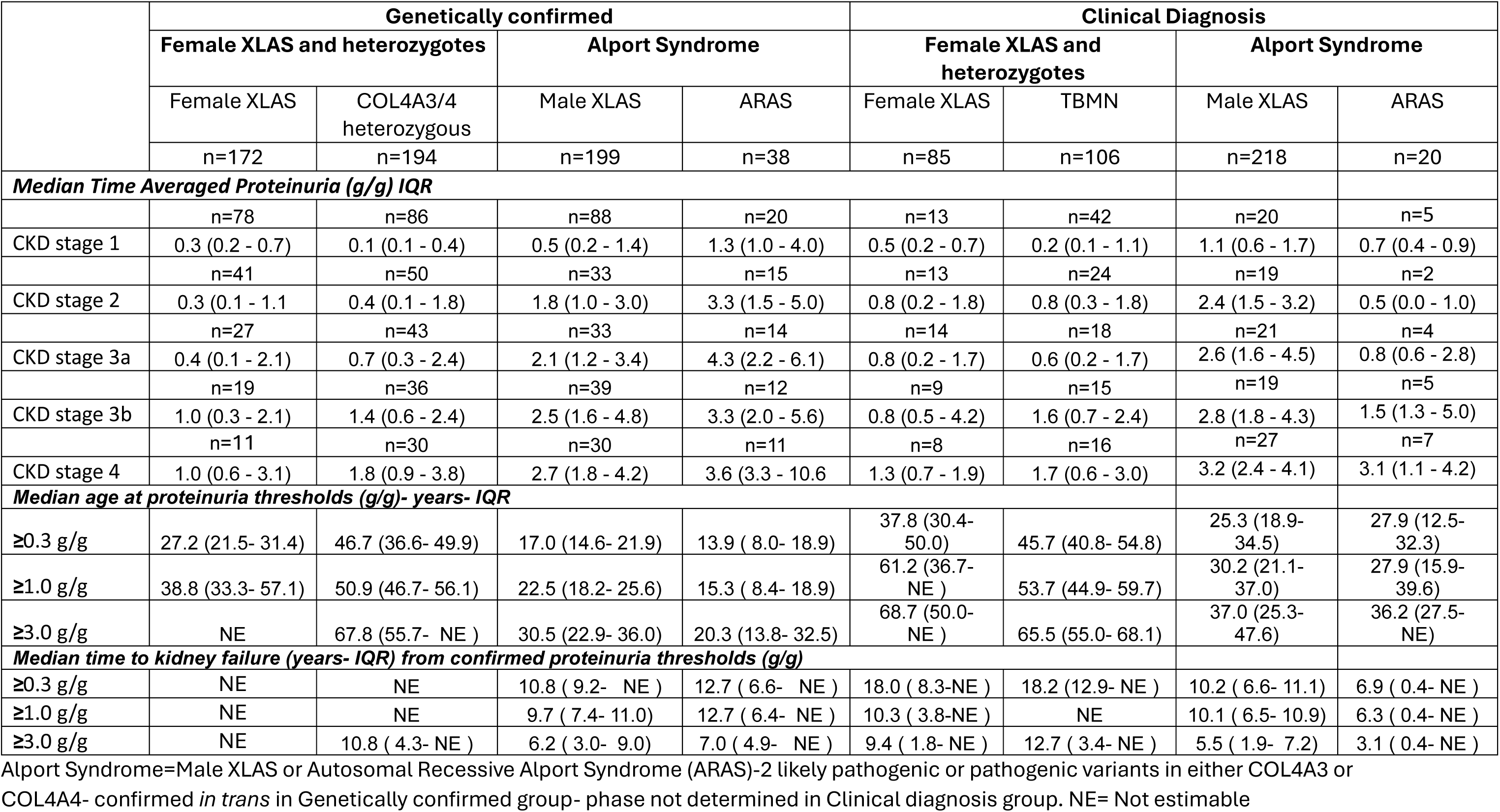
Proteinuria characteristics- stratified by genotype.

|  | Genetically confirmed |  |  |  | Clinical Diagnosis |  |  |  |
| --- | --- | --- | --- | --- | --- | --- | --- | --- |
|  | Female XLAS and heterozygotes |  | Alport Syndrome |  | Female XLAS and heterozygotes |  | Alport Syndrome |  |
|  | Female XLAS | COL4A3/4 heterozygous | Male XLAS | ARAS | Female XLAS | TBMN | Male XLAS | ARAS |
|  | n=172 | n=194 | n=199 | n=38 | n=85 | n=106 | n=218 | n=20 |
| **Median Time Averaged Proteinuria (g/g) IQR** |  |  |  |  |  |  |  |  |
|  | n=78 | n=86 | n=88 | n=20 | n=13 | n=42 | n=20 | n=5 |
| CKD stage 1 | 0.3 (0.2 - 0.7) | 0.1 (0.1 - 0.4) | 0.5 (0.2 - 1.4) | 1.3 (1.0 - 4.0) | 0.5 (0.2 - 0.7) | 0.2 (0.1 - 1.1) | 1.1 (0.6 - 1.7) | 0.7 (0.4 - 0.9) |
|  | n=41 | n=50 | n=33 | n=15 | n=13 | n=24 | n=19 | n=2 |
| CKD stage 2 | 0.3 (0.1 - 1.1) | 0.4 (0.1 - 1.8) | 1.8 (1.0 - 3.0) | 3.3 (1.5 - 5.0) | 0.8 (0.2 - 1.8) | 0.8 (0.3 - 1.8) | 2.4 (1.5 - 3.2) | 0.5 (0.0 - 1.0) |
|  | n=27 | n=43 | n=33 | n=14 | n=14 | n=18 | n=21 | n=4 |
| CKD stage 3a | 0.4 (0.1 - 2.1) | 0.7 (0.3 - 2.4) | 2.1 (1.2 - 3.4) | 4.3 (2.2 - 6.1) | 0.8 (0.2 - 1.7) | 0.6 (0.2 - 1.7) | 2.6 (1.6 - 4.5) | 0.8 (0.6 - 2.8) |
|  | n=19 | n=36 | n=39 | n=12 | n=9 | n=15 | n=19 | n=5 |
| CKD stage 3b | 1.0 (0.3 - 2.1) | 1.4 (0.6 - 2.4) | 2.5 (1.6 - 4.8) | 3.3 (2.0 - 5.6) | 0.8 (0.5 - 4.2) | 1.6 (0.7 - 2.4) | 2.8 (1.8 - 4.3) | 1.5 (1.3 - 5.0) |
|  | n=11 | n=30 | n=30 | n=11 | n=8 | n=16 | n=27 | n=7 |
| CKD stage 4 | 1.0 (0.6 - 3.1) | 1.8 (0.9 - 3.8) | 2.7 (1.8 - 4.2) | 3.6 (3.3 - 10.6) | 1.3 (0.7 - 1.9) | 1.7 (0.6 - 3.0) | 3.2 (2.4 - 4.1) | 3.1 (1.1 - 4.2) |
| **Median age at proteinuria thresholds (g/g)- years- IQR** |  |  |  |  |  |  |  |  |
| ≥0.3 g/g | 27.2 (21.5- 31.4) | 46.7 (36.6- 49.9) | 17.0 (14.6- 21.9) | 13.9 ( 8.0- 18.9) | 37.8 (30.4- 50.0) | 45.7 (40.8- 54.8) | 25.3 (18.9- 34.5) | 27.9 (12.5- 32.3) |
| ≥1.0 g/g | 38.8 (33.3- 57.1) | 50.9 (46.7- 56.1) | 22.5 (18.2- 25.6) | 15.3 ( 8.4- 18.9) | 61.2 (36.7- NE ) | 53.7 (44.9- 59.7) | 30.2 (21.1- 37.0) | 27.9 (15.9- 39.6) |
| ≥3.0 g/g | NE | 67.8 (55.7- NE ) | 30.5 (22.9- 36.0) | 20.3 (13.8- 32.5) | 68.7 (50.0- NE ) | 65.5 (55.0- 68.1) | 37.0 (25.3- 47.6) | 36.2 (27.5- NE) |
| **Median time to kidney failure (years- IQR) from confirmed proteinuria thresholds (g/g)** |  |  |  |  |  |  |  |  |
| ≥0.3 g/g | NE | NE | 10.8 ( 9.2- NE ) | 12.7 ( 6.6- NE ) | 18.0 ( 8.3-NE ) | 18.2 (12.9- NE ) | 10.2 ( 6.6- 11.1) | 6.9 ( 0.4- NE ) |
| ≥1.0 g/g | NE | NE | 9.7 ( 7.4- 11.0) | 12.7 ( 6.4- NE ) | 10.3 ( 3.8-NE ) | NE | 10.1 ( 6.5- 10.9) | 6.3 ( 0.4- NE ) |
| ≥3.0 g/g | NE | 10.8 ( 4.3- NE ) | 6.2 ( 3.0- 9.0) | 7.0 ( 4.9- NE ) | 9.4 ( 1.8- NE ) | 12.7 ( 3.4- NE ) | 5.5 ( 1.9- 7.2) | 3.1 ( 0.4- NE ) |

Alport Syndrome=Male XLAS or Autosomal Recessive Alport Syndrome (ARAS)-2 likely pathogenic or pathogenic variants in either COL4A3 or COL4A4- confirmed *in trans* in Genetically confirmed group- phase not determined in Clinical diagnosis group. NE= Not estimable

Within the genetically confirmed cohort, a linear mixed model of proteinuria by age showed genotype-specific differences: proteinuria increased significantly in males with XLAS (7.4% increase/year, 95% CI (5.5-9.3%), p<0.0001) and ARAS (6.4% increase/year (2.9-10.0%), p<0.0003), but not in heterozygous genotypes (*COL4A3/4* heterozygotes 1.2% increase/year (- 0.3 to 2.8), p=0.13, Female XLAS 0.3% increase/year (-1.5 to 2.1), p=0.75) (Supplementary Figure 12).

However, when modelled using non-linear splines, proteinuria progression appeared bimodal for female XLAS and female *COL4A3/4* heterozygotes, with peaks at 20-30 years, and again after 50 years (Supplementary Figure 13).

In the genetically confirmed cohort, proteinuria thresholds were reached at a younger age by patients with Alport syndrome compared to heterozygous genotypes (Supplementary Figure 14a): 25% of patients with Alport Syndrome genotypes reached ≥3.0g/g by 17 years (IQR 15.0-21), versus 48 years (32.9-54.9) in heterozygous genotypes (Supplementary Figure 14b). Within 5-years after reaching proteinuria thresholds of ≥1g/g and ≥3g/g, there were no significant differences in kidney survival between genotypes (log-rank p=0.14 and p=0.17, respectively, Supplementary Figure 15). Survival curves diverged beyond 5 years, and over total follow-up those with Alport Syndrome genotypes had a significantly shorter time to KF (Figure 4), although the magnitude of these differences, especially from ≥3.0g/g, were small (25% time to KF: Alport Syndrome 2.8 years (1.5-4.9), Heterozygous 5.2 (1.8-10.8), p=0.009). Similar findings were observed in the clinically diagnosed cohort (Supplementary Figures 16-18). Sensitivity analyses, with requirements on subsequent values, showed similar results, with much smaller differences in kidney survival between genotypes once a ≥3.0g/g threshold had been exceeded (Supplementary Figure 15 and 18).

**Figure 4:**
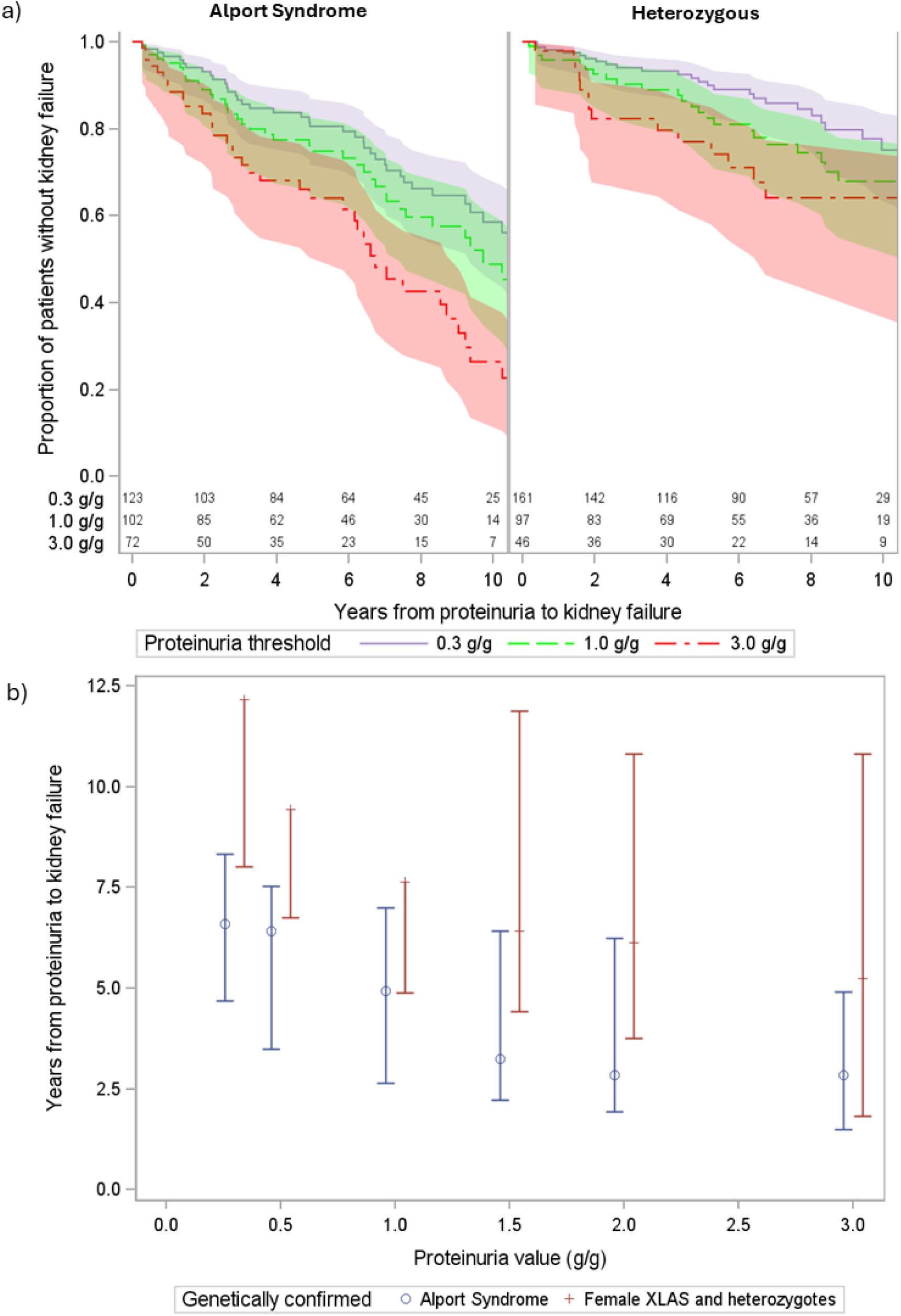
a) Kaplan Meier of time from reaching proteinuria values to kidney failure, stratified by Alport Syndrome and heterozygous genotypes b) 25^th^ centile time to kidney failure from exceeding proteinuria thresholds, for genetically confirmed cohort

No patients with Alport Syndrome genotypes who had not reached a proteinuria threshold of ≥0.3 g/g, and only one patient in the heterozygous group, progressed to KF (Supplementary Figure 19).

### Associations between eGFR, proteinuria and kidney outcomes

Across all AS patients (n=1032), median UPCR in the year prior to an eGFR 90, 60 and 45ml/min/1.73m^2^ were 1.59, 2.11 and 2.61g/g, respectively. Below-median UPCR was associated with a longer time to KF at all eGFR thresholds (eGFR 90 p=0.018, eGFR 60 p=0.012, eGFR 45 ml/min/1.73m^2^ p=0.0001, Figure 5). For instance, median time to KF from an eGFR 45ml/min/1.73m^2^ was 3.0 vs 6.5 years for those with above vs below median UPCR respectively. These analyses were also performed stratified by Alport Syndrome and heterozygous genotypes (Supplementary Figure 20). Median proteinuria levels in the year prior to each eGFR threshold were higher in those with Alport Syndrome vs heterozygous genotypes (eGFR 90ml/min/1.73m^2^: median UPCR 2.22 vs 0.87 g/g, eGFR 60: 2.98 vs 1.58g/g, eGFR 45: 3.85 vs 1.89g/g). Above-median UPCR was associated with shorter times to KF for both genotype categories from eGFR 45ml/min/1.73m^2^ (Alport Syndrome p=0.033, Heterozygous p=0.022).

**Figure 5:**
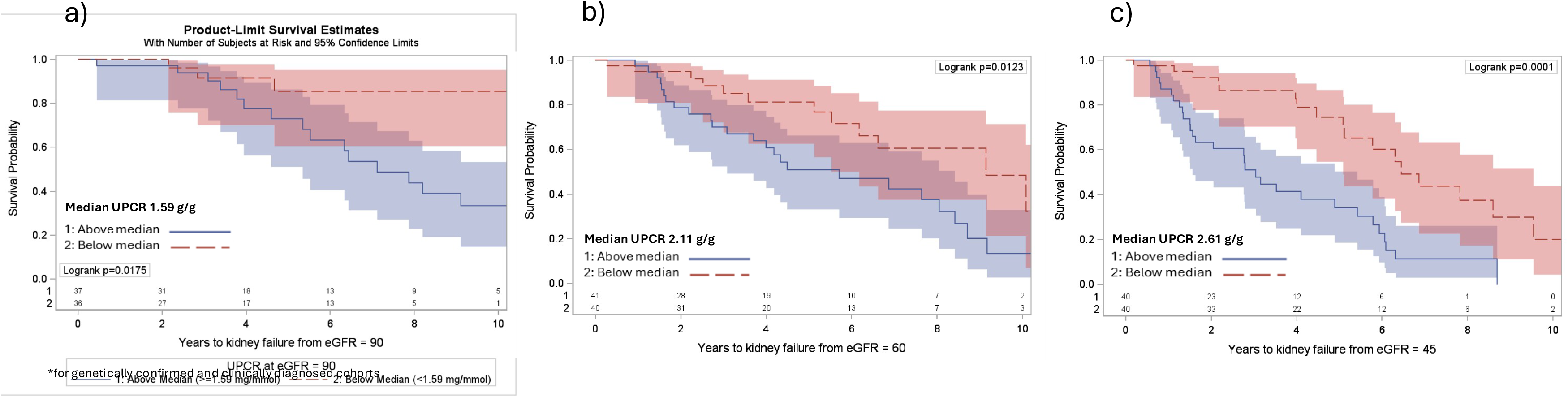
Time to KF from a) eGFR 90 b) eGFR 60 c) eGFR 45 ml/min/1.73m^2^, stratified by median proteinuria in year prior to each eGFR threshold for whole cohort*

## Discussion

In this large national cohort of individuals with AS, we identified consistent and clinically relevant relationships between genotype, proteinuria, and progression to KF. Across all genotypes, decline in kidney function was non-linear and accelerated with advancing CKD stage. Although genotype influenced the age at onset and early disease trajectory, proteinuria was strongly associated with subsequent risk of KF across a range of eGFR thresholds. Notably, once comparable levels of proteinuria were reached, short- to medium-term kidney outcomes were similar across genotype groups.

These findings have implications for both clinical care and clinical trial design. First, they suggest that proteinuria reflects a common downstream pathway of glomerular injury that integrates the effects of genotype and disease duration. While genotype remains important in determining lifetime risk and age at which outcomes occur, the attenuation of between-genotype group outcome differences at similar levels of proteinuria suggests that, beyond a threshold of established glomerular injury, prognosis may be more closely linked to current disease activity than to underlying genotype. For approximately 5-years after reaching proteinuria thresholds, kidney survival did not differ between Alport syndrome and heterozygous genotypes from ≥1.0g/g and ≥3.0g/g thresholds. Once proteinuria exceeded ≥3.0g/g, kidney survival did not differ between genotype groups over 5-years, and differences over total follow-up were modest (<2.5 years across sensitivity analyses). Sensitivity analyses using more stringent proteinuria definitions yielded similar results to those based on single values, supporting the robustness of these observations and approach. This has practical implications for risk-stratification and patient counselling, particularly in individuals with heterozygous genotypes who may otherwise be considered at relatively low risk.

Second, the strong and consistent association between proteinuria and KF across eGFR levels supports its use as a disease-relevant prognostic marker in AS. Nearly all participants developed proteinuria ≥0.3 g/g prior to KF, and higher levels were associated with substantially shorter time to KF at comparable levels of kidney function. These findings are concordant with observations in other glomerular diseases, including IgAN^16^ and C3 glomerulopathy^17^, in which proteinuria captures risk of progression and reflects treatment response^27^. Although our analyses do not, on their own, establish proteinuria as a validated surrogate endpoint for KF, they provide disease-specific evidence that changes in proteinuria are closely linked to clinically meaningful outcomes in AS and may therefore be informative for the design and interpretation of interventional trials. The near-universal presence of significant proteinuria before KF across genotypes provides biological plausibility for this relationship, and confirmation could have immediate implications for trial design and regulatory evaluation in AS, potentially accelerating access to new therapies for patients with this progressive, life-limiting rare disease.

Proteinuria trajectories varied by genotype and disease stage. Among female heterozygotes, trajectories appeared bimodal, with a peak at 20–30 years that may reflect pregnancy-associated exacerbation. Over the long-term however, females with XLAS demonstrated no sustained increase in proteinuria.

We also observed that eGFR decline accelerated markedly with advancing CKD stage across all genotypes. This was most marked in ARAS, where annualised eGFR slope increased from -1.4 ml/min/1.73m^2^/year in CKD stage 1 to -8.1 ml/min/1.73m^2^/year in stage 4. This degree of non-linearity exceeded that reported in general CKD populations^37,38^; in large Swedish and US CKD cohorts, difference in 3-year eGFR slope between patients with eGFR >60 and <60ml/min/1.73m^2^ was <2ml/min/1.73m^2 39^. These findings may haveimportant implications for the use of eGFR slope as a trial endpoint. Differences in baseline eGFR between treatment groups may substantially influence observed slopes, and short-term changes in slope may not be comparable across different disease stages. These findings underscore the importance of careful cohort selection and baseline adjustment when interpreting eGFR-based endpoints in AS and may explain clinical observations of rapid deterioration late in the disease and ‘crash-landing’ –i.e. requiring KRT sooner than anticipated.

Consistent with prior studies ^5,9,40^, individuals with ARAS and male XLAS experienced earlier onset of KF and more severe disease trajectories, particularly in the presence of protein length–altering variants. Among these groups, those with stop-gain variants reached KF approximately 10 years earlier than other truncating variants, highlighting the importance of both genotype and variant type. In contrast, differences by variant type were less apparent in heterozygous genotypes, likely reflecting biological heterogeneity and limited statistical power (owing to lower overall event-rate).

Pathogenic or likely pathogenic heterozygous *COL4A3/4* variants occur in ∼1% of the general population^41^ and individuals with these genotypes recruited to RaDaR represent a clinically enriched subset with more severe disease. In this cohort referred for specialist kidney care, average time to KF once reaching an eGFR of 30 ml/min/1.73m^2^ was only 3.9 years. While this does not alter estimated lifetime KF risk for all individuals with these genotypes, it suggests that heterozygous individuals who reach CKD stage 4 should not necessarily be regarded as “slow progressors” thereafter. The overrepresentation of missense variants in this group raises the possibility that these variants are more likely to be associated with clinically manifest disease, potentially through dominant-negative or gain-of-function effects –e.g. by reducing trimer secretion or activating intracellular stress responses that promote podocyte loss^42^. An alternative explanation is reduced ascertainment of missense variants in those with biallelic disease, although this seems unlikely given the high KF rate observed in ARAS across variant types.

Among patients with Gly substitution missense variants, we did not observe previously described differences in outcomes by other molecular characteristics^9^, however subgroup sizes were small.

This study has several strengths, including its large sample size for a rare disease, long duration of follow-up, and use of a national registry with linkage to KRT data. Longitudinal measurements enabled evaluation of proteinuria and eGFR trajectories, rather than reliance on single baseline measurements and were consistent across multiple sensitivity analyses. Where numbers allowed, findings are presented for both genetically confirmed and clinically diagnosed cohorts. Importantly, associations between eGFR decline, proteinuria, and KF remained significant within the clinically diagnosed cohort. If these findings contribute to the consideration of eGFR slope or proteinuria reduction as surrogate endpoints in AS, they may also support including patients with non-genetically confirmed diagnoses in future studies, avoiding excluding a generation of patients from trials of potentially transformative therapies due to the era of their diagnosis.

## Limitations

Several limitations should be considered. First, this was an observational study, and although the relationship between outcome and proteinuria was strong, we do not provide evidence (or claim) that proteinuria itself causes kidney damage– it may simply reflect cumulative glomerular injury burden. Proteinuria may reflect underlying disease severity as well as treatment effects, and whilst medication data were incomplete so we could not account fully for time-varying therapies, including renin–angiotensin system blockade and SGLT2 inhibitor use, for those with data available, ACE-inhibitor or Angiotensin Receptor Blocker use was comparable with other retrospective studies in AS^2^ (Supplementary Table 3). Second, approximately half of participants lacked a genetically confirmed diagnosis, introducing potential misclassification, although results were consistent in clinically diagnosed individuals. Third, as a UK-based cohort, results may not be fully generalizable to other populations. Finally, missing data and variability in measurement frequency inherent to real-world datasets may have influenced estimates of proteinuria and eGFR trajectories. Despite the large cohort, limited numbers of patients were able to contribute data at specific time-points, due partly to the small proportion of patients at any age in CKD stages 2-5. This reinforces the need for multi-registry collaborative analyses to identify sufficient patients to validate results (previously accomplished in IgAN and INS^15,43^), emphasises the narrow recruitment window for clinical trials, and underscores challenges in using eGFR slope as a surrogate end-point.

## Conclusion

In this large cohort of individuals with AS, proteinuria was strongly associated with progression to KF across genotypes and levels of kidney function, and differences in outcomes by genotype were attenuated once comparable levels of proteinuria were reached. These findings identify proteinuria as a key prognostic marker in AS and could help inform its role in clinical risk stratification and its potential utility as a surrogate endpoint for KF in future AS trials.

## Author contributions

Concept and design: DPG, ANT Acquisition, analysis, or interpretation of data: KW, DP, SM, KT, AB, DPG, ANT Drafting of the manuscript: KW, DPG, DP Critical review of the manuscript for important intellectual content: DPG, KW, DP, SM, KT, AB, TO, SG, HR, KF, E I-E, KA, SL, PD, JL, AM, BH, RL, ANT Obtained funding: DPG Administrative, technical, or material support: AB Supervision: DPG

## Data sharing statement

The RaDaR database is hosted by the UK Renal Registry and its metadata are available via https://rarerenal.org. Individual-level data are not available for export. Proposals to perform analyses using the data for academic, audit, or commercial purposes can be made to the RaDaR Operations Group via https://rarerenal.org.

## References

1. Mabillard H, Ryan R, Tzoumas N, Gear S, Sayer JA: Explaining Alport syndrome-lessons from the adult nephrology clinic. Journal of rare diseases (Berlin, Germany) [Internet] 3: 2024 Available from: https://pubmed.ncbi.nlm.nih.gov/38745975/ [cited 2026 Feb 7]

2. Yamamura T, Horinouchi T, Nagano C, Omori T, Sakakibara N, Aoto Y, et al.: Genotype-phenotype correlations influence the response to angiotensin-targeting drugs in Japanese patients with male X-linked Alport syndrome. Kidney Int. 98: 1605–1614, 2020

3. Gross O, Netzer KO, Lambrecht R, Seibold S, Weber M: Meta-analysis of genotype-phenotype correlation in X-linked Alport syndrome: Impact on clinical counselling. Nephrology Dialysis Transplantation [Internet] 17: 1218–1227, 2002 Available from: https://pubmed.ncbi.nlm.nih.gov/12105244/ [cited 2025 May 12]

4. Bekheirnia MR, Reed B, Gregory MC, McFann K, Shamshirsaz AA, Masoumi A, et al.: Genotype-phenotype correlation in X-linked Alport syndrome. Journal of the American Society of Nephrology [Internet] 21: 876–883, 2010 Available from: https://pubmed.ncbi.nlm.nih.gov/20378821/ [cited 2025 May 12]

5. Yamamura T, Horinouchi T, Adachi T, Terakawa M, Takaoka Y, Omachi K, et al.: Development of an exon skipping therapy for X-linked Alport syndrome with truncating variants in COL4A5. Nature Communications 2020 11:1 [Internet] 11: 1–8, 2020 Available from: https://www.nature.com/articles/s41467-020-16605-x [cited 2025 May 12]

6. Riedhammer KM, Simmendinger H, Tasic V, Putnik J, Abazi-Emini N, Stajic N, et al.: Is there a dominant-negative effect in individuals with heterozygous disease-causing variants in COL4A3/COL4A4? Clin. Genet. [Internet] 105: 406–414, 2024 Available from: /doi/pdf/10.1111/cge.14471 [cited 2025 May 13]

7. Storey H, Savige J, Sivakumar V, Abbs S, Flinter FA: COL4A3/COL4A4 mutations and features in individuals with autosomal recessive alport syndrome. Journal of the American Society of Nephrology [Internet] 24: 1945–1954, 2013 Available from: https://journals.lww.com/jasn/fulltext/2013/12000/col4a3_col4a4_mutations_and_features_in.7.aspx [cited 2025 May 13]

8. Savige J: Heterozygous Pathogenic COL4A3 and COL4A4 Variants (Autosomal Dominant Alport Syndrome) Are Common, and Not Typically Associated With End-Stage Kidney Failure, Hearing Loss, or Ocular Abnormalities. Kidney Int. Rep. [Internet] 7: 1933, 2022 Available from: https://pmc.ncbi.nlm.nih.gov/articles/PMC9458992/ [cited 2025 May 12]

9. Gibson JT, Huang M, Shenelli Croos Dabrera M, Shukla K, Rothe H, Hilbert P, et al.: Genotype–phenotype correlations for COL4A3–COL4A5 variants resulting in Gly substitutions in Alport syndrome. Sci. Rep. [Internet] 12: 2722, 2022 Available from: https://pmc.ncbi.nlm.nih.gov/articles/PMC8854626/ [cited 2025 May 13]

10. Gross O, Rheault MN, Simon J, Knebelmann B, Shen Y, Zhang Q, et al.: Prospective Cohort Study in Alport Syndrome Patients Under Standard Therapy. Kidney Int. Rep. [Internet] 10: 1360–1371, 2025 Available from: https://www.kireports.org/action/showFullText?pii=S2468024925001263 [cited 2025 May 12]

11. Langsford D, Tang M, Djurdjev O, Er L, Levin A: The Variability of Estimated Glomerular Filtration Rate Decline in Alport Syndrome. Can. J. Kidney Health Dis. [Internet] 3: 2054358116679129, 2016 Available from: https://pmc.ncbi.nlm.nih.gov/articles/PMC5518963/ [cited 2025 May 12]

12. Gale DP, Gross O, Wang F, Esteban De La Rosa RJ, Hall M, Sayer JA, et al.: A Randomized Controlled Clinical Trial Testing Effects of Lademirsen on Kidney Function Decline in Adults with Alport Syndrome. Clinical Journal of the American Society of Nephrology [Internet] 19: 995–1004, 2024 Available from: https://journals.lww.com/cjasn/fulltext/2024/08000/a_randomized_controlled_clinical_trial_testing.11.aspx [cited 2025 Oct 2]

13. Gibson JT, de Gooyer M, Huang M, Savige J: A Systematic Review of Pathogenic COL4A5 Variants and Proteinuria in Women and Girls With X-linked Alport Syndrome. Kidney Int. Rep. 7: 2454–2461, 2022

14. Pitcher D, Braddon F, Hendry B, Mercer A, Osmaston K, Saleem MA, et al.: Long-Term Outcomes in IgA Nephropathy. Clin. J. Am. Soc. Nephrol. [Internet] 18: 727–738, 2023 Available from: https://pubmed.ncbi.nlm.nih.gov/37055195/ [cited 2025 Oct 6]

15. Inker LA, Mondal H, Greene T, Masaschi T, Locatelli F, Schena FP, et al.: Early Change in Urine Protein as a Surrogate End Point in Studies of IgA Nephropathy: An Individual-Patient Meta-analysis. Am. J. Kidney Dis. [Internet] 68: 392–401, 2016 Available from: https://pubmed.ncbi.nlm.nih.gov/27032886/ [cited 2025 Mar 13]

16. Pitcher D, Braddon F, Hendry B, Mercer A, Barratt J, Steenkamp R, et al.: Long-Term Outcomes in Nephrotic Syndrome by Kidney Biopsy Diagnosis and Proteinuria. Journal of the American Society of Nephrology [Internet] 36: 1398–1413, 2025 Available from: https://journals.lww.com/jasn/fulltext/2025/07000/long_term_outcomes_in_nephrotic_syndrome_by_kidney.17.aspx [cited 2025 Nov 30]

17. Masoud S, Wong K, Pitcher D, Downward L, Proudfoot C, Webb NJA, et al.: Quantifying association of early proteinuria and estimated glomerular filtration rate changes with long-term kidney failure in C3 glomerulopathy and immune-complex membranoproliferative glomerulonephritis using the United Kingdom RaDaR Registry. Kidney Int. [Internet] 108: 2025 Available from: https://pubmed.ncbi.nlm.nih.gov/40582408/ [cited 2025 Oct 6]

18. Caravaca-Fontán F, Toledo-Rojas R, Huerta A, Pérez-Canga JL, Martínez-Miguel P, Miquel R, et al.: Comparative Analysis of Proteinuria and Longitudinal Outcomes in Immune Complex Membranoproliferative Glomerulonephritis and C3 Glomerulopathy. Kidney Int. Rep. [Internet] 0: 2025 Available from: https://www.kireports.org/action/showFullText?pii=S246802492500049X [cited 2025 Mar 13]

19. Gonçalves PL, de Faria VP, Furlano M, Almeida C, Sampani E, Teran MP, et al.: #2327 Extent of proteinuria in autosomal dominant Alport syndrome compared to X linked Alport syndrome. Nephrology Dialysis Transplantation [Internet] 39: 2024 Available from: 10.1093/ndt/gfae069.245 [cited 2025 Jul 10]

20. Zeng M, Di H, Ding J, Zhang Y, Xu H, Xie J, et al.: A Prediction Model of Disease Progression in X-Linked Alport syndrome Based on Clinical Characteristics and Genetic Variants. Kidney Int. Rep. [Internet] 10: 2024–2034, 2025 Available from: https://www.sciencedirect.com/science/article/pii/S2468024925001342 [cited 2025 Jul 10]

21. Gale DP, Gross O, Wang F, Esteban De La Rosa RJ, Hall M, Sayer JA, et al.: A Randomized Controlled Clinical Trial Testing Effects of Lademirsen on Kidney Function Decline in Adults with Alport Syndrome. Clinical Journal of the American Society of Nephrology [Internet] 19: 995–1004, 2024 Available from: https://journals.lww.com/cjasn/fulltext/2024/08000/a_randomized_controlled_clinical_trial_testing.11.aspx [cited 2025 Oct 6]

22. Boeckhaus J, Gale DP, Simon J, Ding J, Zhang Y, Bergmann C, et al.: SGLT2-Inhibition in Patients With Alport Syndrome. Kidney Int. Rep. [Internet] 9: 3490–3500, 2024 Available from: https://www.sciencedirect.com/science/article/pii/S2468024924019429 [cited 2025 Oct 7]

23. Kim SG, Inker LA, Packham DK, Ranganatha D, Rastogi A, Rheault MN, et al.: WCN23-1126 ATRASENTAN FOR THE TREATMENT OF IGA NEPHROPATHY: INTERIM RESULTS OF THE AFFINITY STUDY. Kidney Int. Rep. [Internet] 8: 1902, 2023 Available from: https://www.kireports.org/action/showFullText?pii=S246802492301207X [cited 2025 Oct 7]

24. Trachtman H, Coppo R, Masthan Ahmed NA, Lieberman K V., Mercer A, Rheault MN, et al.: Sparsentan (SPAR) in Pediatric Patients with Rare Proteinuric Kidney Disease: Preliminary Findings from the EPPIK Study. Journal of the American Society of Nephrology [Internet] 35: 2024 Available from: https://www.asn-online.org/education/kidneyweek/2024/program-abstract.aspx?controlId=4121802 [cited 2025 Dec 18]

25. Inker LA, Heerspink HJL, Tighiouart H, Levey AS, Coresh J, Gansevoort RT, et al.: GFR slope as a surrogate end point for kidney disease progression in clinical trials: A meta-analysis of treatment effects of randomized controlled trials. Journal of the American Society of Nephrology [Internet] 30: 1735–1745, 2019 Available from: https://pubmed.ncbi.nlm.nih.gov/31292197/ [cited 2025 Mar 13]

26. Inker LA, Heerspink HJL, Tighiouart H, Chaudhari J, Miao S, Diva U, et al.: Association of Treatment Effects on Early Change in Urine Protein and Treatment Effects on GFR Slope in IgA Nephropathy: An Individual Participant Meta-analysis. Am. J. Kidney Dis. [Internet] 78: 340–349.e1, 2021 Available from: https://pubmed.ncbi.nlm.nih.gov/33775708/ [cited 2025 Mar 13]

27. Thompson A, Carroll K, Inker LA, Floege J, Perkovic V, Boyer-Suavet S, et al.: Proteinuria Reduction as a Surrogate End Point in Trials of IgA Nephropathy. Clin. J. Am. Soc. Nephrol. [Internet] 14: 469–481, 2019 Available from: https://pubmed.ncbi.nlm.nih.gov/30635299/ [cited 2025 Mar 13]

28. Levey AS, Gansevoort RT, Coresh J, Inker LA, Heerspink HL, Grams ME, et al.: Change in Albuminuria and GFR as End Points for Clinical Trials in Early Stages of CKD: A Scientific Workshop Sponsored by the National Kidney Foundation in Collaboration With the US Food and Drug Administration and European Medicines Agency. Am. J. Kidney Dis. [Internet] 75: 84–104, 2020 Available from: https://pubmed.ncbi.nlm.nih.gov/31473020/ [cited 2025 Mar 13]

29. Heerspink HJL, Collier WH, Chaudhari J, Miao S, Tighiouart H, Appel GB, et al.: A meta-analysis of albuminuria as a surrogate endpoint for kidney failure. Nature Medicine 2025 [Internet] 1–7, 2025 Available from: https://www.nature.com/articles/s41591-025-04057-z [cited 2025 Nov 11]

30. Wong K, Pitcher D, Braddon F, Downward L, Steenkamp R, Annear N, et al.: Effects of rare kidney diseases on kidney failure: a longitudinal analysis of the UK National Registry of Rare Kidney Diseases (RaDaR) cohort. The Lancet [Internet] 403: 1279–1289, 2024 Available from: https://www.thelancet.com/action/showFullText?pii=S014067362302843X [cited 2025 Mar 13]

31. Wong K, Pitcher D, Braddon F, Downward L, Steenkamp R, Masoud S, et al.: Description and Cross-Sectional Analyses of 25,880 Adults and Children in the UK National Registry of Rare Kidney Diseases Cohort. Kidney Int. Rep. [Internet] 9: 2067–2083, 2024 Available from: http://www.kireports.org/article/S2468024924016991/fulltext [cited 2024 Jul 9]

32. Gale DP: From Data to Drug: The Translational Impact of RaDaR, the UK National Registry of Rare Kidney Diseases. Nephrol. Dial. Transplant [Internet] 2025 Available from: https://pubmed.ncbi.nlm.nih.gov/41137724/ [cited 2026 Mar 28]

33. Richards S, Aziz N, Bale S, Bick D, Das S, Gastier-Foster J, et al.: Standards and Guidelines for the Interpretation of Sequence Variants: A Joint Consensus Recommendation of the American College of Medical Genetics and Genomics and the Association for Molecular Pathology. Genet. Med. [Internet] 17: 405, 2015 Available from: https://pmc.ncbi.nlm.nih.gov/articles/PMC4544753/ [cited 2025 Oct 6]

34. Lennon R, Miner JH, Savige J, Weinstock BA, Gear S: Alport. Journal of the American Society of Nephrology [Internet] 2026 Available from: https://journals.lww.com/jasn/fulltext/9900/alportrenaming_an_extended_clinical_spectrum.968.aspx [cited 2026 Apr 24]

35. Levin A, Agarwal R, Herrington WG, Heerspink HL, Mann JFE, Shahinfar S, et al.: International consensus definitions of clinical trial outcomes for kidney failure: 2020. Kidney Int. [Internet] 98: 849–859, 2020 Available from: https://pubmed.ncbi.nlm.nih.gov/32998816/ [cited 2025 Mar 13]

36. Pottel H, Björk J, Courbebaisse M, Couzi L, Ebert N, Eriksen BO, et al.: Development and Validation of a Modified Full Age Spectrum Creatinine-Based Equation to Estimate Glomerular Filtration Rate. 10.7326/M20-4366 [Internet] 174: 183–191, 2020 Available from: /doi/pdf/10.7326/M20-4366?download=true [cited 2025 Oct 6]

37. Haynes R, Lewis D, Emberson J, Reith C, Agodoa L, Cass A, et al.: Effects of lowering LDL cholesterol on progression of kidney disease. Journal of the American Society of Nephrology [Internet] 25: 1825–1833, 2014 Available from: https://pmc.ncbi.nlm.nih.gov/articles/PMC4116066/ [cited 2025 Nov 25]

38. Li L, Astor BC, Lewis J, Hu B, Appel LJ, Lipkowitz MS, et al.: Longitudinal progression trajectory of GFR among patients with CKD. American Journal of Kidney Diseases [Internet] 59: 504–512, 2012 Available from: https://pubmed.ncbi.nlm.nih.gov/22284441/ [cited 2025 Nov 26]

39. Grams ME, Sang Y, Ballew SH, Matsushita K, Astor BC, Carrero JJ, et al.: Evaluating Glomerular Filtration Rate Slope as a Surrogate End Point for ESKD in Clinical Trials: An Individual Participant Meta-Analysis of Observational Data. J. Am. Soc. Nephrol. [Internet] 30: 1746–1755, 2019 Available from: https://pubmed.ncbi.nlm.nih.gov/31292199/ [cited 2025 Mar 13]

40. Savige J, Huang M, Croos Dabrera MS, Shukla K, Gibson J: Genotype-Phenotype Correlations for Pathogenic COL4A3–COL4A5 Variants in X-Linked, Autosomal Recessive, and Autosomal Dominant Alport Syndrome. Front. Med. (Lausanne). [Internet] 9: 2022 Available from: https://pubmed.ncbi.nlm.nih.gov/35602506/ [cited 2025 May 7]

41. Gibson J, Fieldhouse R, Chan MMY, Sadeghi-Alavijeh O, Burnett L, Izzi V, et al.: Prevalence Estimates of Predicted Pathogenic COL4A3-COL4A5 Variants in a Population Sequencing Database and Their Implications for Alport Syndrome. J. Am. Soc. Nephrol. [Internet] 32: 2273–2290, 2021 Available from: https://pubmed.ncbi.nlm.nih.gov/34400539/ [cited 2025 Nov 30]

42. Pieri M, Stefanou C, Zaravinos A, Erguler K, Stylianou K, Lapathitis G, et al.: Evidence for activation of the unfolded protein response in collagen iv nephropathies. Journal of the American Society of Nephrology [Internet] 25: 260–275, 2014 Available from: https://pubmed.ncbi.nlm.nih.gov/24262798/ [cited 2026 Mar 4]

43. Trachtman H, Inrig JK, Komers R: Sparsentan clinical trials in glomerular diseases: defining endpoints and a path forward in light of the parasol initiative. Future Rare Diseases [Internet] 5: 2461966, 2025 Available from: https://www.tandfonline.com/doi/pdf/10.1080/23995270.2025.2461966 [cited 2025 Oct 23]

