## Supplementary Materials for "Quantifying associations of genotype, proteinuria and eGFR with long-term kidney outcomes in Alport Syndrome using data from the UK National Registry of Rare Kidney Diseases (RaDaR)"

| Table of Contents | Page |
|---|---|
| Supplementary Methods | 2 |
| Supplementary Figure 1: Study inclusion diagram | 6 |
| Supplementary Table 1: Available data for each analysis | 7 |
| Supplementary Table 2: Additional demographic data | 8 |
| Supplementary Table 3 : Clinical demographics for genetically confirmed and clinically diagnosed cohorts | 9 |
| Supplementary Figure 2: Diagnosis year for genetically confirmed and clinically diagnosed subgroups, for a) Alport Syndrome and b) Heterozygous genotypes | 10 |
| Supplementary Table 4: Variant classification | 11 |
| Supplementary Figure 3: 25th centile Age at Kidney Failure, by genotype | 12 |
| Supplementary Figure 4: Age at kidney failure, stratified by variant type for a) Male X-Linked AS b) Autosomal Recessive Alport Syndrome c) Female X-Linked AS d) COL4A3/4 heterozygous variants | 13 |
| Supplementary Figure 5: Age at Kidney Failure for those with stop gain variants, other protein length altering and non-protein length altering variants, for a) Alport Syndrome b) Heterozygous Genotypes | 14 |
| Supplementary Figure 6: Kaplan Meier plots of age at KF by variant type, for patients with Male XLAS and 2 COL4A3 or 2 COL4A4 variants | 15 |
| Supplementary Table 5: Distribution of protein length altering and non-protein length altering variants by genotype | 16 |
| Supplementary Figure 7: Age at kidney failure, stratified by molecular characteristics a) Destabilising residue b) In or adjacent to Non-collagenous domain c) Exon position d) Exon position excluding last Non-collagenous domain for Alport Syndrome and Heterozygous genotypes | 17 |
| Supplementary Figure 8: Patient distribution across proteinuria levels (<0.5g/g, 0.5-1.0g/g, >1.0g/g) by age | 18 |
| Supplementary Figure 9: Patient distribution across CKD stages, by age | 19 |
| Supplementary Table 6: Annualised eGFR slope (ml/min/1.73m2/year), by genotype | 20 |
| Supplementary Figure 10: Annualised eGFR slope by CKD stage, for Alport Syndrome and Heterozygous genotypes for clinically diagnosed cohort | 21 |
| Supplementary Figure 11: Annualised eGFR slope by CKD stage, for Alport Syndrome and Heterozygous genotypes, using CKD-EPI 2021 or bedside Schwartz equations for a) genetically confirmed and b) clinically diagnosed cohorts | 22 |
| Supplementary Figure 12: Linear mixed model of proteinuria by age, stratified by genotype | 23 |
| Supplementary Figure 13: Non-linear model of proteinuria trajectory by age for a) Males and b) Females, stratified by genotype | 24 |
| Supplementary Figure 14: a) Cumulative incidence plot of age at proteinuria thresholds for all patients and b) stratified by Alport Syndrome and Heterozygous genotypes, for genetically confirmed cohort | 25 |
| Supplementary Figure 15: Kaplan Meier plots of time from reaching proteinuria thresholds of a) ≥0.3g/g b) ≥1.0 g/g c) ≥3.0 g/g to kidney failure , where date of reaching a proteinuria threshold was defined as a) date of first single value above that threshold, b) date of first value, and the first value >30 days later required to be above at least 50% of the threshold, and with no values within 30 days below 50% of the threshold c) b) date of first value, and the first value >30 days later required to be above at least 70% of the threshold, and with no values within 30 days below 70% of the threshold, for genetically confirmed cohort | 26 |
| Supplementary Figure 16: a) Cumulative plot of age at proteinuria thresholds for all patients and b) stratified by Alport Syndrome and Heterozygous genotypes, for clinically diagnosed cohort | 28 |
| Supplementary Figure 17: a) Kaplan Meier of time from proteinuria values to Kidney Failure, stratified by Alport Syndrome and heterozygous genotypes b) 25th centile time to kidney failure from exceeding proteinuria thresholds, by genotype for clinically diagnosed cohort | 29 |
| Supplementary Figure 18: Kaplan Meier plots of time from reaching proteinuria thresholds of a) ≥0.3g/g b) ≥1.0 g/g c) ≥3.0 g/g to kidney failure , where date of reaching a proteinuria threshold was defined as a) date of first single value above that threshold, b) date of first value, and the first value >30 days later required to be above at least 50% of the threshold, and with no values within 30 days below 50% of the threshold c) b) date of first value, and the first value >30 days later required to be above at least 70% of the threshold, and with no values within 30 days below 70% of the threshold, for clinically diagnosed cohort | 30 |
| Supplementary Figure 19: Time to kidney failure for patients with a confirmed proteinuria value >0.3g/g and those without, for patients with a) Alport Syndrome b) Heterozygous genotypes, for (i) genetically confirmed and (ii) clinically diagnosed cohorts | 32 |

| | |
|---|---|
| Supplementary Figure 20: Time to KF from a) eGFR 90 b) eGFR 60 c) eGFR 45 ml/min/1.73m2, stratified by median proteinuria in year prior to each eGFR threshold for i) Alport Syndrome and (ii) Heterozygous genotypes | 33 |
| National Registry of Rare Kidney Diseases (RaDaR) Consortium | 34 |
| Strengthening the Reporting of Observational Studies in Epidemiology (STROBE) checklist | 41 |



**Supplementary Methods**

*Data source and data linkages*

Data linkage with local hospitals and renal units enables retrospective and automated prospective collection of blood and urine results via the UK Renal Data Collaboration (UKRDC). Linkage with the UK Renal Registry (UKRR) provides validated data on kidney replacement therapy (KRT) initiation (including NHS Blood and Transplant data). RaDaR receives clinical genetic reports from NHS genomics hubs.

*Ethics*

RaDaR has ethical approval as a research registry provided by NHS South-West Central Bristol Research ethics committee (24/SW/0135 IRAS:349779) and the Scotland A ethics committee (24/SS/0092 IRAS:351286).

*Variable and outcome definitions*

| | |
|---|---|
| Date of diagnosis | Date of diagnosis recorded in RaDaR, date of genetic test or biopsy confirming a diagnosis of AS, whichever was earliest. |
| Follow-up time | Follow up time was classified as time from diagnosis to date of data extraction, or death. |
| Time-averaged proteinuria | The time-weighted averages for urinary protein-creatinine ratio (UPCR) used to define time-averaged proteinuria were calculated from the area under the curve of serial measurements, divided by length of follow-up, and stratified by CKD stage. |
| Sustained eGFR | A sustained eGFR below certain threshold was defined as two values at least 28 days apart, both below the threshold without an intervening value above the threshold. |

*Variant classification*

| | |
|---|---|
| Protein length-altering | Splice site, stop-gain/loss, frameshift, large insertions/deletions, exon deletion/duplications |
| Non-protein length altering | Missense glycine and non-glycine substitutions, small insertions/deletions <20 bp |

*Variant Annotation and ClinVar Classification*

Splice-site variants and other variant entries lacking standardised genomic notation, often provided in clinical genetic reports at the cDNA level without genomic coordinates or protein-level consequences, were first converted into valid HGVS genomic or transcript-based identifiers using the Ensembl Variant Effect Predictor (VEP) with HGNC gene models. This step enabled accurate mapping of the reported cDNA changes to their corresponding genomic positions and determination of variant type, such as truncating, missense or splice-site. The variants were then further annotated using VEP, supplemented with a custom query to both ClinVar and the LOVD Alport databases, to retrieve existing clinical classifications for *COL4A3/4* variants, allowing each variant to be confirmed as pathogenic, likely pathogenic, benign, likely benign, or of uncertain significance (VUS).

*Missense glycine substitutions*

Missense glycine substitutions were classified by 1) being adjacent to a non-collagenous (NC1) region 2) Exon position (1-20 or 21-carboxyl terminus, or 1-20 or 21 to last non-collagenous (NC1) region) 3) degree of predicted instability caused by the replacing residue (mildly destabilising: Ala, Ser, Cys; highly destabilising: Arg, Val, Glu, Asp, Trp).

*Methodology for proteinuria thresholds*

For analyses presented in the main text, date at reaching a certain proteinuria threshold was defined as the date of the first value above that chosen threshold, to reflect data available to clinicians in practice. Sensitivity analyses where date at reaching a proteinuria threshold was defined as date of the first value above a certain threshold were performed with a) the first value >30 days later required to be above at least 50% of the threshold, and with no values within 30 days below 50% of the threshold and b) using a 70% of threshold cut-off.

*eGFR slope and proteinuria trajectory calculations*

eGFR slope was estimated using mixed-effects regression models with a random intercept and slope for each patient. Interaction terms between slope, CKD stage and mutation type were included in the models to allow slope within each CKD stage to be estimated for each group. Sensitivity analyses using the CKD-EPI 2021, or the bedside Schwartz for those aged <16 years, instead of the European Kidney Function Consortium eGFR equation are presented in Supplementary Figure 11.

The relationship between log-transformed proteinuria and age was modelled using a mixed-effects regression model to account for repeated measures within individuals, firstly with the assumption that the relationship was linear, and then, to account for non-linearity, estimating age effects using a cubic spline with knots at the $25^{th}$, $50^{th}$ and $75^{th}$ percentiles of the age distribution.

*Methodology for stacked plots*

To demonstrate how many patients at each age have data available within RaDaR, stacked plots showing which ‘proteinuria state’ a patient is in at each year of age are presented in Supplementary Figure 8. Prior to a patient’s earliest proteinuria observation recorded in RaDaR a patient would be defined as in the “Pre-KRT No PCR” group. From the age at which the patient has their first proteinuria value they would then fall into one of the “Pre-KRT <0.5 g/g”, “Pre-KRT 0.5-1.0g/g” or “Pre-KRT > 1.0 g/g” groups depending on the value of the proteinuria observation. A patient can then progress ‘up’ through the groups if their proteinuria values increase, but they will not go back ‘down’ to a lower proteinuria group if their proteinuria was to fall. Once a patient initiates KRT they move into the “Post KRT” group and will remain in this group until their current age or death. To create a plot where the number of patients remains consistent throughout every age, and also to enable visualisation of the proportion of all patients able to contribute data across the entire age range, there is a group for patients who are alive but not yet reached that age; this group is omitted from some of the stacked plots meaning that patients only contribute data up to their current age. Once a patient dies they will be included in the “Deceased” group for the remainder of the ages. The same methodology was applied to sustained eGFR results to determine which CKD stage a patient was in at each year of age (Supplementary Figure 9).

Supplementary References

1. Levin A, Agarwal R, Herrington WG, Heerspink HL, Mann JFE, Shahinfar S, et al.: International consensus definitions of clinical trial outcomes for kidney failure: 2020. *Kidney Int.* [Internet] 98: 849–859, 2020 Available from: https://pubmed.ncbi.nlm.nih.gov/32998816/ [cited 2025 Mar 13]

**Supplementary Figure 1: Study inclusion diagram**

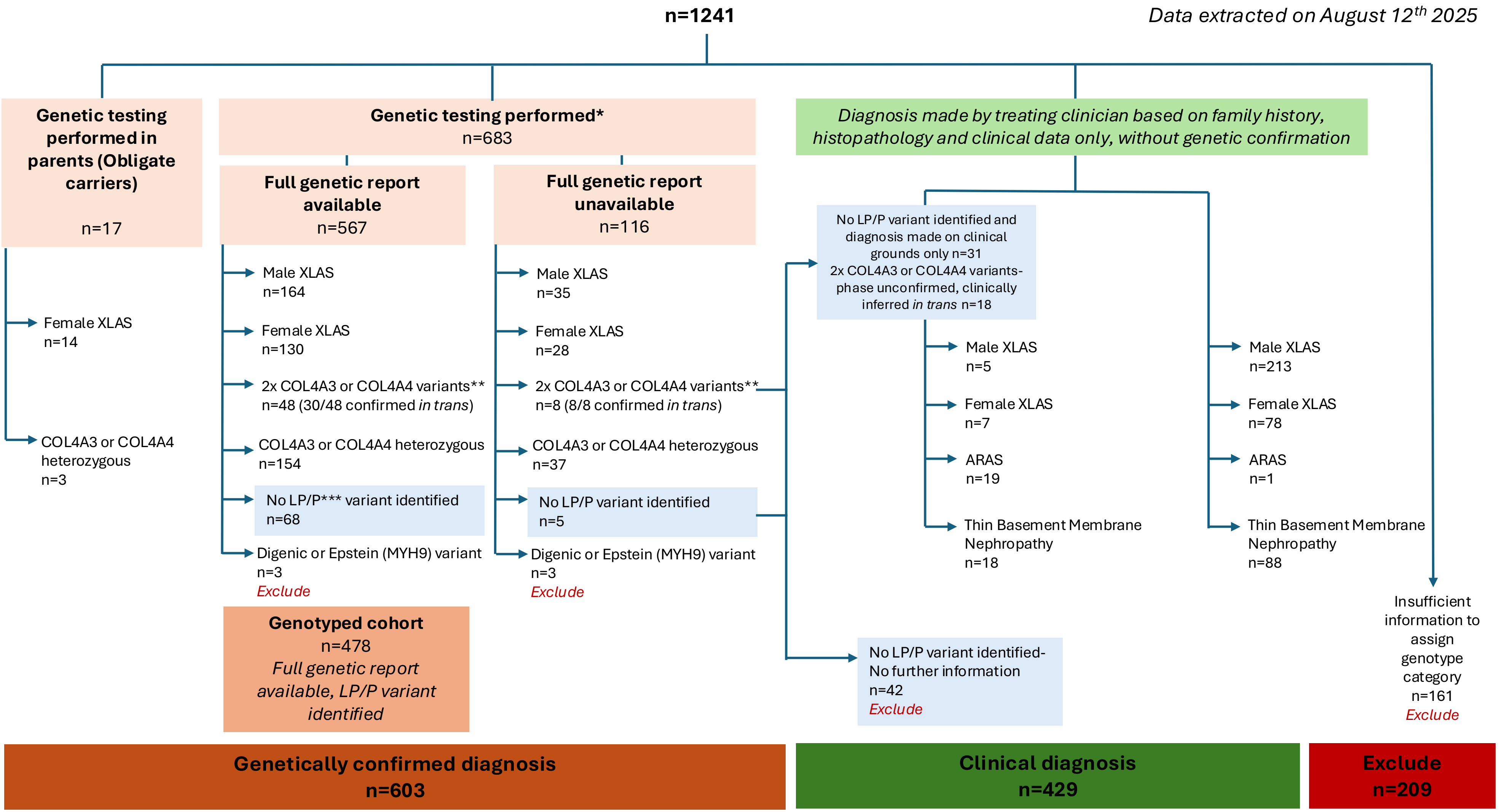


* Gene panels tested and sequencing techniques varied **Autosomal Recessive Alport Syndrome (ARAS)*** LP/P= Likely Pathogenic or Pathogenic, according to ACMG criteria

**Supplementary Table 1: Available data for each analysis**

| Variable | Cohort | | | |
|---|---|---|---|---|
| | Genetically confirmed | | Clinical diagnosis | |
| | n | (%) | n | (%) |
| Diagnosis date | 603 | (100) | 429 | (100) |
| Sex | 603 | (100) | 429 | (100) |
| Ethnicity | 459 | (76) | 369 | (86) |
| IMD Quintile | 533 | (88) | 407 | (95) |
| KF/death | 603 | (100) | 407 | (100) |
| eGFR at diagnosis | 295 | (49) | 103 | (24) |
| Eligible* for eGFR threshold analyses, *n* | | | | |
| eGFR 90 | 95 (16) | | 35 (8) | |
| eGFR 60 | 78 (13) | | 43 (10) | |
| eGFR 45 | 83 (14) | | 54 (13) | |
| UPCR in year prior to eGFR threshold available, n (%) | | | | |
| eGFR 90 | 52 | (55) | 21 | (60) |
| eGFR 60 | 53 | (68) | 28 | (65) |
| eGFR 45 | 53 | (64) | 27 | (50) |

* Sustained eGFR below the threshold (two values below the threshold at least 28 days apart without an intervening value higher than the threshold), with at least one prior eGFR above the threshold

**Supplementary Table 2: Additional demographic data**

| | Genetically confirmed | | | | Clinical Diagnosis | | | |
|---|---|---|---|---|---|---|---|---|
| | Female XLAS and heterozygotes | | Alport Syndrome | | Female XLAS and heterozygotes | | Alport Syndrome | |
| | Female XLAS | COL4A3/4 heterozygous | Male XLAS | ARAS | Female XLAS | TBMN | Male XLAS | ARAS |
| | n=172 | n=194 | n=199 | n=38 | n=85 | n=106 | n=218 | n=20 |
| **Socioeconomic status- Index of Multiple Deprivation Quintile (n, %)** | | | | | | | | |
| Not reported | 20 (11.6) | 27 (13.9) | 20 (10.1) | 3 (7.9) | 6 (7.1) | 13 (12.3) | 3 (1.4) | 0 (0) |
| 1-Most deprived | 27 (15.7) | 31 (16.0) | 43 (21.6) | 7 (18.4) | 18 (21.2) | 23 (21.7) | 35 (16.1) | 4 (20.0) |
| 2 | 33 (19.2) | 29 (14.9) | 32 (16.1) | 8 (21.1) | 19 (22.4) | 22 (20.8) | 45 (20.6) | 3 (15.0) |
| 3 | 32 (18.6) | 37 (19.1) | 37 (18.6) | 7 (18.4) | 13 (15.3) | 20 (18.9) | 45 (20.6) | 4 (20.0) |
| 4 | 28 (16.3) | 32 (16.5) | 40 (20.1) | 9 (23.7) | 13 (15.3) | 13 (12.3) | 45 (20.6) | 4 (20.0) |
| 5- Least deprived | 32 (18.6) | 38 (19.6) | 27 (13.6) | 4 (10.5) | 16 (18.8) | 15 (14.2) | 45 (20.6) | 5 (25.0) |
| **Ethnicity (n, %)** | | | | | | | | |
| Asian | ≤6 (NR) | 20 (>10) | ≤6 (NR) | ≤6 (NR) | 10 (<12) | 10 (>9) | 15 (<7) | ≤6 (NR |
| Black | ≤6 (NR) | ≤6 (NR) | ≤6 (NR) | (0) | ≤6 (NR) | (0) | ≤6 (NR) | (0) |
| Mixed | 10 (<6) | ≤6 (NR) | ≤6 (NR) | ≤6 (NR) | ≤6 (NR) | (0) | ≤6 (NR) | (0) |
| Other | ≤6 (NR) | 10 (<5) | ≤6 (NR) | 0 (0) | 0 (0) | ≤6 (NR) | ≤6 (NR) | (0) |
| White | 115 (>67) | 120 (<62) | 135 (<68) | 20 (>53) | 60 (>71) | 70 (>66) | 180 (<83) | 15 (>75) |
| Missing | 40 (>23) | 45 (<23) | 50 (<25) | 10 (>26) | 15 (<18) | 25 (<24) | 20 (>9) | ≤6 (NR |
| **Comorbidities (n, %)** | | | | | | | | |
| Comorbidity data available | 158 (91.9) | 180 (92.8) | 177 (88.9) | 34 (89.5) | 76 (89.4) | 92 (86.8) | 194 (89.0) | 18 (90.0) |
| Hypertension pre-diagnosis | 14 (8.9) | 35 (19.4) | 35 (19.8) | 8 (23.5) | 11 (14.5) | 16 (17.4) | 37 (19.1) | 5 (27.8) |
| Hypertension pre-KF | 132 (83.5) | 130 (72.2) | 135 (76.3) | 25 (73.5) | 48 (63.2) | 51 (55.4) | 139 (71.6) | 13 (72.2) |
| Diabetes pre-diagnosis | 0 (0) | 3 (1.7) | 1 (0.6) | 0 (0) | 0 (0) | 3 (3.3) | 4 (2.1) | 1 (5.6) |
| Diabetes Pre-KF | 3 (1.9) | 3 (1.7) | 4 (2.3) | 0 (0) | 1 (1.3) | 7 (7.6) | 2 (1.0) | 0 (0) |
| **ACE inhibitor or Angiotensin receptor blocker usage (n, %)** | | | | | | | | |
| ACE-I or ARB data pre-KRT | 78 (45.3) | 104 (53.6) | 119 (60.0) | 24 (63.2) | 30 (35.2) | 46 (43.3) | 43 (19.7) | 9 (45.0) |
| ACE-I or ARB pre-KRT | 64 (82.1) | 72 (69.2) | 105 (88.2) | 24 (100.0) | 19 (63.3) | 30 (65.2) | 32 (74.4) | 7 (77.8) |

NR= Not reported. Numbers ≤6 per cell have been suppressed, and corresponding percentages rounded to nearest 5 to avoid inadvertent calculation of counts. ARAS= Autosomal Recessive Alport Syndrome**.** TBMN = Thin basement membrane nephropathy KRT=Kidney Replacement Therapy

**Supplementary Table 3: Clinical demographics for genetically confirmed and clinically diagnosed cohorts**

| | Alport syndrome genotypes | | | Heterozygous genotypes | | |
|---|---|---|---|---|---|---|
| | Genetically confirmed | Clinical diagnosis | P-value* | Genetically confirmed | Clinical diagnosis | P-value |
| Female (n%) | 20 (8.4) | 10 (4.2) | 0.06 | 293 (80.1) | 158 (82.7) | 0.45 |
| White ethnicity (n%) | 154 (86.5) | 193 (90.6) | 0.20 | 232 (82.6) | 133 (85.8) | 0.38 |
| Current age (Years, IQR) | 26.5 (17.2-41.6) | 50.5 (38.7-61.1) | <0.0001 | 36.5 (20.4-53.8) | 54.1 (40.6-66.0) | <0.0001 |
| Year of diagnosis (Median) | 2015 | 2003 | <0.0001 | 2017 | 2007 | <0.0001 |
| Age at diagnosis (Median Years, IQR) | 15.3 (7.5-29.7) | 22.4 (9.4-35.8) | 0.007 | 27.2 (10.4-45.3) | 31.9 (19.9-46.2) | 0.0016 |
| eGFR at diagnosis (ml/min/1.73m$^2$, IQR) | 79.3 (37.8-111.1) | 57.1 (22.5-99.1) | 0.01 | 92.3 (52.7-110.2) | 72.0 (45.3-103.6) | 0.081 |
| Follow up time | 10.6 (6.7-13.1) | 23.1 (12.3-33.4) | <0.0001 | 8.9 (5.5-12.2) | 17.9 (10.7-26.4) | <0.0001 |
| KF events (n%) | 98(41.4) | 193 (81.1) | <0.0001 | 51 (13.9) | 69 (36.1) | <0.0001 |

*Chi-Square and Kruskall-Wallis tests

**Supplementary Figure 2: Diagnosis year for genetically confirmed and clinically diagnosed subgroups, for a) Alport Syndrome and b) Heterozygous genotypes**

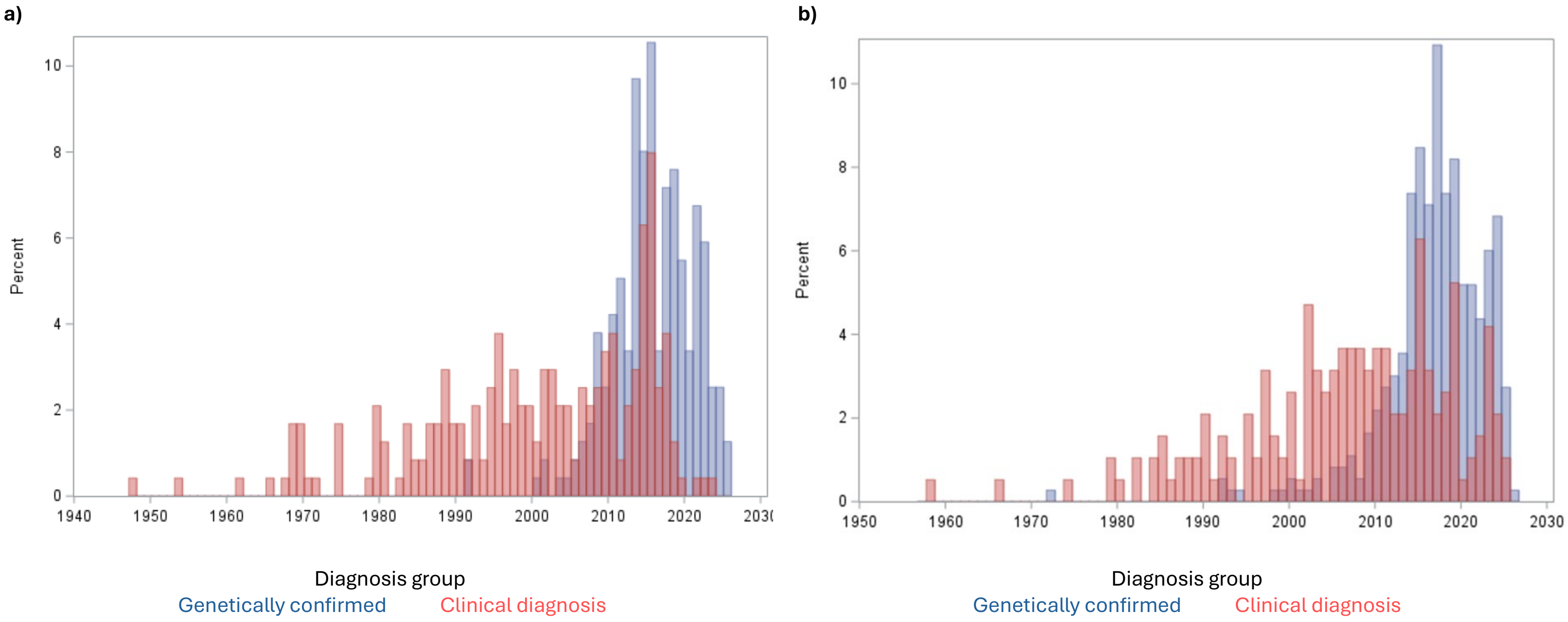

**Supplementary Table 4: Variant classification**

| | **Male X-linked Alport Syndrome** | **ARAS*** | **Female X-linked Alport Syndrome** | **COL4A3/4 Heterozygous** |
|---|---|---|---|---|
| **Protein length altering variants **** | **n=68** | **n=25** | **n=58** | **n=63** |
| Exon deletion | 12 (17.6) | 0 (0) | 8 (13.8) | 3 (4.8) |
| Exon duplications | 6 (8.8) | 0 (0) | 3 (5.2) | (.0) |
| Frameshift | 17 (25.0) | 11 (44.0) | 11 (19.0) | 7 (11.1) |
| Insertion/deletion/Insertion and deletion | 0 (0) | 0 (0) | 2 (3.4) | 2 (3.2) |
| Splice site variant | 22 (32.4) | 1 (4.0) | 23 (39.7) | 19 (30.2) |
| Stop gain or stop loss | 11 (16.2) | 13 (52.0) | 11 (18.9) | 32 (50.8) |
| **Non-protein length altering variants** | **n=96** | **n=5** | **n=72** | **n=91** |
| Glycine substitution | 82 (85.4) | 5 (100.0) | 65 (90.3) | 76 (83.5) |
| Non-glycine substitution | 9 (9.4) | (.0) | 1 (1.4) | 8 (8.8) |
| Small insertion/deletion (<20 base pairs) | 5 (5.2) | (.0) | 6 (8.3) | 7 (7.7) |
| **Glycine substitutions** | | | | |
| Non-destabilising substitution | 15 (18.3) | 0 (0.0) | 16 (24.6) | 8 (10.5) |
| Destabilising substitution*** | 67 (81.7) | 5 (2.2) | 49 (75.4) | 68 (89.5) |
| Collagenous domain variant | 71 (86.6) | 4 (80.0) | 56 (86.2) | 72 (94.7) |
| Non collagenous (NC) domain variant | 11 (13.4) | 1 (0.4) | 9 (13.8) | 4 (5.3) |
| Exons 1-20 | 13 (15.9) | 1 (20.0) | 6 (9.2) | 13 (17.1) |
| Exons 20-carboxyl terminus | 69 (84.1) | 4 (1.8) | 59 (90.8) | 63 (82.9) |
| Exons 1-20 | 13 (16.7) | 1 (25.0) | 6 (10.0) | 13 (17.6) |
| Exons 20-last NC domain | 65 (83.3) | 3 (75.0) | 54 (90.0) | 61 (82.4) |

*ARAS; Autosomal Recessive Alport Syndrome – 2 likely pathogenic or pathogenic variants in either *COL4A3* or *COL4A4*, confirmed *in trans* **For patients with 2 differing variant types, most damaging variant used for analyses *** Mildly destabilising: Ala, Ser, Cys; Highly destabilising: Arg, Val, Glu, Asp, Trp

**Supplementary Figure 3: $25^{th}$ centile Age at Kidney Failure, by genotype**

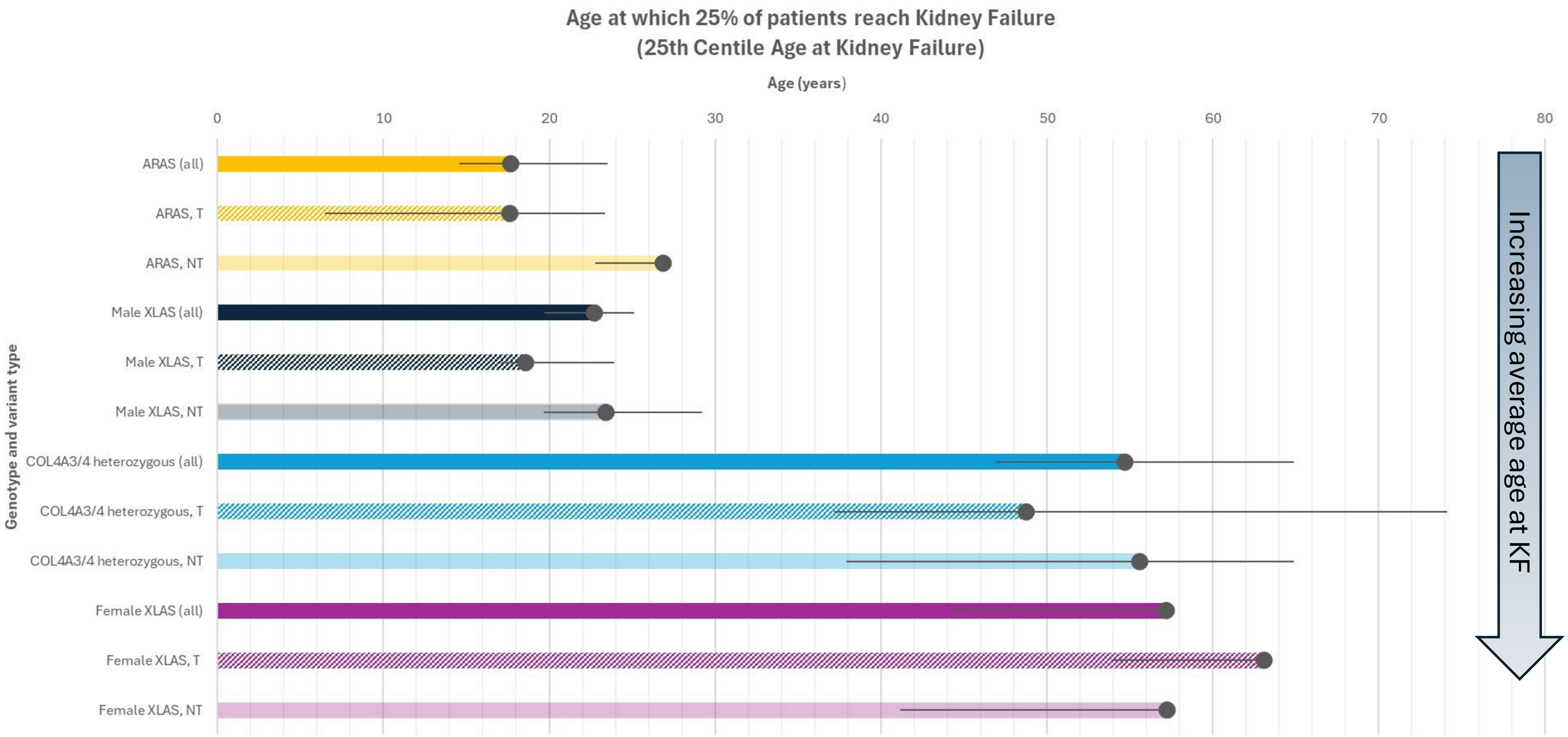


| | Female XLAS, NT | Female XLAS, T | Female XLAS (all) | COL4A3/4 heterozygous, NT | COL4A3/4 heterozygous, T | COL4A3/4 heterozygous (all) | Male XLAS, NT | Male XLAS, T | Male XLAS (all) | ARAS, NT | ARAS, T | ARAS (all) |
|---|---|---|---|---|---|---|---|---|---|---|---|---|
| 25% age at KF | 57.2 | 63.1 | 57.2 | 55.6 | 48.8 | 54.7 | 23.4 | 18.6 | 22.7 | 26.9 | 17.7 | 17.7 |

ARAS= Autosomal Recessive Alport Syndrome XLAS= X-linked Alport Syndrome T= Truncating or Protein length altering variant NT= Non truncating or Non protein length altering variant. Error bars shown are 95% Confidence Intervals. *For female XLAS, no upper 95% CI limits were estimable

**Supplementary Figure 4: Age at kidney failure, stratified by variant type for a) Male X-Linked AS b) Autosomal Recessive Alport Syndrome c) Female X-Linked AS d) COL4A3/4 heterozygous variants**

**a) Male X-Linked Alport Syndrome**

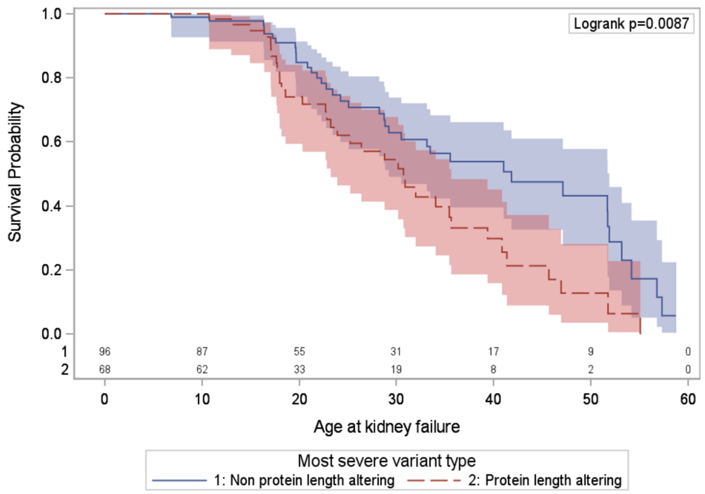


**b) ARAS**

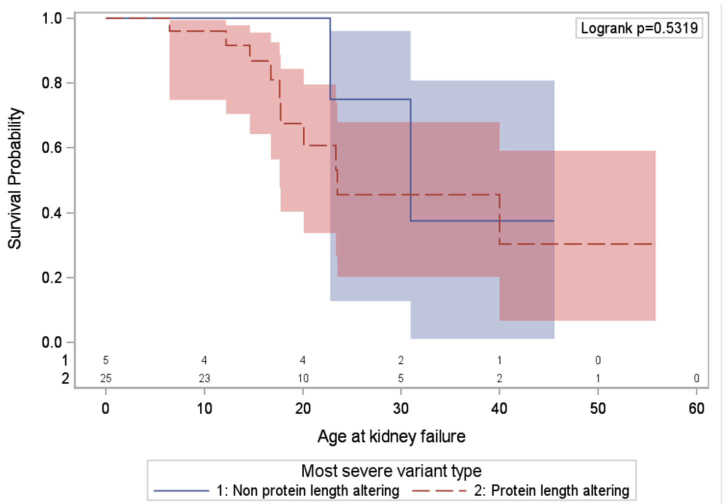


**c) Female X-Linked Alport Syndrome**

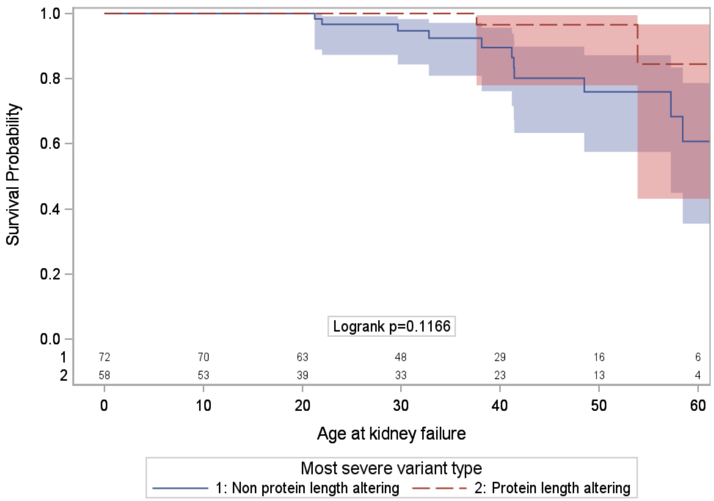


**d) COL4A3/4 heterozygous variants**

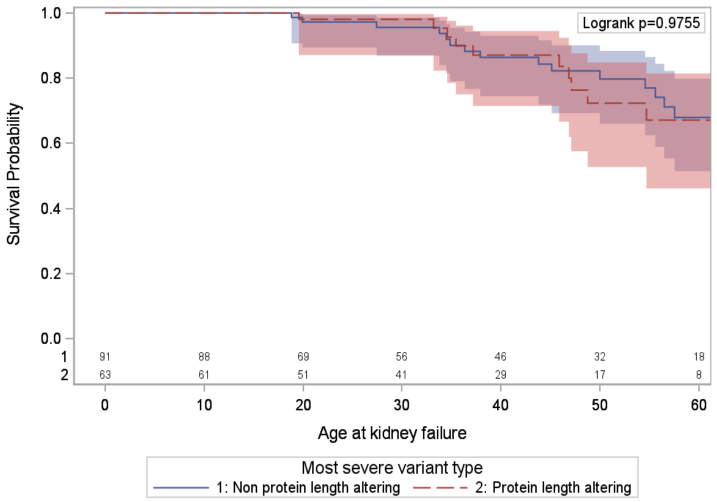


*genotyped cohort

**Supplementary Figure 5: Age at Kidney Failure for those with stop gain variants, other protein length altering and non-protein length altering variants, for a) Alport Syndrome b) heterozygous genotypes***

a)

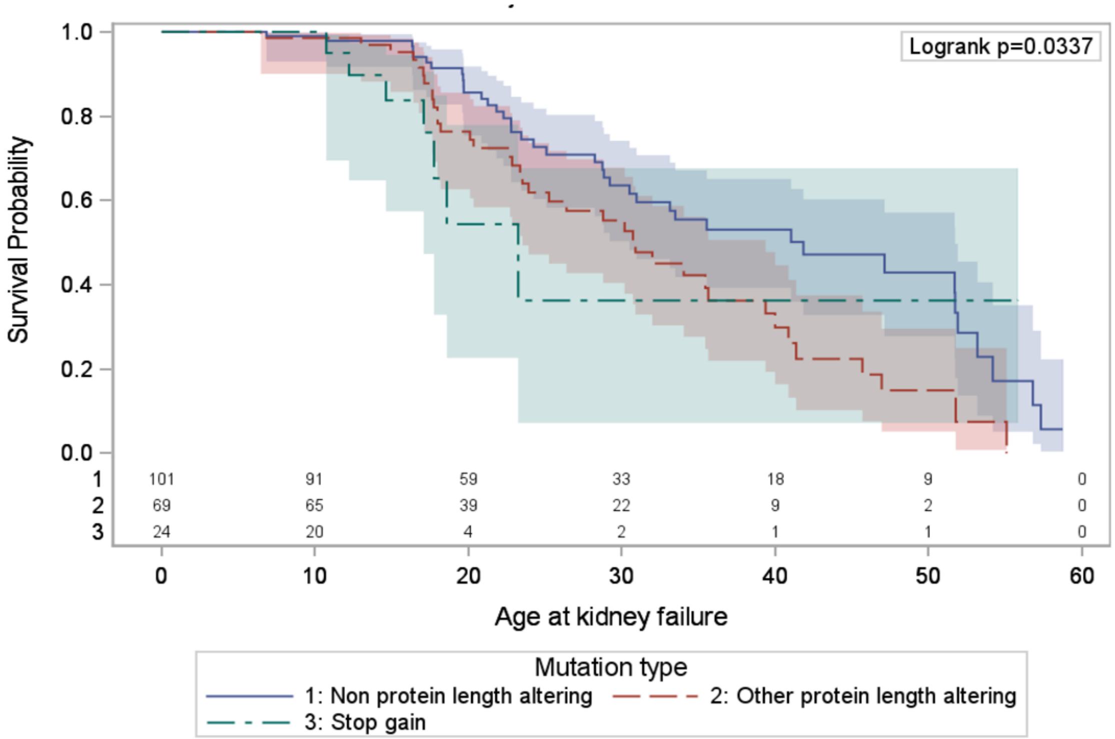


b)

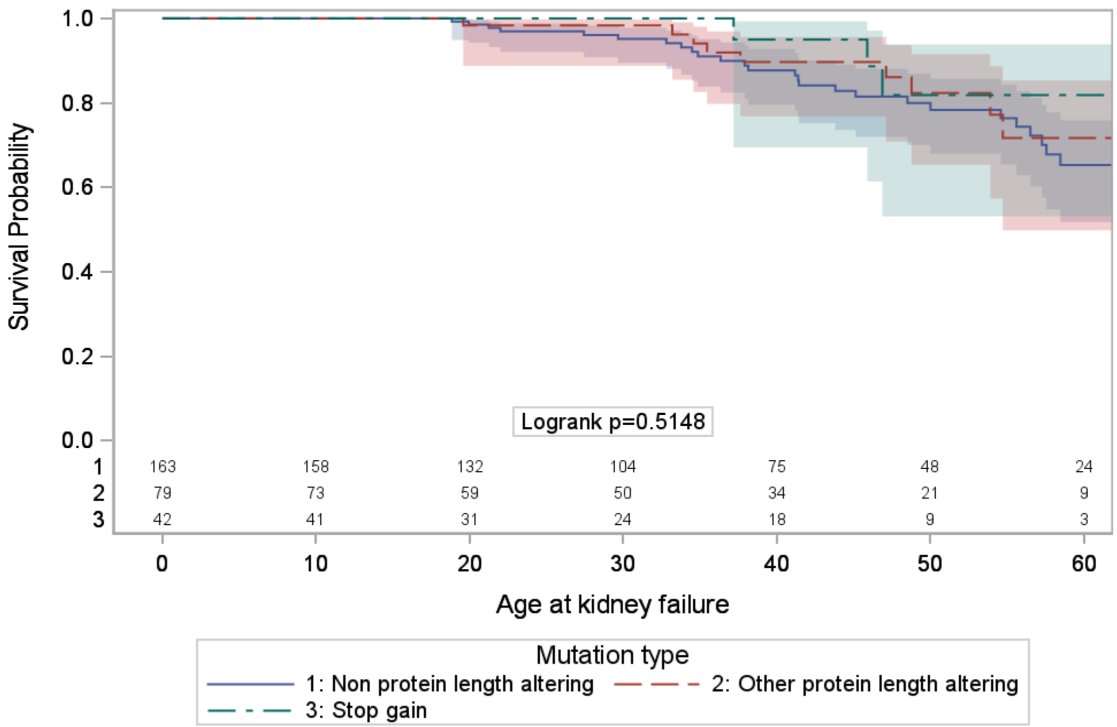


*genotyped cohort

**Supplementary Figure 6: Kaplan Meier plots of age at KF for patients with Male XLAS and 2 COL4A3 or 2 COL4A4 variants with a) protein length-altering b) non protein length-altering variants**

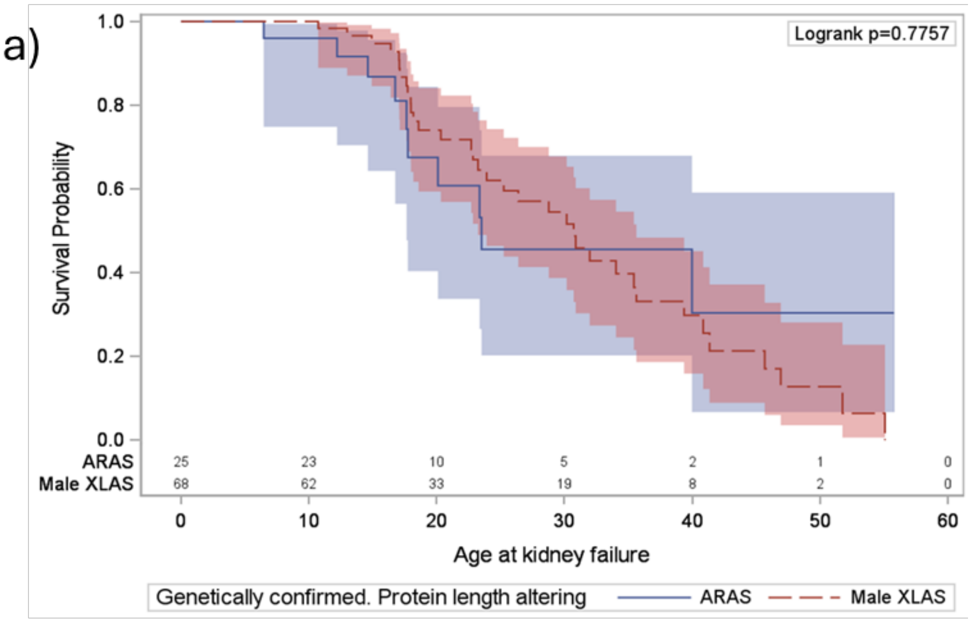


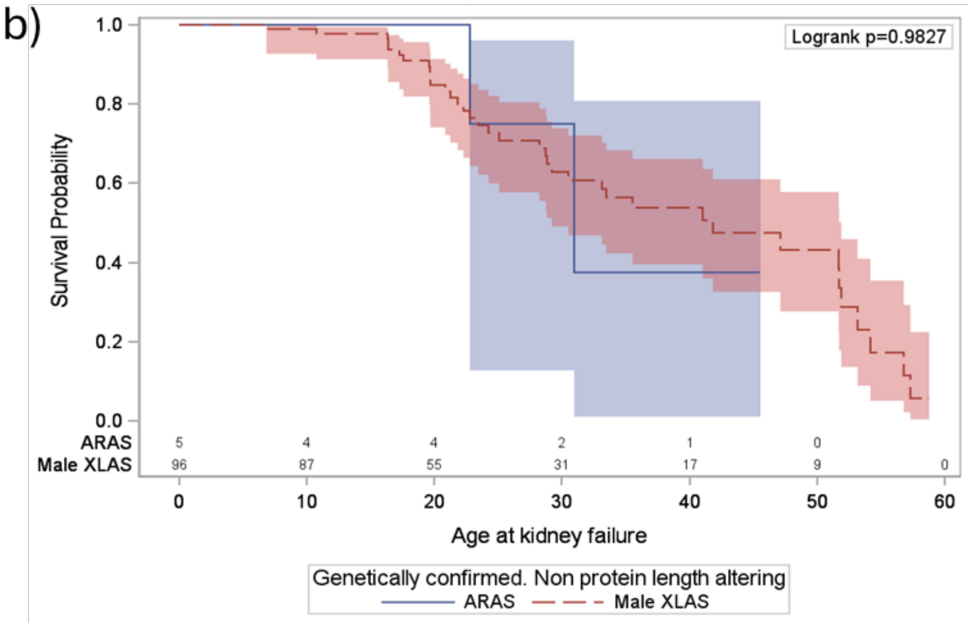


*Patients with 2x COL4A3 or COL4A4 variants with >1 variant type excluded

**Supplementary Table 5: Distribution of protein length altering and non-protein length altering variants by genotype**

| | Variant type | | | | | | |
|---|---|---|---|---|---|---|---|
| | **Non protein length altering** | | **Protein length altering** | | **Total** | | **Chi2 p-value** |
| **Genotype** | n | (%) | n | (%) | n | (%) | |
| ARAS* | 20 | (33.3) | 40 | (66.7) | 60 | (100) | 0.001 |
| COL4A3/4 heterozygous | 91 | (59.1) | 63 | (40.9) | 154 | (100) | |
| Male XLAS | 98 | (59.0) | 68 | (41.0) | 166 | (100) | |

ARAS; Autosomal Recessive Alport Syndrome – 2 likely pathogenic or pathogenic variants in either *COL4A3* or *COL4A4*, both variants included in counts. 2 patients with Male XLAS had 2 pathogenic/likely pathogenic variants identified; both variants included in counts. Female XLAS excluded as ascertainment will be affected by severity in Male relatives.

**Supplementary Figure 7: Age at kidney failure, stratified by molecular characteristics a) Destabilising residue b) In or adjacent to Non-collagenous domain c) Exon position d) Exon position excluding last Non-collagenous (NC) domain for Alport Syndrome and heterozygous genotypes**

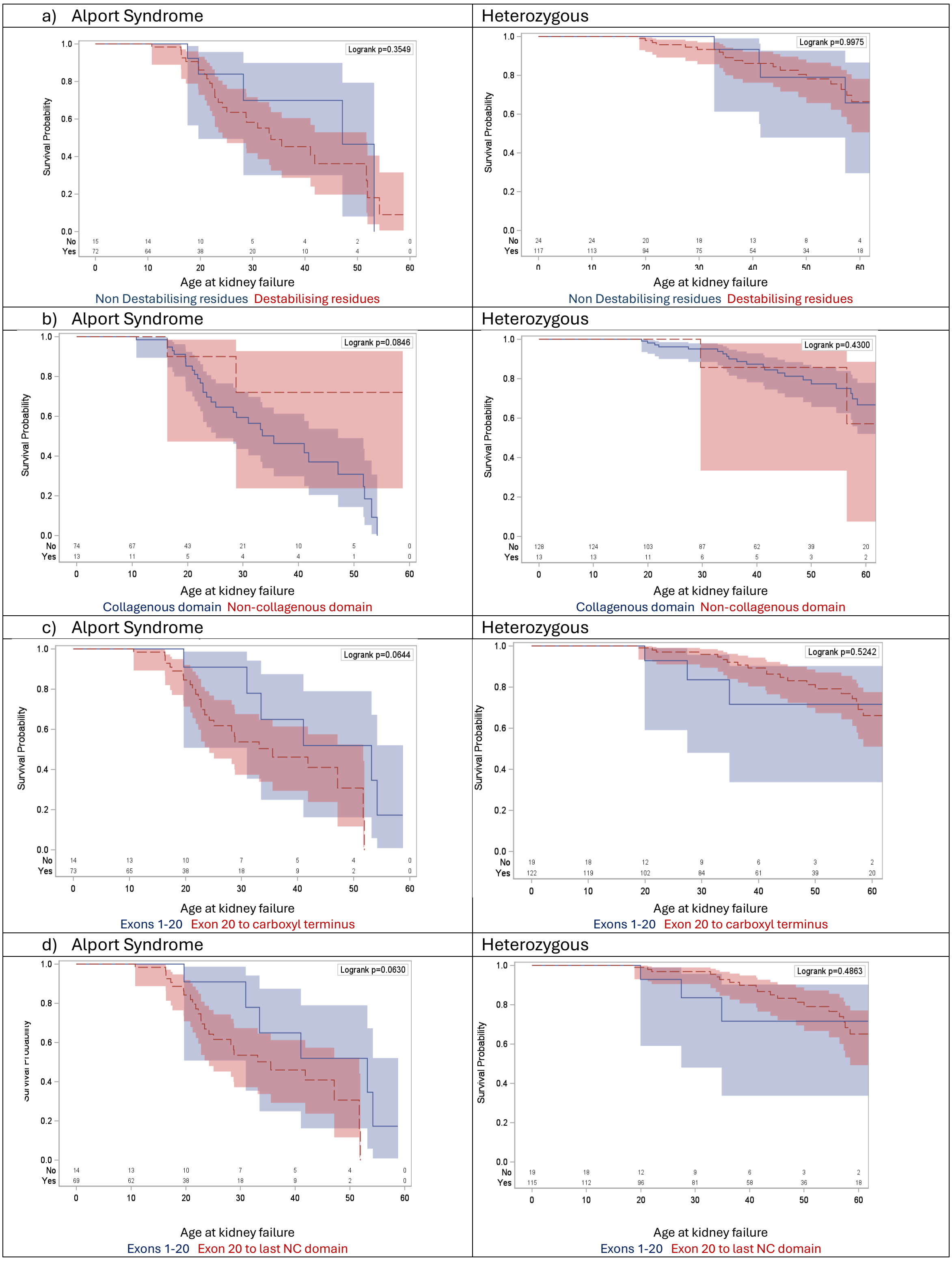

**Supplementary Figure 8 : Patient distribution across proteinuria levels (<0.5g/g, 0.5-1.0g/g, >1.0g/g) by age**

### Alport Syndrome

a) Including patients not yet reaching specified age

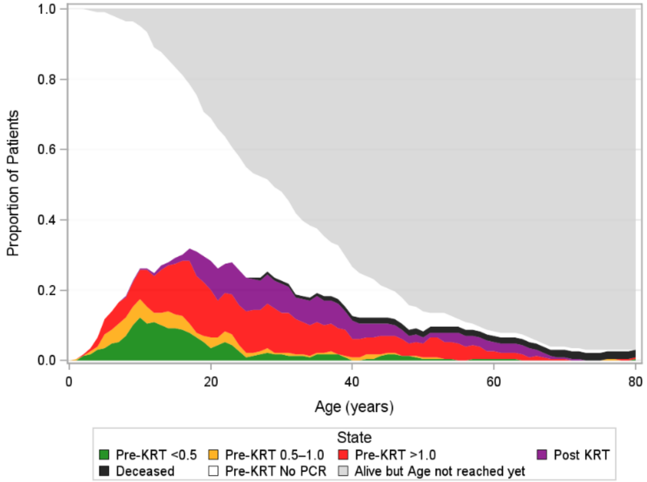


b) Excluding patients not yet reaching specified age

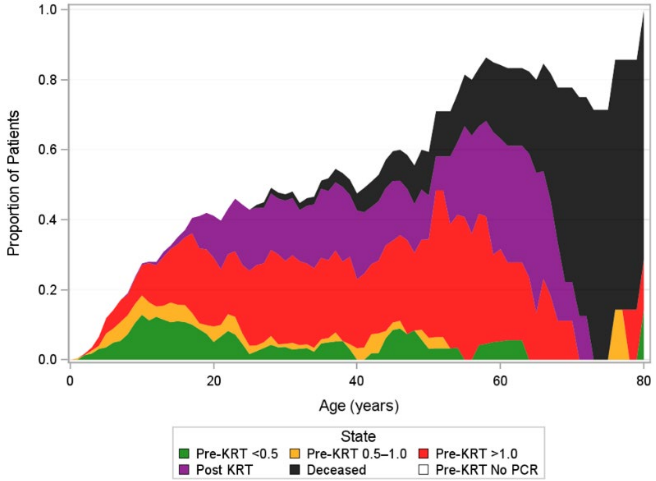


### Heterozygous

c) Including patients not yet reaching specified age

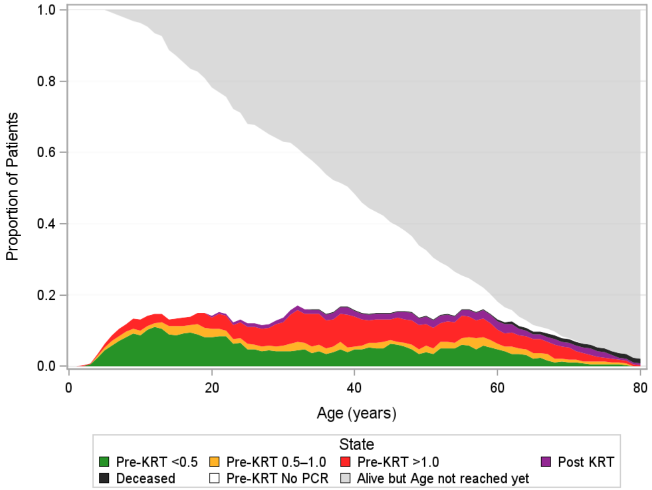


d) Excluding patients not yet reaching specified age

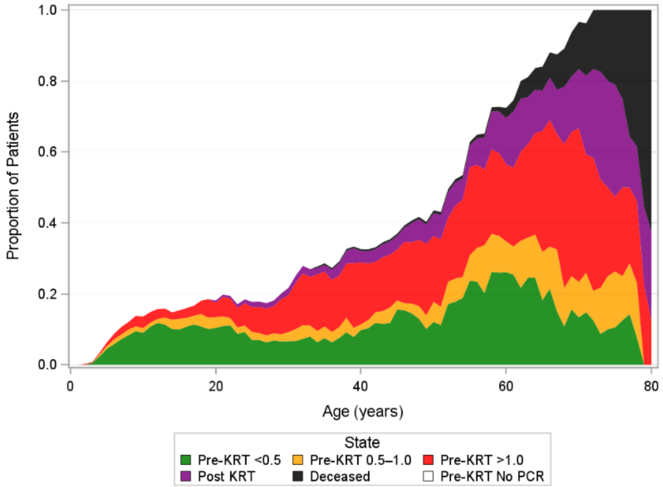

**Supplementary Figure 9: Patient distribution across CKD stages, by age**

#### Alport Syndrome

a) Including patients not yet reaching specified age

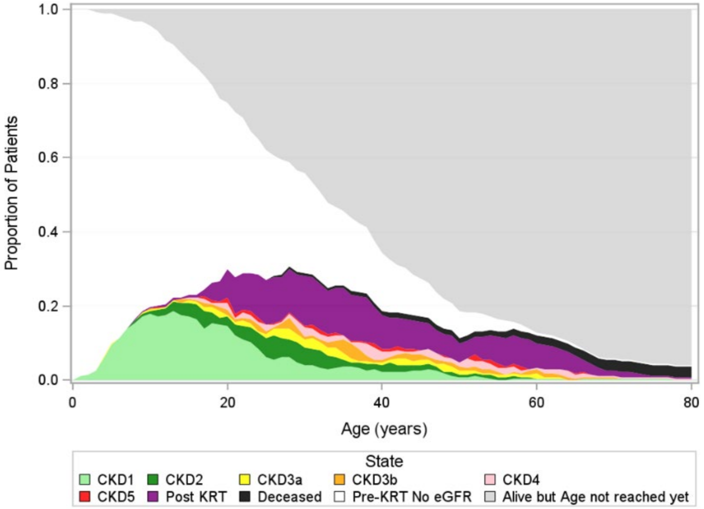


b) Excluding patients not yet reaching specified age

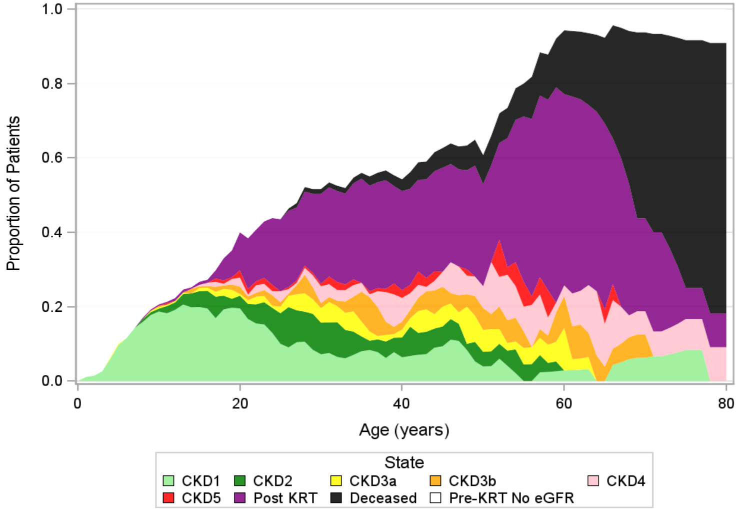


#### Heterozygous

c) Including patients not yet reaching specified age

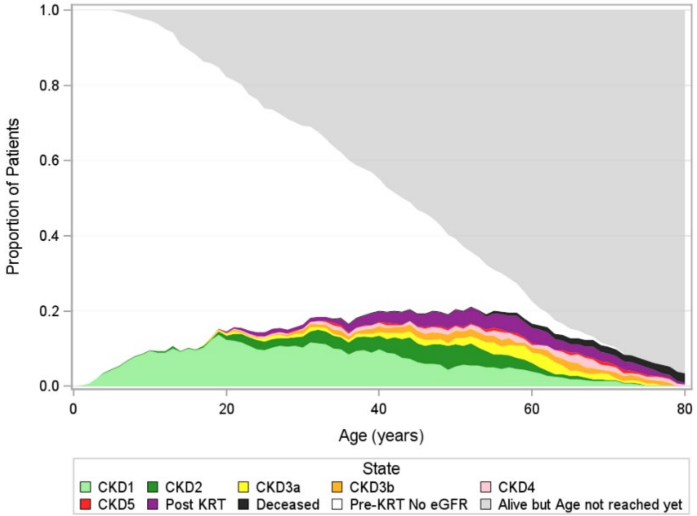


d) Excluding patients not yet reaching specified age

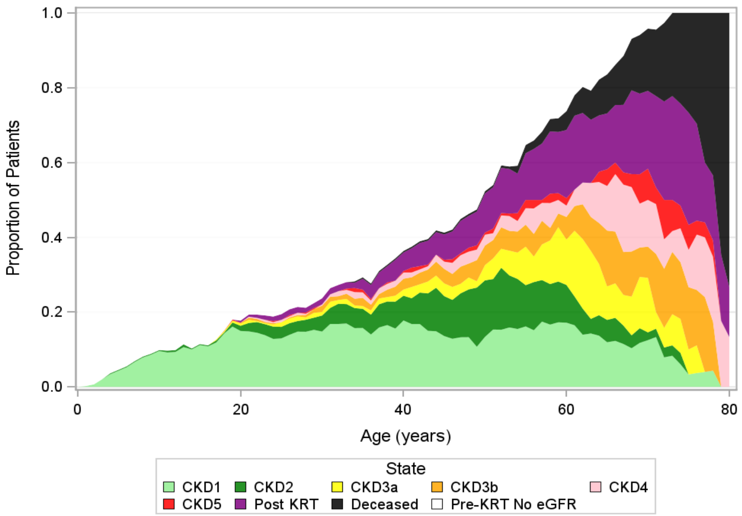

**Supplementary Table 6: Annualised eGFR slope (ml/min/1.73m$^2$/year), by genotype**

| | | Female XLAS and heterozygous | | | | Alport Syndrome | | | |
|---|---|---|---|---|---|---|---|---|---|
| | | Genetically confirmed | | Clinical diagnosis | | Genetically confirmed | | Clinical diagnosis | |
| | | Female XLAS | COL4A3/4 Heterozygous | Female XLAS | TBMN | Male XLAS | ARAS | Male XLAS | ARAS |
| Mean annualised eGFR slope (ml/min/1.73m$^2$/year), 95% CI | CKD stage 1 | -0.3 (-0.9 to 0.3) | -0.7 (-1.2 to -0.1) | 0.2 (-0.8 to 1.2) | 0.0 (-0.8 to 0.8) | -1.4 (-2.0 to -0.8) | -1.4 (-2.6 to -0.3) | -0.8 (-1.6 to 0.1) | 0.2 (-1.8 to 2.2) |
| | CKD stage 2 | -1.2 (-1.8 to -0.6) | -1.8 (-2.3 to -1.2) | -1.4 (-2.3 to -0.4) | -1.5 (-2.3 to -0.7) | -4.2 (-4.7 to -3.6) | -4.5 (-5.7 to -3.4) | -3.2 (-4.0 to -2.3) | -4.4 (-6.6 to -2.3) |
| | CKD stage 3a | -2.0 (-2.7 to -1.4) | -3.4 (-4.0 to -2.9) | -2.9 (-3.9 to -2.0) | -2.0 (-2.8 to -1.3) | -5.3 (-5.9 to -4.7) | -6.0 (-7.1 to -4.8) | -4.8 (-5.7 to -4.0) | -5.6 (-7.7 to -3.6) |
| | CKD stage 3b | -2.6 (-3.2 to -2.0) | -3.7 (-4.2 to -3.1) | -3.3 (-4.2 to -2.3) | -2.7 (-3.5 to -1.9) | -5.9 (-6.5 to -5.3) | -6.9 (-8.0 to -5.7) | -5.9 (-6.7 to -5.1) | -6.3 (-8.3 to -4.4) |
| | CKD stage 4 | -2.9 (-3.6 to -2.3) | -3.8 (-4.4 to -3.3) | -3.8 (-4.7 to -2.8) | -2.8 (-3.6 to -2.0) | -6.4 (-7.0 to -5.8) | -8.1 (-9.2 to -6.9) | -6.4 (-7.2 to -5.5) | -6.8 (-8.8 to -4.8) |
| Time from eGFR 30 to 15 ml/min/1.73m$^2$ or KRT initiation, years, 95% CI | | 5.1 (4.2 to 6.5) | 3.9 (3.4 to 4.6) | 4.0 (3.2 to 5.3) | 5.3 (4.1 to 7.4) | 2.4 (2.2 to 2.6) | 1.9 (1.6 to 2.2) | 2.4 (2.1 to 2.7) | 2.2 (1.7 to 3.1) |

| Annualised eGFR slope | Key |
|---|---|
| <1 ml/min/1.73m$^2$/year | |
| 1 to <3 ml/min/1.73m$^2$/year | |
| 3 to 5 ml/min/1.73m$^2$/year | |
| >5 ml/min/1.73m$^2$/year | |

**Supplementary Figure 10: Annualised eGFR slope by CKD stage, for Alport Syndrome and Heterozygous genotypes for clinically diagnosed cohort**

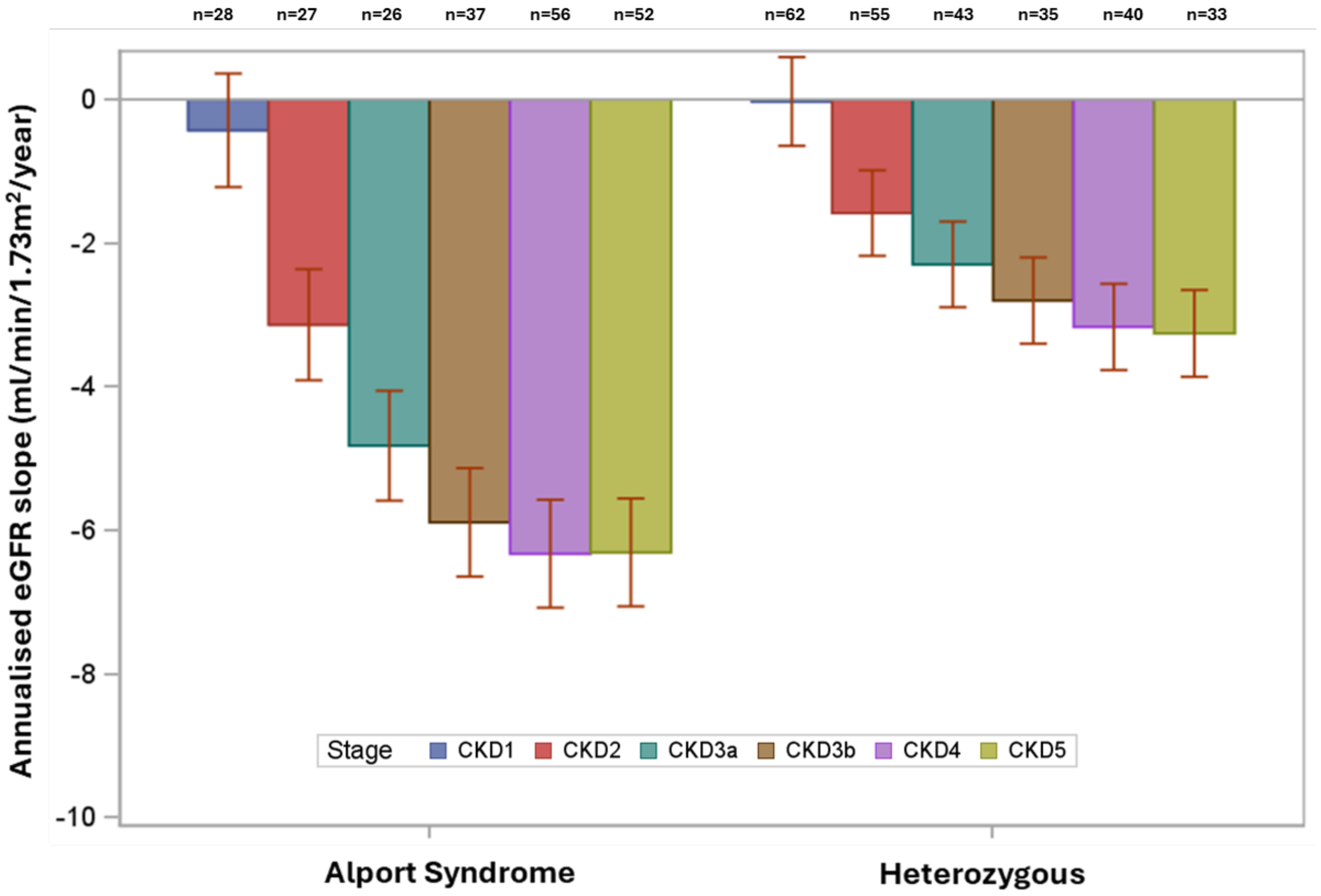


| | Alport Syndrome CKD1 | Alport Syndrome CKD2 | Alport Syndrome CKD3a | Alport Syndrome CKD3b | Alport Syndrome CKD4 | Alport Syndrome CKD5 | Heterozygous CKD1 | Heterozygous CKD2 | Heterozygous CKD3a | Heterozygous CKD3b | Heterozygous CKD4 | Heterozygous CKD5 |
|---|---|---|---|---|---|---|---|---|---|---|---|---|
| Annualised eGFR slope, 95% CI | **-0.43** (-1.22 to 0.36) | **-3.14** (-3.91 to -2.36) | **-4.82** (-5.59 to -4.06) | **-5.89** (-6.65 to -5.14) | **-6.33** (-7.08 to -5.58) | **-6.31** (-7.06 to -5.56) | **-0.03** (-0.65 to 0.59) | **-1.58** (-2.18 to -0.99) | **-2.30** (-2.90 to -1.70) | **-2.80** (-3.40 to -2.20) | **-3.17** (-3.77 to -2.57) | **-3.26** (-3.86 to -2.65) |

**Supplementary Figure 11: Annualised eGFR slope by CKD stage, for Alport Syndrome and Heterozygous genotypes, using CKD-EPI 2021 or bedside Schwartz equations for a) genetically confirmed and b) clinically diagnosed cohorts**

a)

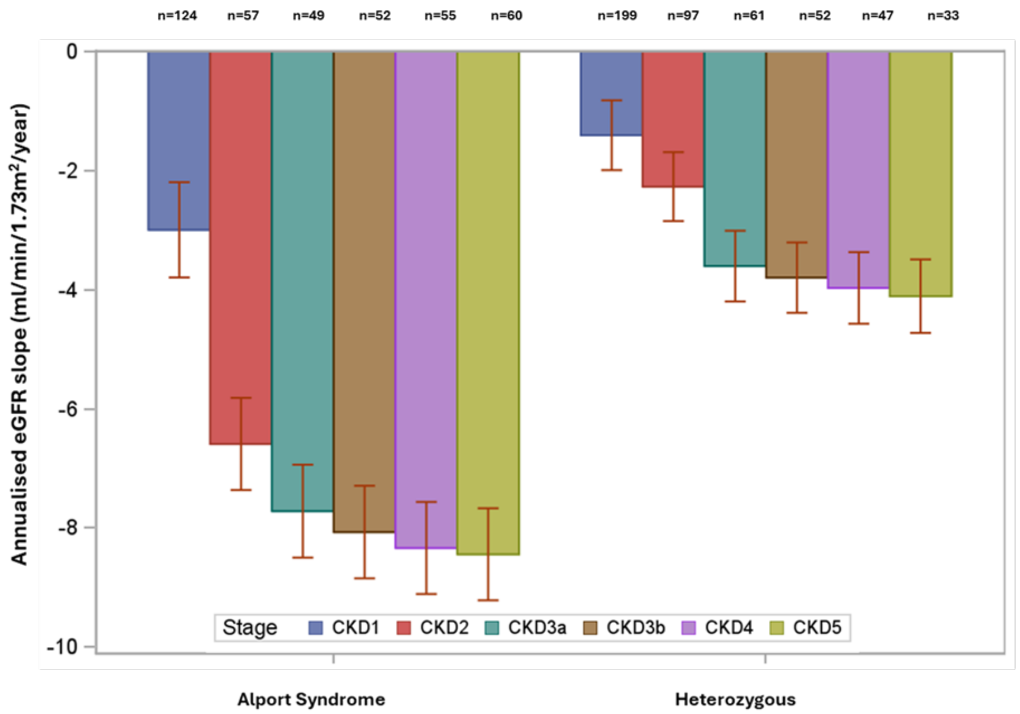


Alport Syndrome

| | CKD1 | CKD2 | CKD3a | CKD3b | CKD4 | CKD5 |
|---|---|---|---|---|---|---|
| Annualised eGFR slope, 95% CI | -3.00 (-3.80 to -2.20) | -6.59 (-7.36 to -5.82) | -7.72 (-8.50 to -6.94) | -8.07 (-8.85 to -7.29) | -8.34 (-9.11 to -7.56) | -8.44 (-9.22 to -7.67) |

Heterozygous

| | CKD1 | CKD2 | CKD3a | CKD3b | CKD4 | CKD5 |
|---|---|---|---|---|---|---|
| Annualised eGFR slope, 95% CI | -1.41 (-1.99 to -0.82) | -2.27 (-2.85 to -1.69) | -3.60 (-4.20 to -3.01) | -3.80 (-4.39 to -3.21) | -3.97 (-4.57 to -3.37) | -4.11 (-4.73 to -3.49) |

b)

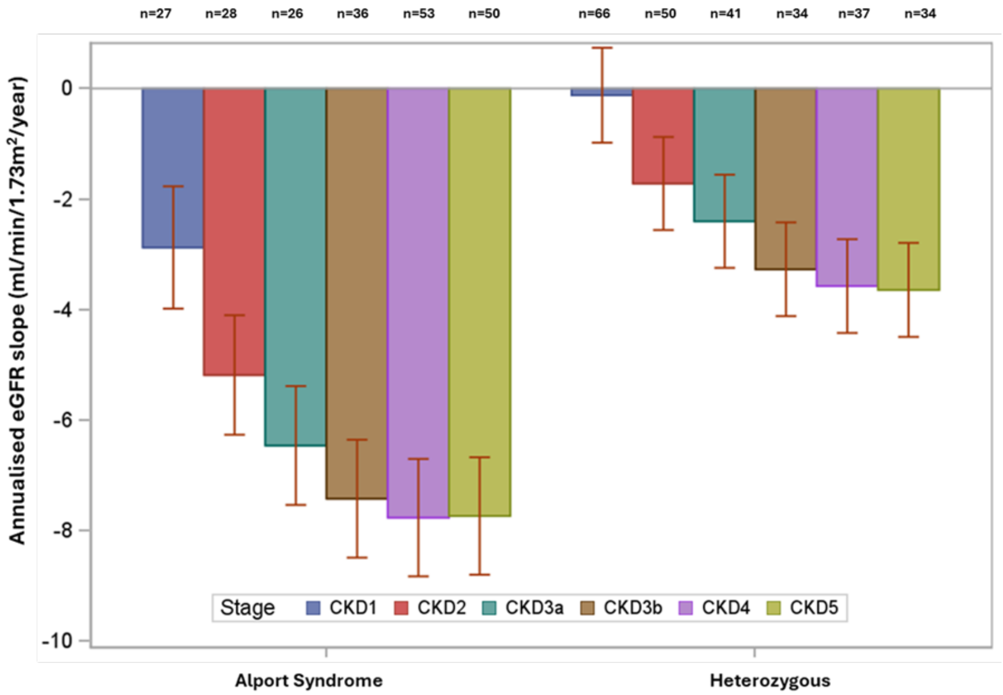


Alport Syndrome

| | CKD1 | CKD2 | CKD3a | CKD3b | CKD4 | CKD5 |
|---|---|---|---|---|---|---|
| Annualised eGFR slope, 95% CI | -2.88 (-3.98 to -1.77) | -5.19 (-6.27 to -4.11) | -6.46 (-7.54 to -5.39) | -7.43 (-8.49 to -6.36) | -7.77 (-8.83 to -6.70) | -7.74 (-8.80 to -6.68) |

Heterozygous

| | CKD1 | CKD2 | CKD3a | CKD3b | CKD4 | CKD5 |
|---|---|---|---|---|---|---|
| Annualised eGFR slope, 95% CI | -0.12 (-0.98 to 0.74) | -1.72 (-2.56 to -0.88) | -2.40 (-3.25 to -1.56) | -3.27 (-4.12 to -2.42) | -3.58 (-4.43 to -2.73) | -3.65 (-4.50 to -2.80) |

*CKD-EPI 2021, bedside Schwartz for those aged <16 years

**Supplementary Figure 12: Linear mixed model of proteinuria by age, stratified by genotype for genetically confirmed cohort**

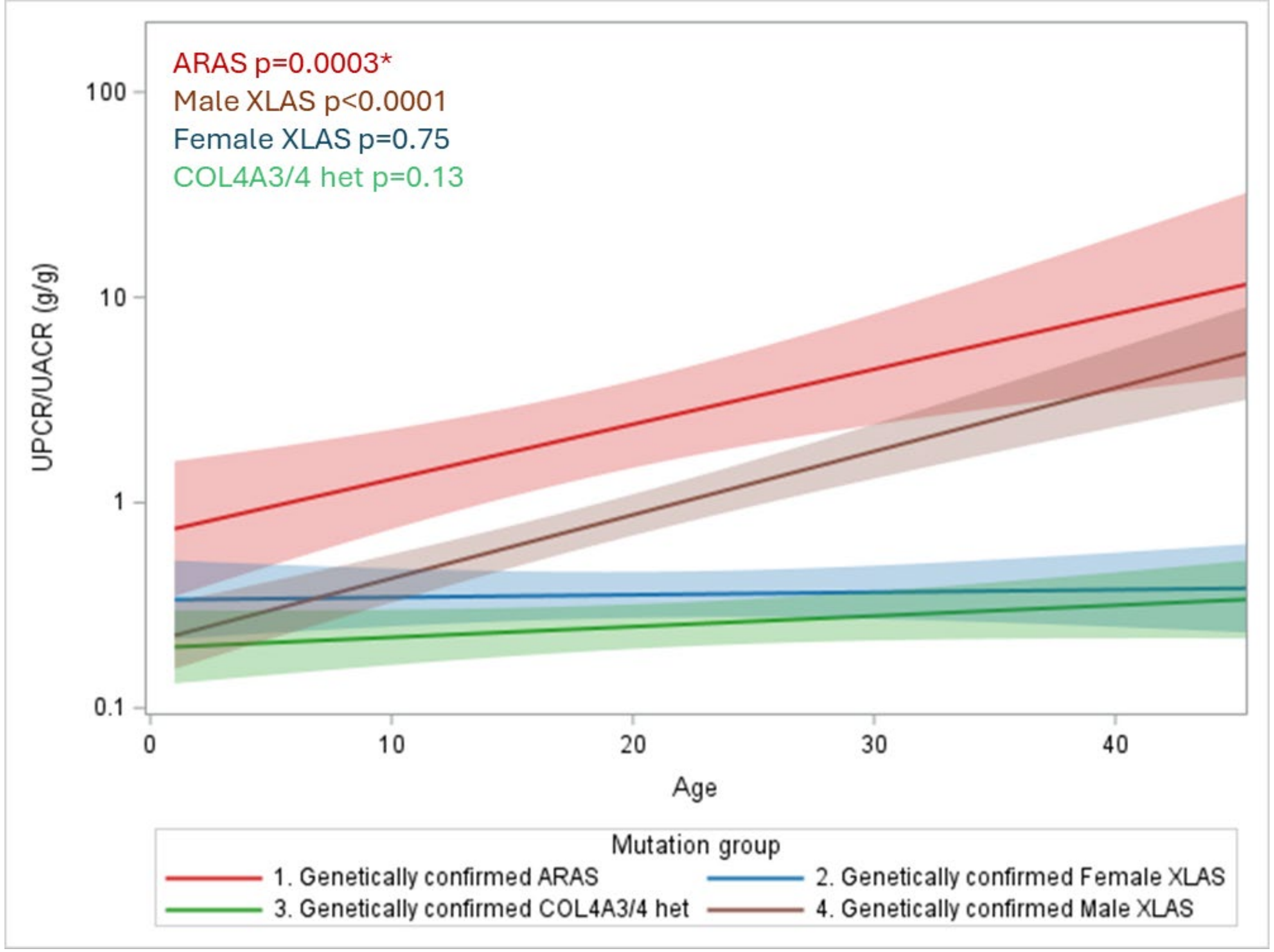


*Statistical testing performed against zero slope

**Supplementary Figure 13: Non-linear model of proteinuria trajectory by age for a) Males and b) Females, stratified by genotype for genetically confirmed cohort**

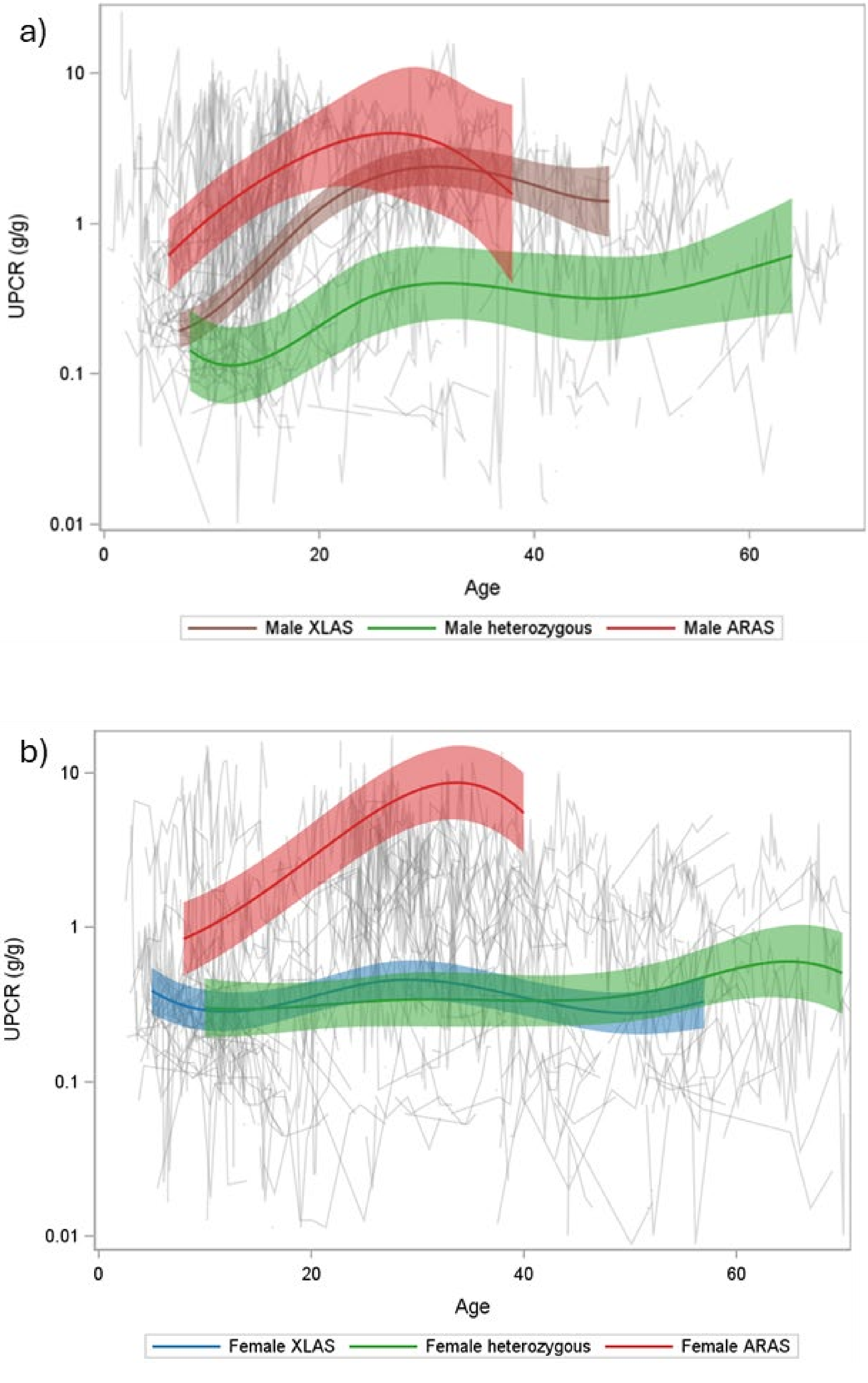

**Supplementary Figure 14: a) Cumulative incidence plot of age at proteinuria thresholds for all patients and b) stratified by Alport Syndrome and Heterozygous genotypes, for genetically confirmed cohort**

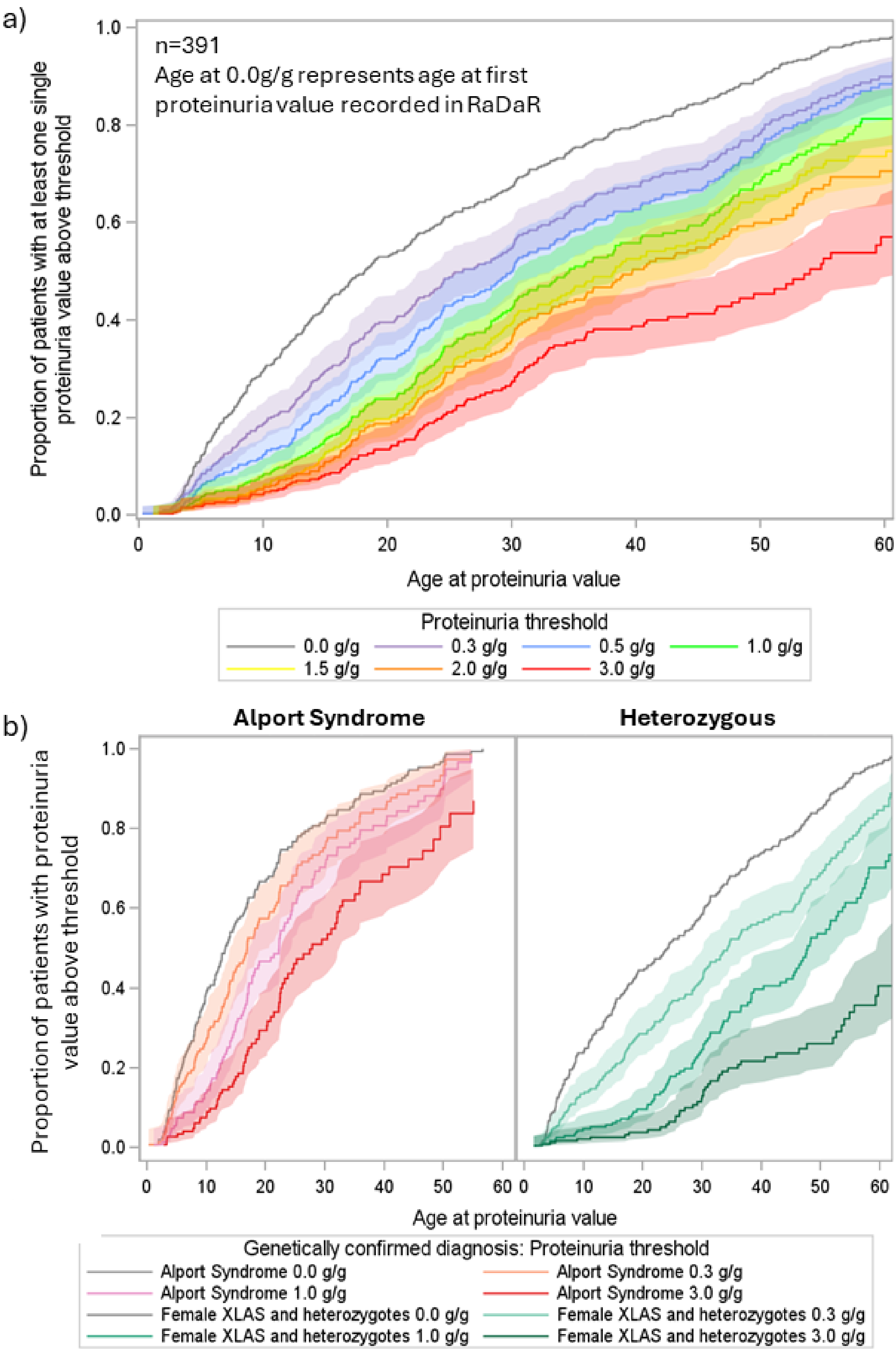


*For genetically confirmed cohort only

**Supplementary Figure 15: Kaplan Meier plots of time from reaching proteinuria thresholds of a) ≥0.3g/g b) ≥1.0 g/g c) ≥3.0 g/g to kidney failure , where date of reaching a proteinuria threshold was defined as a) date of first single value above that threshold, b) date of first value, and the first value >30 days later required to be above at least 50% of the threshold, and with no values within 30 days below 50% of the threshold c) date of first value, and the first value >30 days later required to be above at least 70% of the threshold, and with no values within 30 days below 70% of the threshold, for genetically confirmed cohort**

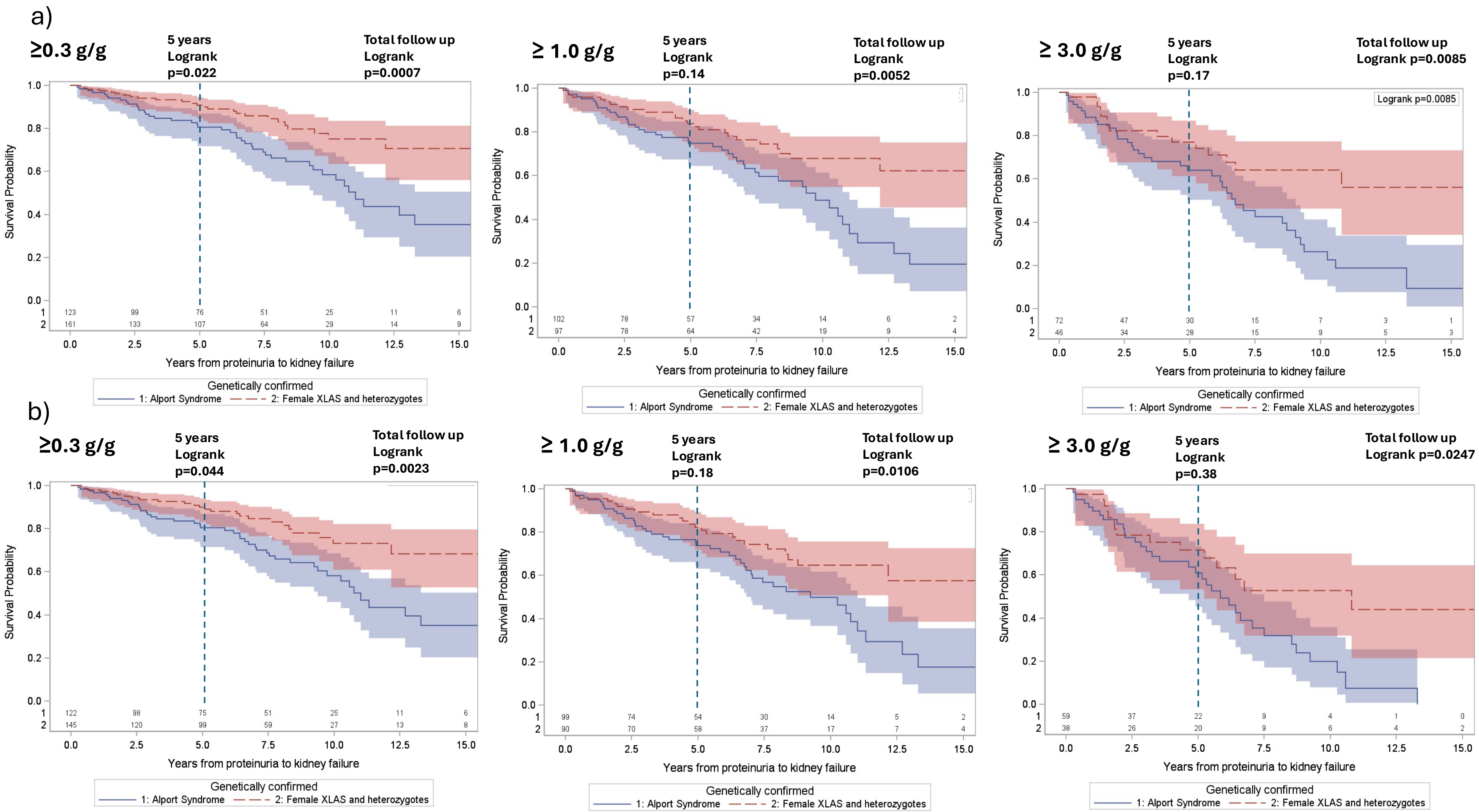

**Supplementary Figure 15 (Continued)**

c)

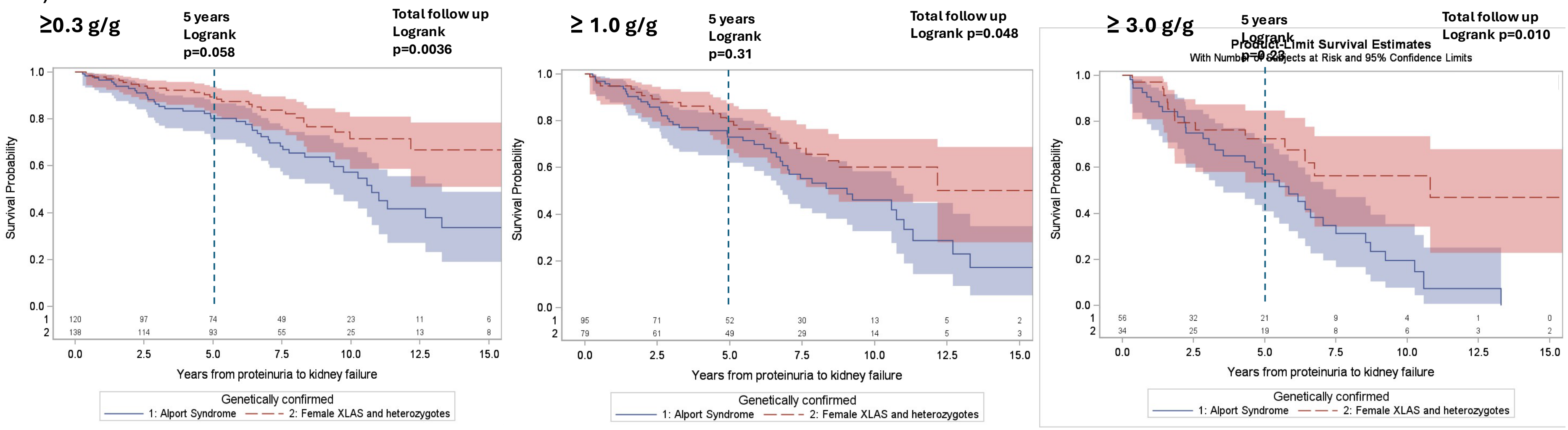

**Supplementary Figure 16: a) Cumulative plot of age at proteinuria thresholds for all patients and b) stratified by Alport Syndrome and Heterozygous genotypes, for clinically diagnosed cohort**

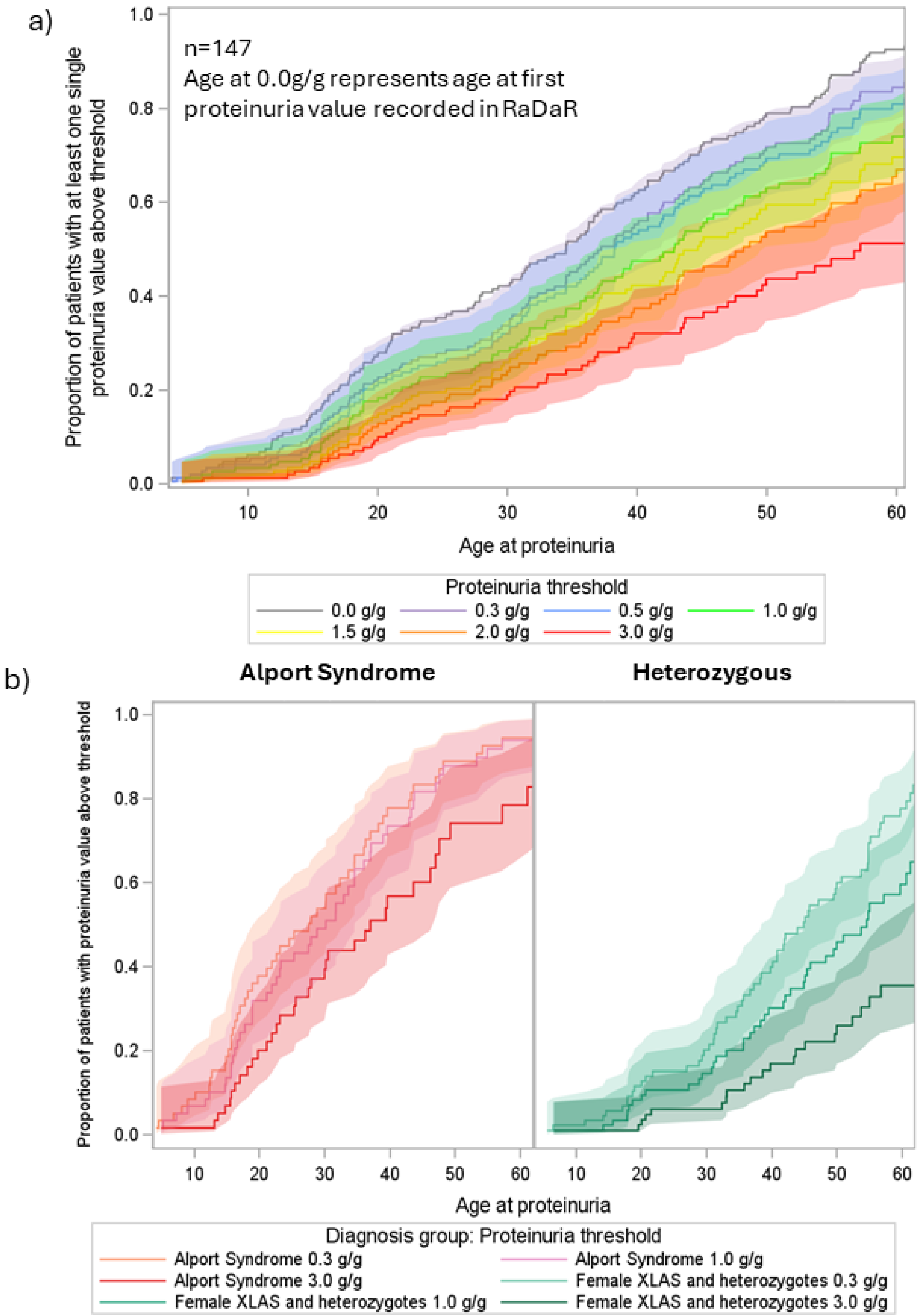

**Supplementary Figure 17: a) Kaplan Meier of time from proteinuria values to Kidney Failure, stratified by Alport Syndrome and heterozygous genotypes b) 25th centile time to kidney failure from exceeding proteinuria thresholds, by genotype for clinically diagnosed cohort**

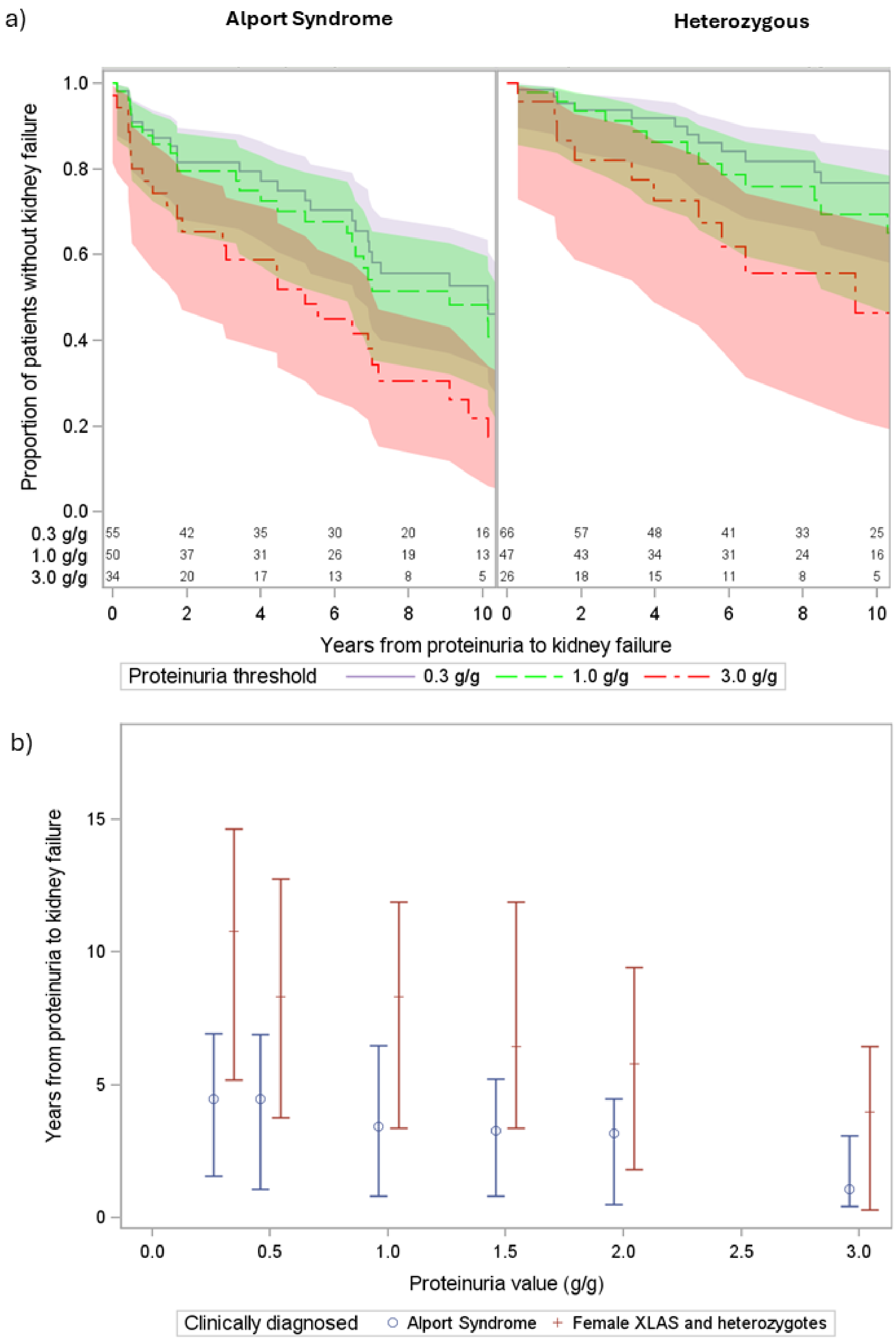


*for clinically diagnosed cohort only

**Supplementary Figure 18: Kaplan Meier plots of time from reaching proteinuria thresholds of a) ≥0.3g/g b) ≥1.0 g/g c) ≥3.0 g/g to kidney failure , where date of reaching a proteinuria threshold was defined as a) date of first single value above that threshold, b) date of first value, and the first value >30 days later required to be above at least 50% of the threshold, and with no values within 30 days below 50% of the threshold c) b) date of first value, and the first value >30 days later required to be above at least 70% of the threshold, and with no values within 30 days below 70% of the threshold, for clinically diagnosed cohort**

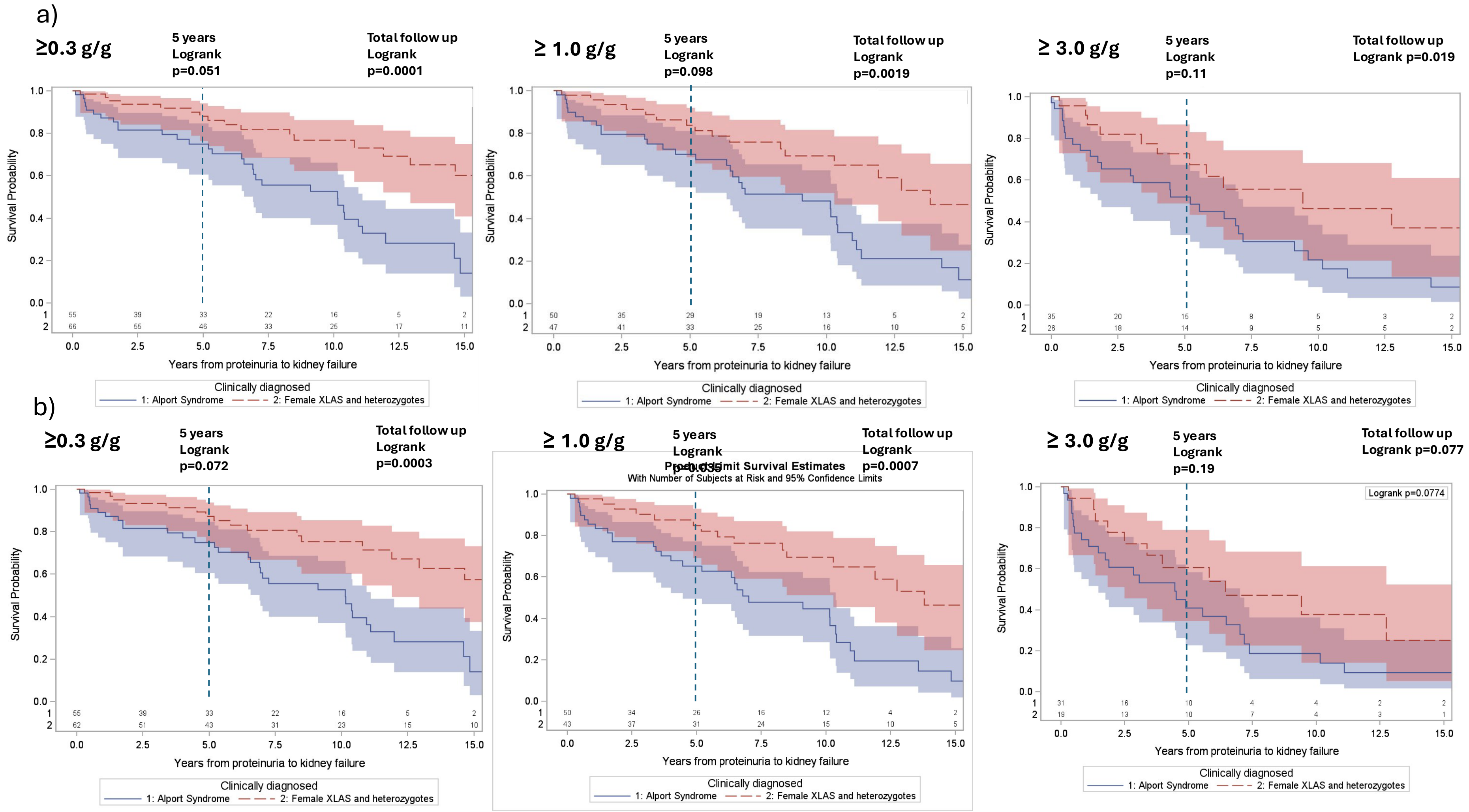

**Supplementary Figure 18 (Continued)**

c)

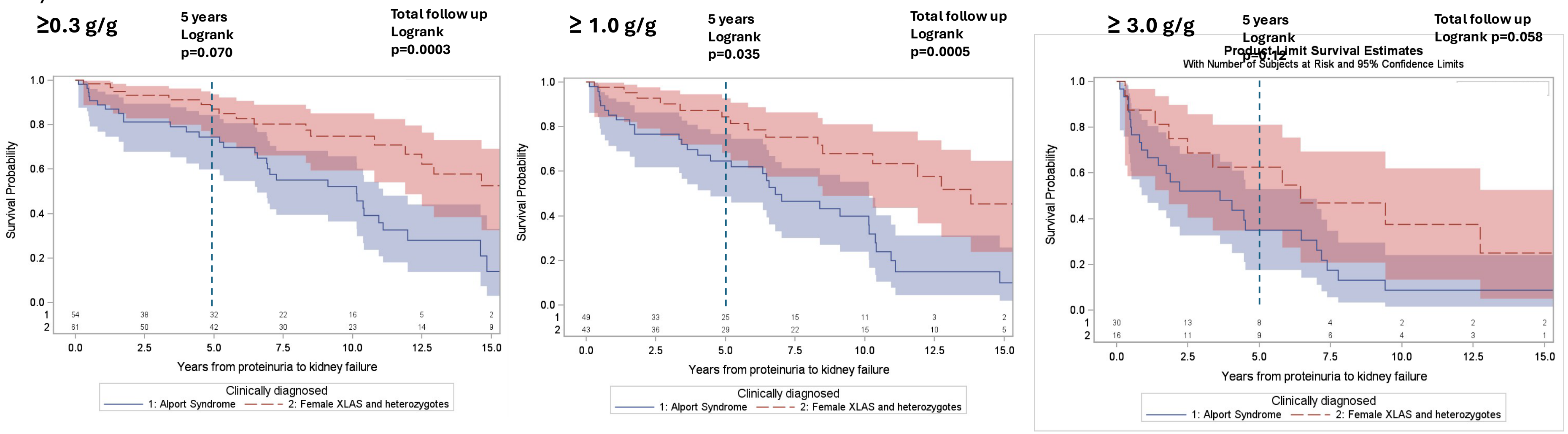

**Supplementary Figure 19: Time to kidney failure for patients with a confirmed proteinuria value >0.3g/g and those without, for patients with a) Alport Syndrome b) Heterozygous genotypes, for (i) genetically confirmed and (ii) clinically diagnosed cohorts**

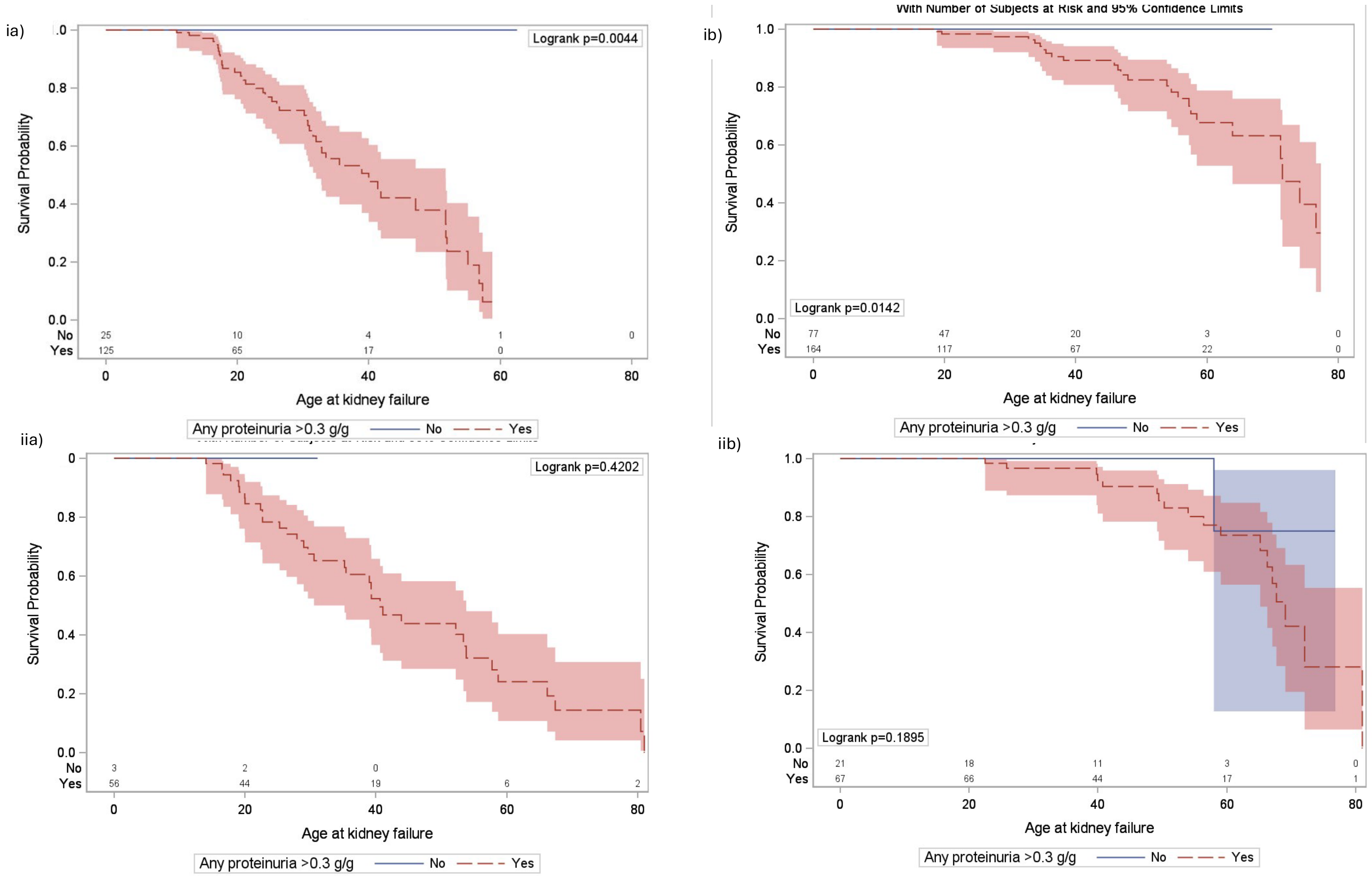

**Supplementary Figure 20: Time to KF from a) eGFR 90 b) eGFR 60 c) eGFR 45 ml/min/1.73m², stratified by median proteinuria in year prior to each eGFR threshold for i) Alport Syndrome and (ii) Heterozygous genotypes**

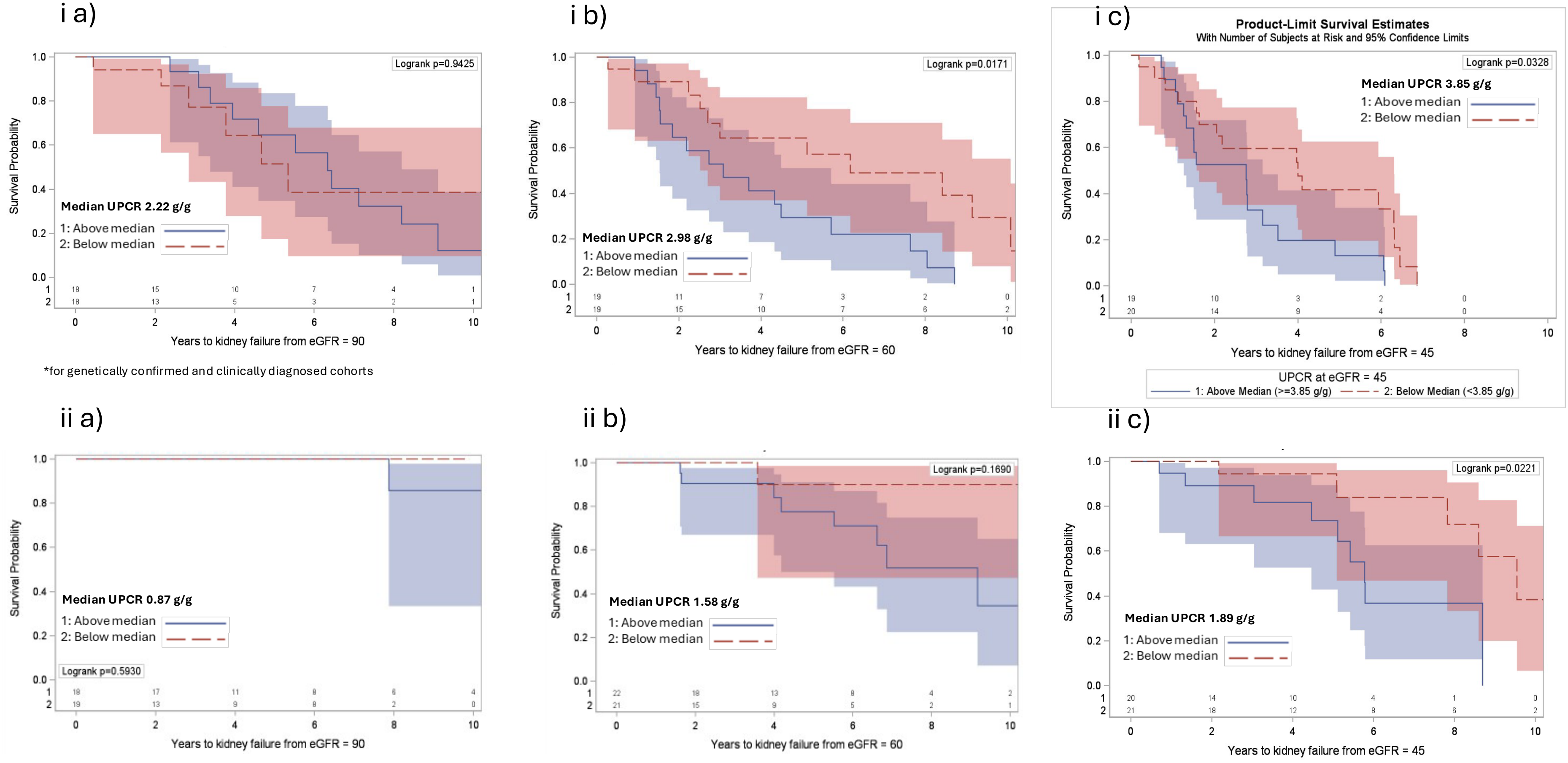


*including both genetically confirmed and clinically diagnosed cohorts

**National Registry of Rare Kidney Diseases (RaDaR) Consortium**

Sharirose Abat,[1] Shazia Adalat,[2] Atinuke Adedapo,[103] Joy Agbonmwandolor,[3] Zubaidah Ahmad,[4] Abdulfattah Alejmi,[5] Rashid Almasarwah,[6] Nicholas Annear,[1] Zainab Arslan[112], Ellie Asgari,[4] Samantha Ashleigh[84], Amanda Ayers,[7] Jyoti Baharani,[8] Gowrie Balasubramaniam,[9] Lauren Axon[84], Felix Jo-Bamba Kpodo,[10] Tarun Bansal,[11] Alison Barratt,[12] Jonathan Barratt[79], Megan Bates,[13] Natalie Bayne,[14] Janet Bendle,[15] Sarah Benyon,[16] Carsten Bergmann,[17,18] Sunil Bhandari,[19] Coralie Bingham,[20] Preetham Boddana,[21] Sally Bond,[22] Fiona Braddon,[23] Kate Bramham,[23] Angela Branson,[15] Stephen Brearey,[24] Vicky Brocklebank,[25] Sharanjit Budwal,[26] Conor Byrne,[27] Hugh Cairns,[28] Brian Camilleri[29], Gary Campbell[30], Alys Capell[31], Margaret Carmody,[8] Marion Carson[32], Tracy Cathcart,[19] Christine Catley,[9] Karine Cesar[33], Melanie Chan,[6] Houda Chea,[15] James Chess[34], Chee Kay Cheung,[26] Katy-Jane Chick[35], Nihil Chitalia[36], Martin Christian[37], Tina Chrysochou[38,39], Katherine Clark[40], Christopher Clayton[41], Rhian Clissold,[20] Helen Cockerill[33], Joshua Coelho[42], Elizabeth Colby[43], Viv Colclough[44], Eileen Conway[45], H. Terence Cook[46], Wendy Cook[47], Theresa Cooper[48], Richard J Coward[43], Sarah Crosbie,[22] Gabor Cserep[49], Sunil Daga[55], Anjali Date[50], Katherine Davidson[48], Amanda Davies[51], Neeraj Dhaun[52], Ajay Dhaygude[53], Lynn Diskin,[12] Abhijit Dixit[41,54], Eunice Ann Doctolero[35], Suzannah Dorey[55], Lewis Downard,[23] Mark Drayson[56], Gavin Dreyer,[27] Tina Dutt[57], Kufreabasi Etuk,[28] Dawn Evans[58], Jenny Finch[29], Frances Flinter[59], James Fotheringham[60], Lucy Francis[82], Daniel P. Gale[61], Hugh Gallagher[62], David Game,[4] Eva Lozano Garcia[42], Madita Gavrila,[22] Susie Gear[63], Colin Geddes[64], Mark Gilchrist[65], Matt Gittus[66], Paraskevi Goggolidou[67], Christopher Goldsmith[57], Patricia Gooden[68], Andrea Goodlife,[26] Priyanka Goodwin[53], Tassos Grammatikopoulos,[28,69] Barry Gray[70], Megan Griffith[46], Steph Gumus,[9] Sanjana Gupta[71], Patrick Hamilton[72], Lorraine Harper[56], Tess Harris[73], Louise Haskell[74], Samantha Hayward[43], Shivaram Hegde[75], Jennifer Henderson[114], Bruce Hendry[76], Sue Hewins[77], Nicola Hewitson[78], Kate Hillman,[15] Mrityunjay Hiremath[57], Alexandra Howson[79], Zay Htet,[28] Sharon Huish,[16] Richard Hull,[1] Alister Humphries[68], David P. J. Hunt[119], Karl Hunter[80], Samantha Hunter,[19] Marilyn Ijeomah-Orji,[6] Nick Inston[81], David Jayne[82], Gbemisola Jenfa[31], Alison Jenkins[83], Sally Johnson[118], Caroline A Jones[84], Colin Jones[85], Amanda Jones,[5] Rachel Jones[82], Lavanya Kamesh[81], Durga Kanigicherla[39], Fiona Karet Frankl[82], Mahzuz Karim[86], Amrit Kaur[87], David Kavanagh,[25] Kelly Kearley[88], Larissa Kerecuk,[14] Arif Khwaja[70], Garry King,[23] Grant King[89], Ewa Kislowska,[4] Edyta Klata[29], Maria Kokocinska,[14] Ania Koziell[2] Mark Lambie[90], Laura Lawless[41], Thomas Ledson[80], Rachel Lennon[91], Adam P Levine[92], Andrew Lewington[55], Ling Wai Maggie Lai,[16] Graham Lipkin[81], Graham Lovitt[93], Paul Lyons[94], Holly Mabillard[95], Katherine Mackintosh,[7] Khalid Mahdi[96], Eamonn Maher[97], Kevin J. Marchbank,[25] Patrick B Mark[64], Sherry Masoud,[23] Bridgett Masunda,[9] Zainab Mavani[31], Jake Mayfair,[4] Stephen McAdoo,[6] Joanna Mckinnell[98], Nabil Melhem,[2] Simon Meyrick[51], Shabbir Moochhala[61], Putnam Morgan[99], Ann Morgan[100,101], Fawad Muhammad,[5] Srividya Muppavaram[70], Shona Murray[30], Ailish Nimmo[83], Kristina Novobritskaya,[22] Albert CM Ong[66,70], Louise Oni[102], Kate Osmaston,[23] Neal Padmanabhan[64], Sharon Parkes,[14] Jean Patrick,[7] James Pattison,[4] Riny Paul,[1] Rachel Percival[103], Stephen J. Perkins[104], Alexandre Persu[105,106], William G Petchey[107], Matthew C. Pickering[46], Jennifer Pinney[81], David Pitcher,[23] Lucy Plumb[43], Zoe Plummer,[23] Joyce Popoola,[1] Frank Post,[28] Albert Power[83], Guy Pratt[56], Charles Pusey[46], Susan Pywell[23], Trijntje Rennie[119], Ria Rabara,[22] May Rabuya,[4]Tina Raju[42], Chadd Javier[108], Ian SD Roberts,[22] Candice Roufosse[109], Adam Rumjon,[28] Alan Salama[61], Moin Saleem[43], RN Sandford[97], Kanwaljit S. Sandu[110], Nadia Sarween[81], John A. Sayer[95], Neil Sebire[111,112], Haresh Selvaskandan,[26] Sapna Shah,[28] Asheesh Sharma[57], Edward J Sharples,[22] Neil Sheerin,[25] Harish Shetty[53], Rukshana Shroff[112], Roslyn Simms[70], Manish Sinha,[2] Smeeta Sinha[113], Kerry Smith[29], Lara Smith,[15] Shalabh Srivastava[114], Retha Steenkamp,[23] Ian Stott[115], Katerina Stroud[97], Pauline Swift[42], Justyna Szklarzewicz,[26] Fred Tam[46], Kay Tan[116], Robert Taylor[117], Marc Tischkowitz[97], Kay Thomas,[4] Yincent Tse[118], Alison Turnbull[85], A.

Neil Turner[119], Kay Tyerman[55], Miranda Usher[120], Gopalakrishnan Venkat-Raman[121], Alycon Walker[122], Stephen B. Walsh[61], Aoife Waters[123], Angela Watt[68], Phil Webster,[6] Ashutosh Wechalekar[124], Gavin Iain Welsh[43], Nicol West[125], David Wheeler[61], Kate Wiles,[27] Lisa Willcocks[107], Angharad Williams[33], Emma Williams[29], Karen Williams,[4] Deborah H Wilson[126], Patricia D. Wilson[127], Paul Winyard,[9] Edwin Wong,[25] Katie Wong,[23] Grahame Wood[58], Emma Woodward,[15] Len Woodward[128], Adrian Woolf[129], and David Wright

1 St George’s University Hospitals NHS Foundation Trust, UK
2 Evelina London Children's Hospital, UK
3 David Evans Medical Research Centre, Nottingham University Hospital NHS Trust, UK
4 Guy’s and St Thomas NHS Foundation Trust, UK
5 Ysbyty Gwynedd, Betsi Cadwaladr University Health Board, UK
6 Imperial College Healthcare NHS Trust, UK
7 James Paget University Hospital NHS Foundation Trust, UK
8 Heart of England NHS Foundation Trust, Birmingham, UK
9 Mid and South Essex NHS Foundation Trust, UK
10 Royal Berkshire NHS Foundation Trust, UK
11 Bradford Teaching Hospitals NHS Foundation Trust, UK
12 Royal United Hospital Bath NHS Trust, UK
13 Freeman Hospital, Newcastle Upon Tyne, UK
14 Birmingham Women's and Children's NHS Foundation Trust, UK
15 Manchester University NHS Foundation Trust, UK
16 Royal Devon University Healthcare NHS Foundation Trust, UK
17 Medizinische Genetik Mainz, Mainz, Germany
18 Department of Medicine, Faculty of Medicine, Medical Center-University of Freiburg, Freiburg, Germany
19 Hull University Teaching Hospitals NHS Trust, UK
20 Exeter Kidney Unit, Royal Devon University Healthcare NHS Foundation Trust, UK
21 Gloucestershire Hospitals NHS Foundation Trust, UK
22 Oxford University Hospitals NHS Foundation Trust, UK
23 UK Kidney Association, UK
24 Countess of Chester NHS Foundation Trust, UK
25 National Renal Complement Therapeutics Centre, Newcastle upon Tyne Hospitals NHS Foundation Trust, Newcastle upon Tyne, UK
26 University Hospitals of Leicester NHS Trust, UK
27 Barts Health NHS Trust, London, UK
28 King’s College Hospital NHS Foundation Trust, UK
29 East Suffolk and North Essex NHS Foundation Trust, UK
30 Ninewells Hospital and Medical School, Dundee, UK
31 North West Anglia NHS Foundation Trust, UK
32 Northern Health and Social Care Trust and Northern Ireland Clinical Research Network

33 West Suffolk NHS Foundation Trust, UK
34 Morriston Hospital, Swansea Bay Health Board, UK
35 Lister Hospital, East and North Hertfordshire NHS Trust, UK
36 Dartford and Gravesham NHS Trust, UK
37 Nottingham Children's Hospital, UK
38 Salford Royal Hospital, Northern Care Alliance NHS Foundation Trust, Salford, UK
39 University of Manchester, UK
40 King's College London, UK
41 Nottingham University Hospitals NHS Trust, UK
42 Epsom and St Helier University Hospitals NHS Trust, UK
43 University of Bristol Medical School, Bristol, UK
44 Royal Stoke University Hospital, UK
45 Manchester Royal Infirmary, UK
46 Centre for Inflammatory Disease, Imperial College London, UK
47 Nephrotic Syndrome Trust (NeST), UK
48 North Cumbria Integrated Care NHS Foundation Trust, UK
49 Colchester General Hospital, UK
50 Tameside and Glossop Integrated Care NHS Foundation Trust, UK
51 Wye Valley NHS Trust, UK
52 BHF Centre for Cardiovascular Science, The Queen's Medical Research Institute, University of Edinburgh, UK
53 Lancashire Teaching Hospital, UK
54 School of Medicine, University of Nottingham, UK
55 Leeds Teaching Hospitals NHS Trust, UK
56 University of Birmingham, UK
57 Liverpool University Hospitals Foundation NHS Trust, UK
58 Salford Royal NHS Foundation Trust, UK
59 Department of Clinical Genetics, Guy's and St Thomas' NHS Foundation Trust, UK
60 Centre for Health and Related Research, School of Population Health, University of Sheffield, UK
61 University College London Department of Renal Medicine, Royal Free Hospital, UK
62 SW Thames Renal Unit, Epsom and St Helier University Hospitals NHS Trust, UK
63 Alport UK, UK
64 Queen Elizabeth University Hospital, Glasgow, UK

65 College of Medicine and Health, University of Exeter, UK
66 Division of Population Health, University of Sheffield, UK
67 University of Wolverhampton, UK
68 Patient Representative, UK
69 Institute of Liver Studies, King's College London, UK
70 Sheffield Kidney Institute, Sheffield Teaching Hospitals NHS Foundation Trust, UK
71 Royal Free Hospital, UK
72 Manchester Institute of Nephrology and Transplantation, Manchester Royal Infirmary, UK
73 PKD Charity, UK
74 University Hospital Southampton NHS Foundation Trust, UK
75 Children's Kidney Centre, University Hospital of Wales, UK
76 Travere Therapeutics, UK
77 University Hospitals Coventry and Warwickshire NHS Trust, UK
78 County Durham & Darlington NHS Foundation Trust, UK
79 University of Leicester, UK
80 Wirral University Teaching Hospital NHS Foundation Trust, UK
81 University Hospitals Birmingham NHS Foundation Trust, UK
82 Department of Medicine, University of Cambridge, UK
83 North Bristol NHS Trust, UK
84 Alder Hey Children's NHS Foundation Trust, UK
85 York & Scarborough Teaching Hospitals NHS Foundation Trust, UK
86 Norfolk and Norwich University Hospitals NHS Trust, UK
87 Royal Manchester Children's Hospital, Manchester, UK
88 PTEN UK and Ireland Patient Group
89 HNF1B Support Group, UK
90 School of Medicine, Keele University, UK
91 Wellcome Centre for Cell-Matrix Research, University of Manchester, UK
92 Research Department of Pathology, University College London, UK
93 HLRCC Foundation, UK
94 Cambridge Institute of Therapeutic Immunology and Infectious Disease, Cambridge, UK
95 Newcastle University, UK
96 United Lincolnshire Hospitals NHS Trust, UK
97 Department of Medical Genetics, University of Cambridge, UK
98 University Hospitals of Derby and Burton NHS Foundation Trust, UK

99 Retroperitoneal Fibrosis (RF) Group, UK
100 National Institute of Health and Care Research Leeds Biomedical Research Centre, Leeds Teaching Hospitals NHS Trust, UK
101 School of Medicine, University of Leeds, UK
102 University of Liverpool, UK
103 Newcastle Upon Tyne Hospitals NHS Foundation Trust, UK
104 Research Department of Structural and Molecular Biology, University College London, UK
105 Division of Cardiology, Cliniques Universitaires Saint-Luc, Belgium
106 Pole of Cardiovascular Research, Institut de Recherche Expérimentale et Clinique, Université Catholique de Louvain, Brussels, Belgium
107 Cambridge University Hospitals NHS Foundation Trust, UK
108 East and North Hertfordshire NHS Trust, UK
109 Department of Immunology and Inflammation, Faculty of Medicine, Imperial College London, UK
110 Shrewsbury and Telford Hospital NHS Trust, UK
111 National Institute of Health and Care Research Great Ormond Street Hospital Biomedical Research Centre, UK
112 UCL Great Ormond Street Institute of Child Health, UK
113 Northern Care Alliance NHS Foundation Trust, UK
114 South Tyneside and Sunderland NHS Foundation Trust, UK
115 Doncaster and Bassetlaw Teaching Hospitals, UK
116 New Cross Hospital, Wolverhampton, UK
117 Wellcome Centre for Mitochondrial Research, Translational & Clinical Research Institute, Faculty of Medical Sciences, Newcastle University, UK
118 Great North Children's Hospital, Newcastle Upon Tyne, UK
119 University of Edinburgh, UK
120 Calderdale & Huddersfield Foundation Trust, UK
121 Royal Surrey County Hospital, Guildford, UK
122 South Tees Hospitals NHS Foundation Trust, UK
123 University College Cork, Ireland
124 National Amyloidosis Centre, University College London, UK
125 Great Western Hospital, Swindon, UK
126 North Tees and Hartlepool NHS Foundation Trust, UK
127 University College London, UK

128 aHUS Alliance, UK

129 School of Biological Sciences, University of Manchester, UK

STROBE Statement—Checklist of items that should be included in reports of ***cohort studies***

| | Item No | Recommendation |
|---|---|---|
| **Title and abstract** | 1 | **(*a*) Indicate the study's design with a commonly used term in the title or the abstract**<br><br>Page 1: "Quantifying associations of genotype, proteinuria and eGFR with long-term kidney outcomes in Alport Syndrome using data from the UK National Registry of Rare Kidney Diseases (RaDaR)" |
| | | **(*b*) Provide in the abstract an informative and balanced summary of what was done and what was found**<br><br>Page 3: *Methods* In this retrospective cohort study of individuals with AS in the UK National Registry of Rare Kidney Diseases, patients were classified as having AS or heterozygous genotypes and followed to assess proteinuria progression, eGFR slope and kidney survival. Proteinuria and eGFR trajectories were analysed using mixed-effects regression models; kidney survival using Kaplan–Meier analysis.<br><br>Page 3: *Results* Among 1032 participants (median follow-up 11.6 years; 47% female), 475 (46%) had AS genotypes (Male XLAS or autosomal recessive AS). eGFR decline accelerated with advancing chronic kidney disease (CKD) stage across all genotypes (p<0.001). Proteinuria increased as eGFR declined and occurred earlier in AS genotypes. After reaching proteinuria thresholds of ≥1.0 and ≥3.0g/g, kidney survival over the subsequent 5-years did not differ significantly between genotypes (log-rank p=0.14, p=0.17, respectively), although modest differences emerged over longer follow-up. Across eGFR thresholds (90, 60, and 45mL/min/1.73m$^2$), higher proteinuria was associated with shorter time to KF; for example, at eGFR 45mL/min/1.73m$^2$, median time to KF was 3.0 years (IQR, 1.6–5.4) for above-median vs 6.5 years (5.1–not estimable) for below-median proteinuria (p<0.0001). Almost all patients who reached KF had developed proteinuria ≥0.3g/g. |
| **Introduction** | | |
| Background/rationale | 2 | **Explain the scientific background and rationale for the investigation being reported**<br><br>Page 5: "Clinical outcomes vary widely, even within families, and limited long-term prognostic data can leave patients and clinicians uncertain and anxious about expected disease course" AND "Data on rate of kidney function decline prior to KF in AS, and whether this differs by genotype or across disease stage, are limited." AND "Proteinuria is associated with adverse kidney outcomes[13] but, whilst its prognostic value in non-monogenic glomerular disorders such as IgA nephropathy (IgAN)[14,15] idiopathic nephrotic syndrome (INS)[16], and immune-complex MPGN/C3 glomerulopathy[17,18] has been well delineated, there are minimal studies reporting longitudinal trajectories of proteinuria[19] in AS. Previous studies have used "baseline" proteinuria values at arbitrary time-points (e.g. "first clinic visit")[10,13,20] these values may represent patients at different points in their disease course and with differing levels of kidney function, making these results difficult to interpret. This question has become more important with recent therapeutic developments in AS include a clinical trial of lademirsen[21], and studies evaluating broader proteinuria-lowering therapies such as SGLT2-inhibitors[22] and |

| | | |
|---|---|---|
| | | endothelin-targeted therapies[22–2422]Meta-analyses in other kidney diseases support short-term changes in eGFR slope and proteinuria as surrogate end-points for long-term KF [15,25–29] , but whether, and how, these findings can be extrapolated to AS remains uncertain. |
| Objectives | 3 | **State specific objectives, including any prespecified hypotheses**<br><br>Page 6: "We therefore examined associations of genotype, proteinuria and eGFR with long-term renal outcomes in 1008 individuals with AS recruited to the UK National Registry of Rare Kidney Diseases (RaDaR)". |
| **Methods** | | |
| Study design | 4 | **Present key elements of study design early in the paper**<br><br>Page 3: "In this retrospective cohort study..." |
| Setting | 5 | **Describe the setting, locations, and relevant dates, including periods of recruitment, exposure, follow-up, and data collection**<br><br>Page 6: "*Data source and study population* RaDaR recruits patients with rare kidney disease from 108 UK National Health Service sites with informed consent. AS recruitment began in 2013. Data were extracted on August 12, 2025. Study design, eligibility criteria, data linkage, and variable definitions have been previously reported[30–32], with details in Supplementary Methods AND<br><br>Supplementary Methods |
| Participants | 6 | **(*a*) Give the eligibility criteria, and the sources and methods of selection of participants. Describe methods of follow-up**<br><br>Page 6: "Study design, eligibility criteria, data linkage, and variable definitions have been previously reported[30–32], with further details in Supplementary Methods." AND<br><br>Supplementary Materials, Page 2: "Supplementary Methods : *Eligibility criteria*<br><br>The RaDaR Alport Syndrome cohort began recruitment in 2013. Eligibility criteria defined at that time included: a) AS definite or probable b) Alport carrier definite or probable c) Female heterozygote for XLAS (*COL4A5*) d) Heterozygote for autosomal AS (*COL4A3*/*COL4A4*) e) Thin basement membrane nephropathy (TBMN). *Data source and data linkages* Data linkage with local hospitals and renal units enables retrospective and automated prospective collection of blood and urine results via the UK Renal Data Collaboration (UKRDC). Linkage with the UK Renal Registry (UKRR) provides validated data on kidney replacement therapy (KRT) initiation (including NHS Blood and Transplant data). RaDaR receives clinical genetic reports from NHS genomics hubs." AND<br><br>"Follow up time was classified as time from diagnosis to date of data extraction, or death." |
| | | ~~(*b*) For matched studies, give matching criteria and number of exposed and unexposed~~ |

| | | |
|---|---|---|
| Variables | 7 | **Clearly define all outcomes, exposures, predictors, potential confounders, and effect modifiers. Give diagnostic criteria, if applicable**<br><br>Page 6: "...variable definitions have been previously reported[30–32], with further details in Supplementary Methods." AND<br><br>Supplementary Materials, Page 2: "Supplementary Methods : *Variable and outcome definitions table"* |
| Data sources/ measurement | 8* | **For each variable of interest, give sources of data and details of methods of assessment (measurement). Describe comparability of assessment methods if there is more than one group**<br><br>Supplementary Materials, Page 2: "*Data source and data linkages*<br><br>Data linkage with local hospitals and renal units enables retrospective and automated prospective collection of blood and urine results via the UK Renal Data Collaboration (UKRDC). Linkage with the UK Renal Registry (UKRR) provides validated data on kidney replacement therapy (KRT) initiation (including NHS Blood and Transplant data). RaDaR receives clinical genetic reports from NHS genomics hubs." |
| Bias | 9 | **Describe any efforts to address potential sources of bias**<br><br>Page 7: "Variant details were cross-checked against contemporary databases (description in Supplementary Methods)." AND "Sensitivity analyses using alternative eGFR equations and proteinuria definitions are described in supplementary materials." |
| Study size | 10 | **Explain how the study size was arrived at**<br><br>Page 7: "Patients were included in the 'Genetically Confirmed' group if testing in the patient or a parent identified a variant classified as pathogenic (P) or likely pathogenic (LP) by American College of Medical Genetics criteria[30] (Supplementary Figure 1). The 'Clinical Diagnosis' group comprised patients where diagnosis was made based on family history, histopathology and clinical features, with genetic testing either not performed or where no P/LP variant was identified.<br><br>Patients in both groups were then categorised as having either an 'Alport Syndrome' (male XLAS or homozygous or biallelic *COL4A3*/*COL4A4* variants, ARAS) or 'Heterozygous' genotypes, (heterozygous *COL4A3*/*COL4A4* variants, TBMN and female XLAS), with the definition of Alport Syndrome genotypes consistent with current naming guidance[33].<br><br>AND<br><br>Supplementary Figure 1. |
| Quantitative variables | 11 | **Explain how quantitative variables were handled in the analyses. If applicable, describe which groupings were chosen and why**<br><br>Page 7: "Definitions of diagnosis date, follow-up, time-averaged proteinuria, and KF are provided in the Supplement."<br><br>And Supplementary Materials, Page 2: "Supplementary Methods : *Variable and outcome definitions table"* |

| | | |
|---|---|---|
| Statistical methods | 12 | **(*a*) Describe all statistical methods, including those used to control for confounding**<br><br>Page 8-9, section "*Statistical Analyses"* |
| | | **(*b*) Describe any methods used to examine subgroups and interactions**<br><br>Page 7-8, section "*Statistical Analyses:* Kaplan-Meier analysis and log-rank testing were used to compare age at KF and time from diagnosis to KF by genotype and variant category. Time to KF from sustained eGFR thresholds of 90, 60, and 45 mL/min/1.73 $m^2$ was estimated according to median proteinuria in the preceding year. eGFR slope and proteinuria trajectories were modelled using mixed-effects regression." AND<br><br>Supplementary Materials, Supplementary Methods Page 4 "*eGFR slope and proteinuria trajectory calculations*<br><br>eGFR slope was estimated using mixed-effects regression models with a random intercept and slope for each patient. Interaction terms between slope, CKD stage and mutation type were included in the models to allow slope within each CKD stage to be estimated for each group. Sensitivity analyses using the CKD-EPI 2021, or the bedside Schwartz for those aged <16 years, instead of the European eGFR are presented in Supplementary Figure 8. The relationship between log-transformed proteinuria and age was modelled using a mixed-effects regression model to account for repeated measures within individuals, firstly with the assumption that the relationship was linear, and then, to account for non-linearity, estimating age effects using a cubic spline with knots at the $25^{th}$, $50^{th}$ and $75^{th}$ percentiles of the age distribution." |
| | | **(*c*) Explain how missing data were addressed**<br><br>Page 9: "Available data are presented in Supplementary Table 2; analyses used complete cases", and Supplementary Table 2 |
| | | **(*d*) If applicable, explain how loss to follow-up was addressed**<br><br>N/A |
| | | **(*e*) Describe any sensitivity analyses**<br><br>Page 7: "Sensitivity analyses using alternative eGFR equations and proteinuria definitions are described in the Supplement." AND Supplementary Materials, Supplementary Methods Page 4: "*Methodology for proteinuria thresholds*... Sensitivity analyses where date at reaching a proteinuria threshold was defined as date of the first value above a certain threshold were performed with a) the first value >30 days later required to be above at least 50% of the threshold, and with no values within 30 days below 50% of the threshold and b) using a 70% of threshold cut-off." AND "*eGFR slope and proteinuria trajectory calculations*... Sensitivity analyses using the CKD-EPI 2021, or the bedside Schwartz for those aged <16 years, instead of the European eGFR are presented in Supplementary Figure 8." |

**Results**

| | | |
|---|---|---|
| Participants | 13* | **(a) Report numbers of individuals at each stage of study—eg numbers potentially eligible, examined for eligibility, confirmed eligible, included in the study, completing follow-up, and analysed**<br><br>Page 8 "A total of 1032 individuals with AS were included (aged 3 to 92 years; 481 (47%) female) with median follow-up 11.6 years (IQR 7.6-19.1)" AND 3 patients with two affected Alport genes or *MYH9* variants were excluded.<br><br>AND Table 1. |
| | | **(b) Give reasons for non-participation at each stage**<br><br>N/A- informed consent from all patients. Data availability in Supplementary Table 2. |
| | | (c) Consider use of a flow diagram<br><br>Supplementary Figure 1 |
| Descriptive data | 14* | **(a) Give characteristics of study participants (eg demographic, clinical, social) and information on exposures and potential confounders**<br><br>Table 1, Supplementary Table 3 |
| | | **(b) Indicate number of participants with missing data for each variable of interest**<br><br>Supplementary Table 2 |
| | | **(c) Summarise follow-up time (eg, average and total amount)**<br><br>Page 8 "A total of 1032 individuals with AS were included (aged 3 to 92 years; 481 (47%) female) with median follow-up 11.6 years (IQR 7.6-19.1)" AND 3 patients with two affected Alport genes or *MYH9* variants were excluded.<br><br>AND Table 1. |
| Outcome data | 15* | **Report numbers of outcome events or summary measures over time**<br><br>Table 2, row "Kidney Failure (KF) events |
| Main results | 16 | **(*a*) Give unadjusted estimates and, if applicable, confounder-adjusted estimates and their precision (eg, 95% confidence interval). Make clear which confounders were adjusted for and why they were included**<br><br>Page 8: Results section. All results presented with 95% CI or Interquartile Range (IQR) Supplementary Materials, Supplementary Methods "*eGFR slope and proteinuria trajectory calculations"* |
| | | **(*b*) Report category boundaries when continuous variables were categorized**<br><br>Page 8: "Time to KF from sustained eGFR thresholds of 90, 60, and 45 mL/min/1.73 $m^2$ was estimated according to median proteinuria in the preceding year." |
| | | **(*c*) If relevant, consider translating estimates of relative risk into absolute risk for a meaningful time period** |

| | | |
|---|---|---|
| | | Table 2, row "10-year kidney survival from diagnosis (years, IQR)" and "10-year survival from reaching eGFR 30 from 90ml/min/1.73m$^2$, (years, IQR)" |
| Other analyses | 17 | **Report other analyses done—eg analyses of subgroups and interactions, and sensitivity analyses**<br><br>All analyses performed are reported |
| **Discussion** | | |
| Key results | 18 | **Summarise key results with reference to study objectives**<br><br>Page 13: "In this large national cohort of individuals with Alport syndrome (AS), we identified consistent and clinically relevant relationships between genotype, proteinuria, and progression to KF. Across all genotypes, decline in kidney function was non-linear and accelerated with advancing CKD stage. Although genotype influenced the age at onset and early disease trajectory, proteinuria was strongly associated with subsequent risk of KF across a range of eGFR thresholds. Notably, once comparable levels of proteinuria were reached, short- to medium-term kidney outcomes were similar across genotype groups" |
| Limitations | 19 | **Discuss limitations of the study, taking into account sources of potential bias or imprecision. Discuss both direction and magnitude of any potential bias**<br><br>Page 16: "Several limitations should be considered. First, this was an observational study, and although the relationship between outcome and proteinuria was strong, we do not provide evidence (or claim) that proteinuria itself causes kidney damage – it may simply reflect cumulative glomerular injury burden. . Proteinuria may reflect underlying disease severity as well as treatment effects, and whilst medication data were incomplete so we could not account fully for time-varying therapies, including renin–angiotensin system blockade and SGLT2 inhibitor use, for those with data available, ACE-inhibitor or Angiotensin Receptor Blocker use was comparable with other retrospective studies in AS[2] (Supplementary Table 3). Second, approximately half of participants lacked a genetically confirmed diagnosis, introducing potential misclassification, although results were consistent in clinically diagnosed individuals. Third, as a UK-based cohort, results may not be fully generalizable to other populations. Finally, missing data and variability in measurement frequency inherent to real-world datasets may have influenced estimates of proteinuria and eGFR trajectories" |
| Interpretation | 20 | **Give a cautious overall interpretation of results considering objectives, limitations, multiplicity of analyses, results from similar studies, and other relevant evidence**<br><br>Page 17: Conclusion "In this large cohort of individuals with AS, proteinuria was strongly associated with progression to KF across genotypes and levels of kidney function, and differences in outcomes by genotype were attenuated once comparable levels of proteinuria were reached. These findings identify proteinuria as a key prognostic marker in AS and support its use in clinical risk stratification and potential to act as a surrogate endpoint for kidney failure in future AS trials" AND |

| | | |
|---|---|---|
| | | Page 16: |
| Generalisability | 21 | **Discuss the generalisability (external validity) of the study results**<br><br>Page 9: “Third, as a UK-based cohort, results may not be fully generalizable to other populations.” “Several limitations should be considered. First, this was an observational study, and although the relationship between outcome and proteinuria was strong, we do not provide evidence (or claim) that proteinuria itself causes kidney damage – it may simply reflect cumulative glomerular injury burden. . Proteinuria may reflect underlying disease severity as well as treatment effects, and whilst medication data were incomplete so we could not account fully for time-varying therapies, including renin–angiotensin system blockade and SGLT2 inhibitor use, for those with data available, ACE-inhibitor or Angiotensin Receptor Blocker use was comparable with other retrospective studies in AS[2] (Supplementary Table 3). Second, approximately half of participants lacked a genetically confirmed diagnosis, introducing potential misclassification, although results were consistent in clinically diagnosed individuals. Third, as a UK-based cohort, results may not be fully generalizable to other populations. Finally, missing data and variability in measurement frequency inherent to real-world datasets may have influenced estimates of proteinuria and eGFR trajectories” |
| **Other information** | | |
| Funding | 22 | **Give the source of funding and the role of the funders for the present study and, if applicable, for the original study on which the present article is based**<br><br>Page 23: Funding |

*Give information separately for exposed and unexposed groups.

**Note:** An Explanation and Elaboration article discusses each checklist item and gives methodological background and published examples of transparent reporting. The STROBE checklist is best used in conjunction with this article (freely available on the Web sites of PLoS Medicine at http://www.plosmedicine.org/, Annals of Internal Medicine at http://www.annals.org/, and Epidemiology at http://www.epidem.com/). Information on the STROBE Initiative is available at http://www.strobe-statement.org.
